# Effects of low-dose iron supplementation on iron status, safety outcomes and gut microbiota in female soccer players: a randomized controlled study

**DOI:** 10.64898/2026.08.19.26360791

**Authors:** Stine Sofie Strømland, Trude Elise Aspholm, Gøran Paulsen, Monica Hauger Carlsen, Lisa Medina Grimestad, Anne Mari Herfindal, Anu Koivisto-Mørk, Nasser Ezzatkhah Bastani, Knut Rudi, Jørgen Valeur, Truls Raastad, Siv Kjølsrud Bøhn

## Abstract

**Purpose:** Iron deficiency impairs sports performance, and female athletes are particularly vulnerable. High-dose iron supplements, commonly used to prevent iron depletion and performance impairments, may cause gastrointestinal side effects and disrupt the gut microbiota. Whether lower doses can improve iron status without adverse effects remains unclear. The aim of this study was to characterize iron intake and iron status in female soccer players during the competitive season and investigate effects of low-dose iron supplementation on iron status, safety-related outcomes and gut microbiota.

**Methods:** In a two-arm parallel randomized controlled trial, female soccer players (median age 21) were randomized to an intervention group (n=12) receiving 3-month low-dose iron supplementation (27 mg elemental iron/day) or a control group (n=11) without supplementation. Blood/fecal samples were collected at baseline and 3-month follow-up. Dietary intake was estimated using 7-day food diaries. Between-group differences were analyzed per protocol (n=18) using ANCOVA with baseline adjustment.

**Results:** The players had inadequate baseline iron intake (median 11.2 mg/day) and 43% had serum ferritin indicating iron depletion (<35 µg/L). At follow-up, no significant between-group difference was found for serum ferritin, but fewer athletes in the intervention group experienced decreases from baseline to follow-up (*P*<0.05). Moreover, serum iron was higher in the intervention group (*P*_group_=0.05). No between-group differences were observed for gastrointestinal symptoms or liver damage biomarkers. On the contrary, the intervention led to lower IL-6 (*P*_group_=0.04) and higher gut microbial α-diversity (*P*_group_=0.01) compared to controls.

**Conclusions:** The low-dose iron supplementation was well tolerated, attenuated decreases in iron stores, increased gut microbial diversity and attenuated systemic inflammation in female soccer players with suboptimal dietary iron intake. However, potential adverse effects of long-term exposure cannot be excluded.

**Clinical Trial Registration:** This trial was registered at clinicaltrials.gov as NCT04526678, first submitted on 2020-08-13 (https://clinicaltrials.gov/study/NCT04526678).

## INTRODUCTION

Iron is a key micronutrient in sports nutrition, playing essential roles in oxygen transport and storage (as part of hemoglobin and myoglobin, respectively) and in energy production (as part of cytochromes in the electron transport chain) (1). Yet, iron deficiency remains one of the most common micronutrient deficiencies among athletes, particularly female athletes (2), and can impair both physical and cognitive performance (3). Because the human body cannot synthesize iron, iron status depends on efficient recycling and sufficient dietary intake to replace losses (4). According to the Nordic Nutrition Recommendations from 2023 (NNR-2023) (5), the average requirement (AR) and recommended intake (RI) for adult females are 9 and 15 mg iron/day, respectively. However, these recommendations may be insufficient for female athletes who face additional iron losses from exercise (2). It has been suggested that endurance athletes may require 30–70% more iron (6, 7) due to increased erythropoiesis, inflammation-induced reductions in iron absorption and recycling, and increased exercise-related iron losses through sweating, mechanical hemolysis (e.g. foot-strike hemolysis), hematuria and gastrointestinal (GI) bleeding (2, 8).

Soccer is a high-intensity intermittent sport that places considerable demands on endurance, strength and power (9). Reports of a high prevalence of iron depletion in female players (10, 11) and declines in iron status during the competitive season (12) suggest that regular iron monitoring, as recommended for endurance athletes (1, 2), may also be warranted in soccer. Iron status is usually assessed using multiple biomarkers that reflect different aspects of iron metabolism, including ferritin, hemoglobin, total iron-binding capacity (TIBC) and transferrin saturation (3). Ferritin, the iron storage protein, is the most widely used indicator of iron deficiency (1). In the general population, deficiency is typically defined by ferritin concentrations below 15 µg/L (13, 14); however, the optimal cut-off for athletes remains debated and previous studies have used thresholds ranging from 12 to 40 µg/L (15, 16).

Peeling et al. (15) describe three stages of iron deficiency with increasing severity: (1) iron depletion (ferritin < 35 µg/L), (2) iron-deficient erythropoiesis (ferritin < 20 µg/L) and (3) iron-deficient anemia (ferritin < 12 µg/L). In the final stage, iron stores and circulating iron no longer support adequate erythropoiesis (16), resulting in low hemoglobin levels (< 11.7 g/dL) (14) and impaired oxygen delivery. While iron deficiency anemia is well known to impair performance, the functional consequences of nonanemic iron deficiency (stages 1 and 2) are more equivocal (2) but may still negatively affect athletic performance (17, 18).

Over the years, many athletes have used iron supplements in the belief that they enhance physical performance (3, 16), despite limited evidence that supranormal iron levels offer any additional benefit (3). On the contrary, uncritical use of high-dose supplements may lead to iron overload as the human body lacks an active mechanism for iron excretion (19). Iron’s ability to act as both an electron donor (ferrous iron, Fe^2+^) and acceptor (ferric iron, Fe^3+^) makes free (non-transferrin-bound) iron highly reactive and capable of promoting oxidative stress through free radical formation (20). Iron supplementation is also commonly associated with adverse side effects such as nausea, constipation and abdominal pain (21), possibly related to disturbances in the gut microbiota (22, 23). Most gut bacteria require iron for growth and increased luminal iron availability may alter the gut microbiota composition, potentially favoring growth of pathogenic species and thereby increasing susceptibility to infections (24, 25). However, insufficient iron availability may also disturb microbial balance (26), suggesting that both deficiency and excess could have adverse effects on the gut microbiota.

The effect of iron supplementation on iron status appears to depend largely on baseline ferritin concentrations and study duration, with beneficial effects reported in iron-deficient athletes receiving 16–100 mg of elemental iron daily for six to eight weeks (27). However, dose is an important consideration, as fractional iron absorption decreases with increasing doses, resulting in more unabsorbed iron reaching the intestinal lumen (28). In addition, high doses may stimulate hepcidin synthesis, the key iron-regulatory peptide, which in turn reduces intestinal absorption and iron release from stores (20). This hepcidin response is attenuated at iron doses ≤ 40 mg/day (29), suggesting that a low-dose strategy may optimize absorption efficiency while minimizing potential adverse effects of unabsorbed intestinal iron (22). To date, only one randomized controlled trial has investigated the effects of iron supplementation in female soccer players, reporting that 40 mg of elemental iron daily for four weeks increased iron stores and prevented hemoglobin declines in iron-deficient athletes (30). Furthermore, while some studies indicate that certain pathogenic bacteria thrive with increased iron intake (22), data on the effects of iron supplementation on gut microbiota in humans remain scarce and inconsistent (26) and, to the best of our knowledge, no studies have examined this in athletes.

Thus, the major aims of the current randomized controlled study were to: (1) characterize the dietary iron intake and iron status in young female soccer players, and (2) investigate the effects of a low-dose iron supplementation on iron status biomarkers, safety-related outcomes and gut microbiota. Additionally, we assessed the relationship between bacterial taxa abundances and other study outcomes to explore potential underlying mechanisms.

## METHODS

### Study design and participants

The current study followed a two-arm parallel randomized controlled design and was conducted from August to November 2020. Female tier 3 (highly trained/national level) (31) soccer players were recruited from two soccer clubs in the Oslo region of Norway and followed from midseason in August (baseline) to late season in November (3-month follow-up) (**Supplementary Figure S1**, Supplemental Digital Content 1, study design). The primary outcome was serum ferritin at 3-month follow-up. Secondary outcomes were additional iron status biomarkers (serum iron, transferrin saturation, TIBC, hemoglobin concentration, red blood cells, hepcidin), liver damage biomarkers (aspartate aminotransferase (ASAT), alanine aminotransferase (ALAT), gamma-glutamyl transferase (GGT)), inflammatory biomarkers (C-reactive protein (CRP), neutrophil gelatinase-associated lipocalin (NGAL), lipopolysaccharide-binding protein (LBP), cytokines) and gut microbiota profile (microbial diversity, taxa relative abundances, short-chain fatty acids (SCFA)) at 3-month follow-up. We also estimated dietary iron intake at baseline using a digital 7-day food diary. Although we had planned to assess the effects of the intervention on physical performance, the available data from the Yo-Yo intermittent recovery test level 1 were unfortunately insufficient for analysis.

The inclusion criterion for the study was being a female soccer player from one of two selected soccer clubs in the Oslo region of Norway. Exclusion criteria were: i) pregnancy, ii) medical conditions that are worsened by taking iron supplements (e.g. hereditary hemochromatosis), and iii) current use of iron-containing supplements. The sample size needed to detect a significant effect of the iron intervention was estimated based on expected changes in serum ferritin concentrations, using an online web calculator (www.powerandsamplesize.com). A change in ferritin of 55 µg/L would raise the lower level (15 µg/L) to 70 µg/L, which is well within the reference area for ferritin (15–200 µg/L) (14). With the following settings: 1) an increase of 55 µg/L in the intervention group, 2) no change in the control group, 3) a standard deviation (SD) of 40 µg/L, 4) 80% power and 5) 5% Type I error rate, the calculated sample size was nine participants per group.

Participants were recruited through information meetings arranged by the two soccer clubs where the project leader presented the study protocol with inclusion and exclusion criteria, formally inviting the players to participate in the study. Those who accepted the invitation and gave written informed consent were randomized in a 1:1 allocation ratio to either an intervention group (n = 12) receiving an oral low-dose iron supplementation (27 mg of elemental iron per day as ferrous bisglycinate, 100 tablets, Nycoplus) or a control group (n = 11) without supplementation. Randomization was performed using simple random allocation: sampling kits were pre-numbered and randomly assigned to one of the two study groups, and each participant selected a random sampling kit as a ‘lottery’. The researcher responsible for generating the allocation sequence had no influence on the assignment of kits to participants. At baseline, participants who were allocated to the intervention group received iron supplements and were instructed to take one tablet daily until the 3-month follow-up visit. No additional or differential care was provided to the two groups, and all participants were instructed to continue their regular diet throughout the study period. The soccer coaches and players were consulted during the planning phase to align data sampling with the club’s training and competition schedule but were not involved in the design of research questions, data analysis or reporting of results.

Participants submitted a digital 7-day food diary at baseline. Blood and fecal samples were collected at two visits: baseline and 3-month follow-up at the Norwegian School of Sport Sciences (Oslo, Norway). In addition, menstrual status, use of contraceptives and medications, and stool consistency were recorded via questionnaires at both visits. While detailed dietary intake was not measured at follow-up, participants were asked about dietary supplement use and potential changes in eating habits from baseline using an open-ended questionnaire. Compliance with the iron intervention was assessed using a questionnaire covering supplement intake frequency (1–7 days/week), reasons for missed doses, whether missed doses were sporadic or continuous, and whether more than one tablet was taken per day. Additionally, plastic vials with unconsumed capsules were collected at follow-up. At the end of the study, participants received personal reports containing information about their iron status and dietary intake, together with individualized nutritional recommendations. Participants with low iron status (flagged as outside the reference limits by the accredited laboratory) were advised to consult their general practitioner for further evaluation and follow-up.

Group allocation was blinded to the soccer coaches and collaborators interpreting the results of the statistical analyses. All participants provided written informed consent prior to participation and were free to withdraw at any time. The study was conducted according to the Declaration of Helsinki. The Regional Committees for Medical and Health Research Ethics in South-East Norway gave ethical approval for this work (ref. no. 100048).

### Dietary assessments

Participants submitted a digital 7-day food diary at the baseline visit for estimation of their habitual dietary intake. Prior to the registration, the participants attended a short presentation with instructions and examples on how to complete the diary. Later, they received a hyperlink to the web-based registration form and were instructed to register all foods and beverages consumed during the registration period. To aid quantification of intakes, they were also encouraged to include pictures of their meals. In the digital registration form, participants recorded the type and amount of each food or beverage using household measures (e.g. glass, cup, handful, spoon), as well as meal type (breakfast, lunch, dinner, snack) and the time and place of consumption. Dietary intake was recorded using household measures instead of weighted amounts in gram to reduce participant burden.

Data from the digital diary were exported to an Excel file where each dietary item was manually assigned a specific food code from the AE-18 version of the food composition database in the food and nutrient calculation system KBS (KBS, AE-18 version 7.3, University of Oslo, Norway). Amounts registered were coded in grams based on standard amounts of the registered household measures and portion sizes. This dataset was then imported into KBS and estimates of daily intakes of energy, nutrients and food groups were calculated. Food diaries with fewer than three registered days were predefined as invalid and would be excluded from analysis. However, all participants recorded their diet for at least three days. Because the digital registration form was developed for the current project and had not been previously validated, its performance was evaluated by analyzing serum carotenoids in relation to reported intake of fruits, berries and vegetables (described further in Supplemental Digital Content 1, **Section 6**, Evaluation of the food diary).

### Assessment of self-reported health-related complaints

Participants reported health-related complaints at baseline and 3-month follow-up using an open-ended question where they were asked if they had been ill, experienced pain or other health problems during the past week. This was included to pick up eventual GI-related problems or other unexpected adverse events that might be related to the iron supplementation.

### Collection and preparation of blood samples

At both study visits, fasting blood samples were collected between 06:00 and 10:00 a.m., prior to soccer training. Venous blood was collected in serum vacuum tubes (5.0 mL w/gel) and EDTA tubes (3.0 mL). Immediately after blood collection, serum tubes were inverted 10 times and incubated at room temperature for 0.5–2 hours to complete the coagulation process, followed by centrifugation at 2,000 × g for 12 minutes at room temperature. From each participant, one serum tube was delivered to an accredited laboratory (Fürst, Furuset, Oslo, Norway) for biomarker analyses and one serum tube was aliquoted into microtubes (2.0 mL). The latter was transported on dry ice to the Norwegian University of Life Sciences (Ås, Norway) and stored at –80°C until further analysis. EDTA tubes were kept at 4°C and analyzed within a few hours after sampling.

### Collection and preparation of fecal samples

Participants received a kit for fecal sample collection which contained instructions, a collection tube, a freezer element and a Bristol Stool Form Scale (BSFS) questionnaire to report stool consistency. Participants were instructed to perform the sampling at home during the same week as the study visits and store the sample in their own freezers immediately after defecation. At each study visit, fecal samples were delivered by the participants in frozen condition using the freezer element. Upon arrival, samples were immediately placed on dry ice and transported to the Norwegian University of Life Sciences (Ås, Norway) for storage at –80°C until aliquotation.

Approximately one day prior to aliquotation, fecal samples were transferred from –80°C to – 20°C. Aliquotation was performed in a laminar flow cabinet, with samples kept on ice throughout the procedure to maintain a semi-frozen condition. From each sample, a small fraction of feces was transferred to 1.5 mL microtubes using sterile disposable scalpels and tweezers. Non-disposable equipment was thoroughly washed between samples with detergent and warm water followed by disinfection with 70% ethanol and DNA decontamination reagent (Cat. No. 43944, Sigma-Aldrich).

Fecal aliquots for enzyme-linked immunosorbent assay (ELISA), SCFA analysis and 16S ribosomal RNA (rRNA) gene sequencing were weighed and logged. Aliquots for 16S rRNA sequencing were added 600 µL Stool Transport and Recovery (S.T.A.R.) buffer (Cat. No. 03335208001, Roche Diagnostics) and vortexed. Negative controls for 16S sequencing were prepared by adding 600 µL S.T.A.R. buffer to 1.5 mL tubes while positive controls were prepared by centrifuging 1.2 mL *E. coli* culture at 15,871 × g for 10 minutes at 20°C, discarding the supernatant and resuspending the resulting pellet in 300 µL S.T.A.R. buffer. Fecal aliquots and control samples were stored at –80°C until further analysis.

### Measurement of iron, inflammation and liver damage biomarkers

In serum samples, ferritin, iron, transferrin saturation, TIBC, ASAT, ALAT, GGT and CRP were measured by an accredited laboratory (Fürst, Furuset, Oslo, Norway). In EDTA plasma, hemoglobin concentration and red blood cell counts were analyzed at the Norwegian Olympic and Paralympic Training Center (Oslo, Norway) using an automated hematology analyzer (pocH-100i, Sysmex, Kobe, Japan). In the current study, iron status was characterized using the Nordic reference intervals for adult females (14) combined with athlete-specific recommendations for ferritin levels (15). Serum ferritin < 35 µg/L was considered indicative of depleted iron stores (stage 1 of iron deficiency) (15) while iron deficiency anemia (stage 3 of iron deficiency) was defined as serum ferritin < 15 µg/L and hemoglobin concentration < 11.7 g/dL (14).

### Measurement of serum NGAL, LBP and hepcidin

Serum concentrations of NGAL, LBP and hepcidin were quantified in duplicates using the Human Lipocalin-2/NGAL DuoSet ELISA (DY1757, R&D Systems), Human LBP DuoSet ELISA (DY870, R&D Systems) and Human Hepcidin DuoSet ELISA (DY8307, R&D Systems), respectively. Assays were performed according to the manufacturer’s instructions with the following modifications: gentle shaking of samples during incubation (300 rpm) to enhance analyte binding, inclusion of extra standard points at the lower end of the curve to improve analyte detection, use of Dulbecco’s phosphate buffered saline (DPBS) without Tween20 for the initial wash step, and overnight plate blocking at 4°C.

Preliminary test experiments (data not shown) were performed for each analyte to determine the most optimal sample dilution that minimized potential inhibitory effects and maximized the probability of concentrations falling within the dynamic range of the standard curves. Based on results from the test experiments, serum samples were diluted 1:200 for NGAL and hepcidin and 1:1000 for LBP using Reagent Diluent (DY995, R&D Systems). Diluted samples were transferred to 96-well storage plates and stored at –20°C until analysis.

To minimize intra-individual variability due to between-plate differences, all samples from the same participant were analyzed on the same assay plate. Samples were distributed across plates to ensure approximately equal representation of study groups per plate. An in-house internal control sample was included on each plate to monitor between-plate variability. Additionally, a negative control, prepared by omitting the capture antibody, was included to assess unspecific binding. Optical density was measured at 450 nm with wavelength correction at 540 nm using a SpectraMax M2 microplate reader (Molecular Devices, USA), and data were acquired using SoftMax Pro software (v.6.5). Analyte concentrations were calculated using four-parameter logistic regression standard curves and are reported as ng/mL for NGAL and hepcidin and as µg/mL for LBP.

### Measurement of serum cytokines

Serum cytokines were quantified using a magnetic bead-based multiplexed immunoassay (Luminex xMAP technology). Serum samples were diluted 1:4, centrifuged at 10,000 × g for 10 minutes at 4°C, transferred to a 96-well storage plate and stored at –20°C until analysis. Interleukin (IL)-2, IL-4, IL-6, IL-8, IL-10, tumor necrosis factor-alpha (TNF-α), granulocyte-macrophage colony-stimulating factor (GM-CSF) and interferon-gamma (IFN-γ) were measured in duplicates, with some exceptions due to limited space. The assay was performed according to the manufacturer’s instructions (Bio-Plex Human Cytokine 8-plex Assay, M50000007A, Bio-Rad Laboratories, Inc., Norway), with the inclusion of an additional standard point at the lower end of the curve to improve sensitivity and detection of low-concentration analytes. According to the product datasheet, typical intra- and inter-assay coefficients of variation (CV) were ≤ 15% and ≤ 25%, respectively.

Data acquisition was performed using a Bio-Plex 200 System equipped with high-throughput fluidics. Cytokine concentrations were calculated using five-parameter logistic regression standard curves and are reported as pg/mL. Several samples fell below the lower limit of quantification, some of which were automatically extrapolated by the software (see **Supplementary Figures S2 and S3**, Supplemental Digital Content 1, for an overview of sample detection in the total study population and by group, respectively). IL-2, IL-4, IL-10 and GM-CSF were excluded from downstream analyses due to a high percentage of samples (> 80%) falling below the detection limit.

### Measurement of fecal NGAL

Fecal aliquots for quantification of NGAL (also known as lipocalin-2) were reconstituted as described by Chassaing et al. (32) with some modifications. Feces (50–100 mg) were mixed with DPBS containing 0.1% Tween20 (Cat. No. P1379, Sigma-Aldrich) at a ratio of 100 mg feces per 1 mL buffer. Samples were homogenized for 20 minutes using a mixer mill (MM2, Retsch) at speed setting 60, followed by centrifugation at 13,523 × g for 10 minutes at 4°C. If the supernatant was not clear after centrifugation, a second centrifugation was performed at 15,871 × g for 10 minutes. Clear supernatants were collected and stored at –20°C until analysis.

Fecal NGAL concentrations were determined using the same ELISA protocol described for serum NGAL (see “Measurement of serum NGAL, LBP and hepcidin” section) with a 1:10 dilution in DPBS. Final concentrations were corrected for wet fecal weight and are reported as ng/g feces.

### Measurement of fecal SCFA

Fecal SCFA concentrations were determined by vacuum distillation followed by gas chromatography, described further in Supplemental Digital Content 1 (**Section 3**, SCFA analysis). The measured SCFA were acetic, propionic, butyric, iso-butyric, valeric, iso-valeric, caproic and iso-caproic acid. Concentrations were corrected for wet fecal weight and are reported as mmol/kg feces. The branched-chain SCFA (iso-butyric, iso-valeric and iso-caproic acid) were analyzed collectively by summing their concentrations as they represent only a small fraction of the total SCFA, particularly iso-caproic acid which had low detectability. None of the iso-SCFA reached statistical significance when analyzed individually (data not shown).

### Illumina 16S rRNA gene sequencing

Microbial diversity and composition in fecal samples were determined by targeted sequencing of the V3–V4 hypervariable region of the 16S rRNA gene. The procedure is described in detail in Supplemental Digital Content 1 (**Section 4.1**, Library preparation).

### Processing of sequencing data and taxa filtering

Processing of raw Illumina sequencing reads is described in detail in Supplemental Digital Content 1 (**Section 4.2**, Sequence processing). For the per protocol analyses (see “Statistical analysis” section), a total of 1,282 OTUs were identified. Across the 36 samples, the total read number was 1,640,176, with a mean (SD) and median (Q1, Q3) of 45,560 (18,134) and 45,176 (36,594, 51,512) reads per sample, respectively. All samples exceeded 10,000 reads. Library sizes by study group are shown in **Supplementary Figure S6** and read counts for control samples are shown in **Supplementary Figure S7** (Supplemental Digital Content 1). To reduce the disturbance of low-abundance taxa and improve suitability of the data for statistical analysis, taxa with a median relative abundance below 0.01% across both visits in at least one study group were excluded from the differential abundance and correlation analyses. Given the median library size of approximately 45,000 reads per sample, this threshold corresponds to taxa represented by fewer than ∼5 reads per sample, on average. After filtering, 171 taxa were retained for analysis, comprising 11 phyla, 11 classes, 20 orders, 36 families and 93 genera. Summary statistics for the included taxa are provided in **Supplementary Table S9** (Supplemental Digital Content 1).

### Statistical analysis

Continuous variables are presented as mean with SD or median with lower and upper quartiles (Q1, Q3) and categorical variables as frequencies with percentages. Due to lack of compliance in three of the 12 participants in the intervention group, a per protocol analysis including only those who adhered to the intervention was used to assess differences between groups. This approach was selected as the primary strategy given the small sample size, with the aim of evaluating the effect of actually receiving the treatment rather than merely being assigned to it (33). However, the full dataset (n = 23) was used for assessment of baseline dietary intakes and iron status. Because data were collected at only two time points, missing values were not imputed. None of the participants included in the per protocol analysis had missing data.

To investigate between-group differences at the 3-month follow-up, we used multiple linear regression, specifically an analysis of covariance (ANCOVA), with the baseline value of the outcome included as a covariate (model: follow-up ∼ baseline + group). This approach is recommended in randomized trials with one follow-up measurement as it properly adjusts for possible baseline differences compared to analysis of change scores (34). For outcomes where a significant baseline × group interaction was detected (*P* < 0.05), the model was extended to include the interaction term (extended model: follow-up ∼ baseline + group + baseline × group). In these cases, both the main effect of group (β_group_) with corresponding *P*-value (*P*_group_) and the interaction term (β_interaction_, *P*_interaction_) were reported with 95% confidence intervals (CI). Given the small sample size, within-group changes from baseline to follow-up were also investigated regardless of between-group effects using paired samples t-tests (model: variable ∼ time). Categorical variables were compared using Fisher’s exact test, which is suitable for small sample sizes and low expected cell counts. In exploratory analyses, potential associations between continuous variables were examined using Pearson’s or Spearman’s rank correlation, depending on the data distribution.

Prior to statistical analysis, model assumptions were thoroughly evaluated. Potential outliers were identified using influence diagnostics, including leverage, standardized and studentized residuals, Cook’s distance, DFFIT and DFBETAS. If an outlier could not be attributed to data entry or measurement error and had a large influence on the model, analyses were conducted both with and without the observation(s). When exclusion did not alter the overall conclusion, results from the full dataset were reported. For variables that deviated from normality, non-parametric tests or analyses of log-transformed data were performed. If results remained consistent, untransformed data were reported for clarity and ease of interpretation.

Serum cytokines that fell below the limit of detection and were not automatically extrapolated by the analysis software were imputed using maximum likelihood estimation (*genasis package*, v.1.0). This approach works well for left-censored data (35). Imputation was performed for IL-6 and IFN-γ (see **Supplementary Table S4**, Supplemental Digital Content 1, for an overview of imputed values).

Statistical analyses and data visualization were performed using R (v.4.5.1) and RStudio (v.2025.05.1 Build 513). The significance level was set to 0.05. As the trial was not powered for secondary outcomes, and many outcome variables were highly intercorrelated due to overlapping physiological mechanisms (e.g. iron-related biomarkers or inflammatory biomarkers), no correction for multiple comparisons was applied (36, 37). One exception was the differential abundance analysis (see “Microbiota data analysis” section) in which the Benjamini–Hochberg method was applied to control the false discovery rate (FDR).

### Microbiota data analysis

Alpha diversity, also known as within-sample diversity, was assessed using the Shannon and Inverse Simpson indices (*vegan package*, v.2.6-10). Between-group differences at 3-month follow-up were investigated using ANCOVA with baseline adjustment and within-group changes were assessed using paired samples t-tests, as described above. Beta diversity, a measure of between-sample diversity that quantifies the (dis)similarities, or ‘distances’, between two samples, was assessed using principal coordinate analysis (PCoA) (*ape package*, v.5.8-1) based on Bray-Curtis dissimilarities (*phyloseq package*, v.1.48.0). The Bray-Curtis dissimilarity ranges from 0 to 1, where higher values indicate greater dissimilarity (less similar composition) and lower values indicate more similar community composition. A scree plot of the first 10 principal coordinate axes is shown in **Supplementary Figure S8** (Supplemental Digital Content 1). Non-metric multidimensional scaling was also performed but produced similar results and is therefore not reported.

To test whether changes in overall microbiota composition from baseline to 3-month follow-up differed between groups, we used permutational multivariate analysis of variance (PERMANOVA) (model: distance matrix ∼ group × visit) with 1,000 permutations and adjustment for repeated measures in the permutation design (*vegan package*, v.2.6-10; *permute package*, v.0.9-7). Additional PERMANOVAs stratified by visit (baseline, follow-up) and by group (control, intervention) were performed to assess between-group differences at each visit (model: distance matrix ∼ group) and within-group changes over time (model: distance matrix ∼ visit). PCoA and centroid calculations were based on the full dataset to ensure consistent comparison of community structure across groups and visits. Average distances to centroids were used to assess the homogeneity of group dispersions (*vegan package*, v.2.6-10).

Between-group differences in taxa relative abundances were investigated at each taxonomic rank (phylum, class, order, family, genus) using ANCOVA with baseline adjustment, as previously described. Analyses were conducted on both untransformed relative abundances (%) and square-root–transformed values to address the typical non-normality and heteroscedasticity of microbiota compositional data. Within-group changes in relative abundances (%) were investigated using paired samples t-tests (for square-root-transformed values) or Wilcoxon signed-rank tests (for untransformed values).

## RESULTS

### Study population

A flow diagram of the study participants is presented in **Supplementary Figure S18** (Supplemental Digital Content 1). A total of 26 female tier 3 soccer players accepted the invitation to participate in the study but three withdrew prior to baseline, leaving 23 participants (control, n = 11; intervention, n = 12) completing the baseline visit. Subsequently, two participants in the control group withdrew prior to the follow-up visit because they transferred to other soccer clubs, resulting in 21 participants completing the trial. Due to lack of compliance with the iron intervention in three of the participants in the intervention group, a total of nine participants per group were included in the per protocol analysis. The reasons for lack of compliance were medical reasons (n = 1) and forgetfulness (n = 2).

Baseline characteristics are presented in **Table 1** for all participants (n = 23) and separately for the control (n = 9) and intervention (n = 9) groups included in the per protocol analysis. The median (Q1, Q3) age of the participants was 21 (20, 23) years and they had a mean (SD) body mass index (BMI) of 22.3 (1.4) kg/m^2^, which is within the normal BMI range. Reported health complaints during the week before the baseline visit were muscle stiffness (n = 1), bone stress injury (n = 1) and migraine (n = 1), and overall medication use was low. Two participants reported not having menstruation (using intrauterine devices) and one reported having skipped the last menstruation (using oral contraceptives). Baseline characteristics presented separately for each group indicate that randomization achieved comparable groups, apart from some menstrual cycle variations.

**Table 1.** Baseline characteristics for all participants (n = 23) and separately for the control (n = 9) and intervention (n = 9) group included in the per protocol analyses.

| Characteristics | All (n = 23) | Per protocol analysis |  |
| --- | --- | --- | --- |
|  |  | Control (n = 9) | Intervention (n = 9) |
| Age (y) | 21 (20, 23) | 20 (20, 23) | 20 (20, 22) |
| Weight (kg) | 64.3 (5.7) | 65.6 (7.4) | 65.2 (3.3) |
| Height (cm) | 169.6 (5.3) | 169.6 (5.4) | 171.6 (4.0) |
| BMI (kg/m<sup>2</sup>) | 22.3 (1.4) | 22.8 (1.7) | 22.2 (1.3) |
| Health-related complaints<sup>1</sup>, n (%) | 3 (13) | 1 (11) | 2 (22) |
| Medicines<sup>2</sup>, n (%) | 7 (30) | 2 (22) | 4 (44) |
| Allergy | 4 (17) | 2 (22) | 1 (11) |
| Asthma | 3 (13) | 1 (11) | 2 (22) |
| Antibiotics | 1 (4) | 0 (0) | 1 (11) |
| Lactic acid tablets | 1 (4) | 1 (11) | 0 (0) |
| Other | 3 (13) | 1 (11) | 2 (22) |
| Contraceptives, n (%) | 10 (43) | 4 (44) | 4 (44) |
| Oral contraceptive pill | 4 (17) | 2 (22) | 0 (0) |
| Subdermal implant | 1 (4) | 0 (0) | 1 (11) |
| Hormonal intrauterine device | 4 (17) | 2 (22) | 2 (22) |
| Copper intrauterine device | 1 (4) | 0 (0) | 1 (11) |
| Menstrual cycle |  |  |  |
| Days since start of last bleeding | 13.5 (6.0, 37.3) | 15.5 (13.3, 36.5) | 6.0 (5.0, 14.0) |
| Duration of last bleeding (days) | 5.0 (4.8, 6.0) | 4.5 (4.0, 5.0) | 6.0 (5.0, 7.0) |
| No menstruation, n (%) | 3 (13) | 3 (33) | 0 (0) |

Values are mean (SD) or median (Q1, Q3) for continuous variables and n (%) for categorical variables.
Abbreviations: BMI, body mass index.
<sup>1</sup> Assessed using an open-ended question asking participants whether they had been ill, experienced pain or other health problems during the past week.
<sup>2</sup> Participants that reported multiple types of medication are counted once in each applicable subcategory and thus totals may exceed the number of unique individuals.

### Dietary intake at baseline

Participants recorded their food intake for a mean (SD) of 6.3 (1.3) days. Most participants (74%) completed the full 7-day diary, while 2 (9%) recorded 6 days, 1 (4%) recorded 5 days, 1 (4%) recorded 4 days and 2 (9%) recorded 3 days. The mean (SD) and median (Q1, Q3) total daily energy intake were 10,270 (1,904) kJ and 9,931 (9,139, 11,273) kJ, respectively. Baseline intake of iron and other selected micronutrients, expressed as percentages of the RI from NNR-2023 (5), is shown in **Supplementary Figure S10** (Supplemental Digital Content 1), while **Figures 1**A and 1B display iron contributions from selected food groups and dietary iron intake as a box plot with individual values, respectively. The mean (SD) and median (Q1, Q3) daily iron intake were 12.1 (3.7) and 11.2 (10.3, 12.6) mg/day, corresponding to 80 and 75% of the RI for the Norwegian population (5), respectively. Overall, 21 (91%) participants had iron intakes below the recommended level of 15 mg/day. In comparison, when applying the U.S. Recommended Dietary Allowance (RDA) for females (18 mg/day) (7), this proportion increased to 96%. However, while only 2 (9%) participants met the RI, having an iron intake between 15 and 60 mg/day (the tolerable upper intake level), all except 1 (96%) had intakes above the AR for the Norwegian population (5).

**Figure 1.**
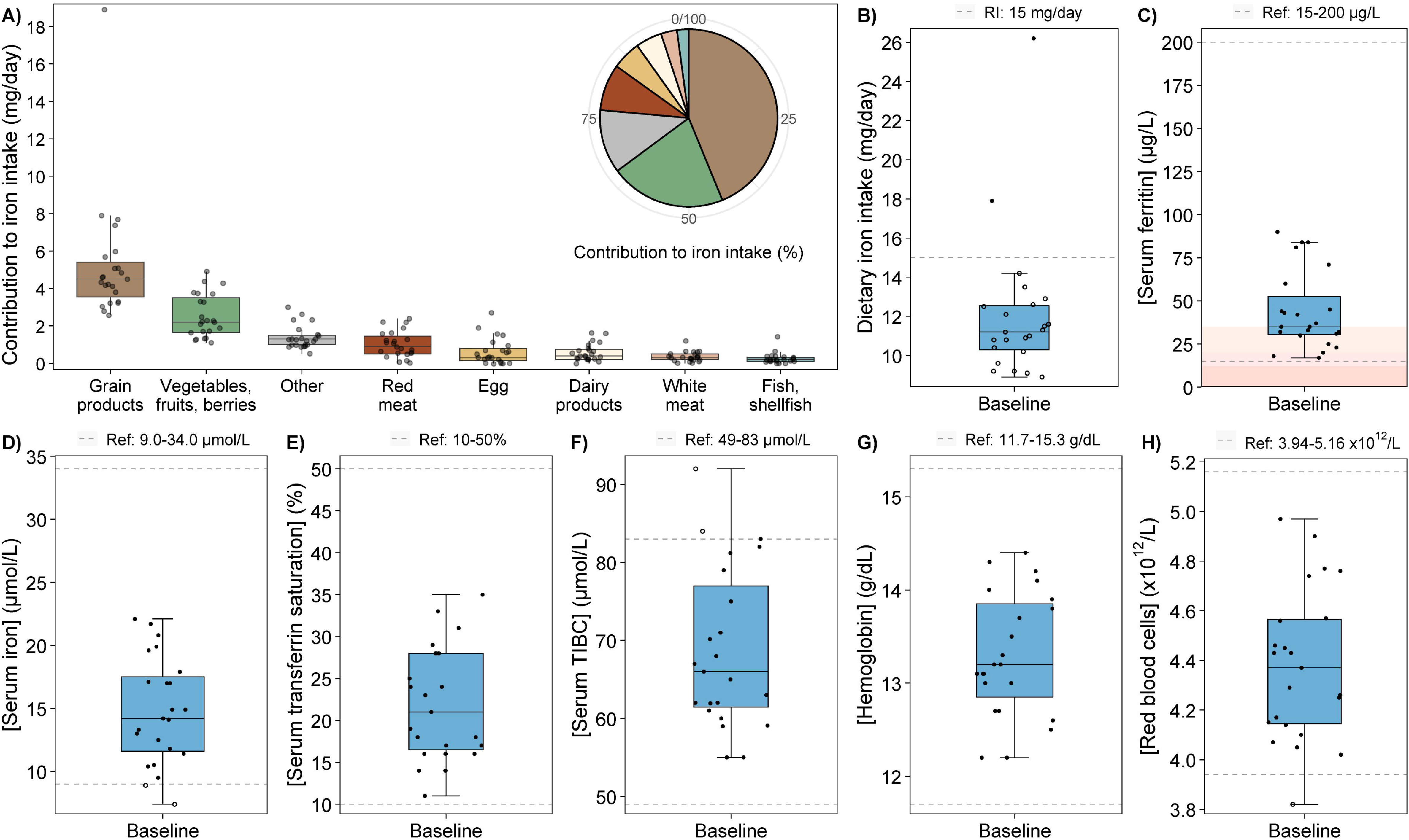
Dietary iron intake and iron status biomarkers at baseline (n = 23). Each dot represents one participant. **(A)** Daily dietary iron intake (mg/day) from selected food groups with percentage contributions shown in the upper right corner (brown, grain products; green, vegetables, fruits and berries; grey, other; red, red meat; yellow, egg; white, dairy products; pink, white meat; blue, fish and shellfish). The “Other” category includes contributions from ungrouped foods, cakes, potatoes, butter, margarine, oil, sugar and sweets, beverages, mixed dishes and spices/salt. **(B–H)** Daily dietary iron intake and iron-related biomarkers with RI or reference values indicated by horizontal dashed lines. Non-filled circles denote observations outside the RI/Ref. (B) Daily dietary iron intake, **(C)** serum ferritin with colored background to indicate the three stages of iron deficiency (iron depletion, < 35 µg/L; iron-deficient erythropoiesis, < 20 µg/L; iron-deficient anemia, < 12 µg/L), **(D)** serum iron, **(E)** serum transferrin saturation, **(F)** serum TIBC, **(G)** hemoglobin concentration and **(H)** red blood cell counts. Ref, reference area; RI, recommended intake; TIBC, total iron-binding capacity.

Daily iron intake was strongly and positively correlated with total daily energy intake (Pearson’s correlation coefficient, *r_p_* = 0.88, *P* < 0.001) (**Supplementary Figure S11**, Supplemental Digital Content 1). To identify which dietary food groups contributed most to iron intake in the current study population, we calculated the amount of iron derived from each food. The estimated daily intakes of selected food groups are presented in **Supplementary Table S12**, and their contribution to daily iron intake is shown in Figure 1A (see **Supplementary Figure S13**, Supplemental Digital Content 1, for percentage contributions for each participant). The largest iron contribution was from grain products (44%), followed by vegetables, fruits and berries (21%), red meat (8%), egg (5%), dairy products (5%), white meat (3%), and fish and shellfish (2%). Other food groups (ungrouped foods, cakes, potatoes, butter, margarine, oil, sweets, beverages, mixed dishes, spices/salt) collectively accounted for 12% of the daily iron intake.

In the evaluation of the dietary assessment method, we found that daily intake of vegetables, fruits and berries correlated positively with serum carotenoids (see **Supplementary Table S14** for correlation coefficients and **Figures S15–S17** for scatterplots, Supplemental Digital Content 1), indicating good performance of the web-based food diary in estimating the intakes of these food groups.

### Iron status at baseline

Baseline iron status of all participants (n = 23) is shown in Figure 1C–H and presented in **Table 2**. The median (Q1, Q3) serum ferritin concentration (Figure 1C) was 35 (30.5, 52.5) µg/L, with levels ranging from 17 to 90 µg/L. While most participants (57%) had adequate iron stores (ferritin ≥ 35 µg/L) according to suggested cut-off values for athletes (15) (pink area in Figure 1C), 10 (43%) participants had ferritin levels indicative of depleted iron stores (ferritin < 35 µg/L) of which 2 (9%) had levels indicating stage 2 iron-deficient erythropoiesis (ferritin < 20 µg/L). Serum iron (Figure 1D) and transferrin saturation (Figure 1E) were mostly within the respective reference areas (dashed lines in Figures 1D and E) except for 2 (9%) participants having serum iron below the recommended area. Additionally, 2 (9%) participants displayed an above normal serum TIBC (Figure 1F). Hemoglobin concentrations (Figure 1G) and red blood cells (Figure 1H) were normal, except for 1 (4%) participant with red blood cell levels below the reference area.

**Table 2.** Serum biomarkers at baseline and 3-month follow-up with estimated between-group differences at follow-up.

|  | Control (n = 9) |  | Intervention (n = 9) |  | Per protocol ANCOVA |  |
| --- | --- | --- | --- | --- | --- | --- |
| | Baseline | 3 months | Baseline | 3 months | $\beta_{\text{group}}$ | $P_{\text{group}}$ |
| **Iron status** |  |  |  |  |  |  |
| Ferritin ($\mu\text{g/L}$) | 43.4 (27.2) | 40.9 (20.8) | 35.2 (8.0) | 44.4 (16.3) | 9.1 [−4.7, 22.9] | 0.18 |
| Iron<sup>1</sup> ($\mu\text{mol/L}$) | 15.9 (4.8) | 17.3 (5.6) | 12.9 (4.1) | 20.3 (6.0) | 6.5 [0.1, 13.0] | **0.05** |
| Transferrin saturation (%) | 24.1 (7.7) | 26.9 (10.0) | 18.6 (4.6) | 30.6 (7.9) | 46.9 [20.6, 73.2]<br>−2.0 [−3.3, −0.8]<sup>§</sup> | **0.002**<br>**0.004<sup>§</sup>** |
| TIBC ($\mu\text{mol/L}$) | 67.6 (12.6) | 66.7 (14.8) | 69.8 (8.8) | 66.4 (6.7) | −2.4 [−6.9, 2.1] | 0.28 |
| Hemoglobin (g/dL) | 13.1 (0.8) | 13.3 (0.9) | 13.5 (0.5) | 13.5 (0.5) | −0.2 [−0.7, 0.3] | 0.42 |
| Red blood cells ($\times 10^{12}/\text{L}$) | 4.3 (0.3) | 4.4 (0.3) | 4.5 (0.3) | 4.5 (0.2) | −0.1 [−0.2, 0.1] | 0.47 |
| Hepcidin (ng/mL) | 13.0 (9.6) | 16.9 (7.4) | 10.7 (4.3) | 13.2 (5.1) | −2.7 [−8.6, 3.1] | 0.34 |
| **Liver damage** |  |  |  |  |  |  |
| ASAT (U/L) | 22.4 (4.6) | 19.0 (4.9) | 26.9 (11.2) | 21.7 (9.7) | −0.7 [−5.1, 3.6] | 0.73 |
| ALAT (U/L) | 20.8 (4.0) | 20.4 (6.0) | 23.8 (7.5) | 20.3 (7.3) | −2.7 [−7.2, 1.8] | 0.22 |
| GGT (U/L) | 16.1 (4.9) | 15.9 (3.8) | 14.1 (4.3) | 13.8 (3.8) | −1.0 [−3.9, 2.0] | 0.49 |
| **Inflammation** |  |  |  |  |  |  |
| CRP (mg/L) | 1.7 (1.9) | 1.4 (1.5) | 0.7 (0.9) | 0.6 (0.7) | −0.04 [−0.3, 0.2] | 0.68 |
| IL-6<sup>2</sup> (pg/mL) | 0.3 (0.3) | 0.4 (0.4) | 0.3 (0.4) | 0.2 (0.3) | −0.5 [−0.9, −0.03] | **0.04** |
| IL-8 (pg/mL) | 4.3 (1.8) | 3.8 (2.3) | 3.8 (1.7) | 3.3 (1.9) | −0.07 [−1.3, 1.2] | 0.90 |
| TNF-α (pg/mL) | 6.2 (2.9) | 4.9 (3.3) | 8.7 (9.5) | 6.8 (8.2) | −0.2 [−1.6, 1.2] | 0.72 |
| IFN-γ (pg/mL) | 0.3 (0.2) | 0.3 (0.2) | 0.8 (1.8) | 1.0 (2.0) | 0.2 [−0.1, 0.4] | 0.20 |
| NGAL (ng/mL) | 84.9 (14.6) | 82.0 (14.8) | 108.9 (34.9) | 96.3 (25.5) | 7.0 [−15.1, 29.1] | 0.51 |
| LBP ($\mu\text{g/mL}$) | 7.5 (4.2) | 7.8 (5.5) | 8.1 (2.3) | 9.1 (3.0) | 0.9 [−2.9, 4.6] | 0.62 |

Values are mean (SD). $\beta$ estimates with 95% CI and corresponding $P$ -values are derived from per protocol ANCOVA adjusted for baseline values of the outcome. The $\beta$ estimate represents the between-group difference at follow-up with a positive estimate indicating a higher value in the intervention group compared to the control group. Statistically significant $P$ -values are shown in bold ( $P < 0.05$ ).
Abbreviations: ALAT, alanine aminotransferase; ANCOVA, analysis of covariance; ASAT, aspartate aminotransferase; CRP, C-reactive protein; GGT, gamma-glutamyl transferase; IFN-γ , interferon-gamma; IL, interleukin; LBP, lipopolysaccharide-binding protein; NGAL, neutrophil gelatinase-associated lipocalin; TIBC, total iron-binding capacity; TNF-α , tumor necrosis factor-alpha.
<sup>1</sup> One participant in the intervention group was excluded from the analysis (when included, $P_{\text{group}} = 0.17$ ).
<sup>2</sup> Data were log-transformed prior to analysis, reported $\beta$ estimate and $P$ -value are based on transformed data.
<sup>§</sup> Estimate for the baseline × group interaction ( $\beta_{\text{interaction}}$ ) with corresponding $P$ -value ( $P_{\text{interaction}}$ ) derived from the extended ANCOVA model (follow-up ∼ baseline + group + baseline × group).

### Effects of low-dose iron supplementation on iron status

The effects of iron supplementation on iron-related biomarkers were assessed using ANCOVA with baseline adjustment. Results are presented in Table 2 as β estimates with 95% CI for between-group comparisons and visualized in **Figure 2**. Within-group changes are reported in **Supplementary Table S19** (Supplemental Digital Content 1).

**Figure 2.**
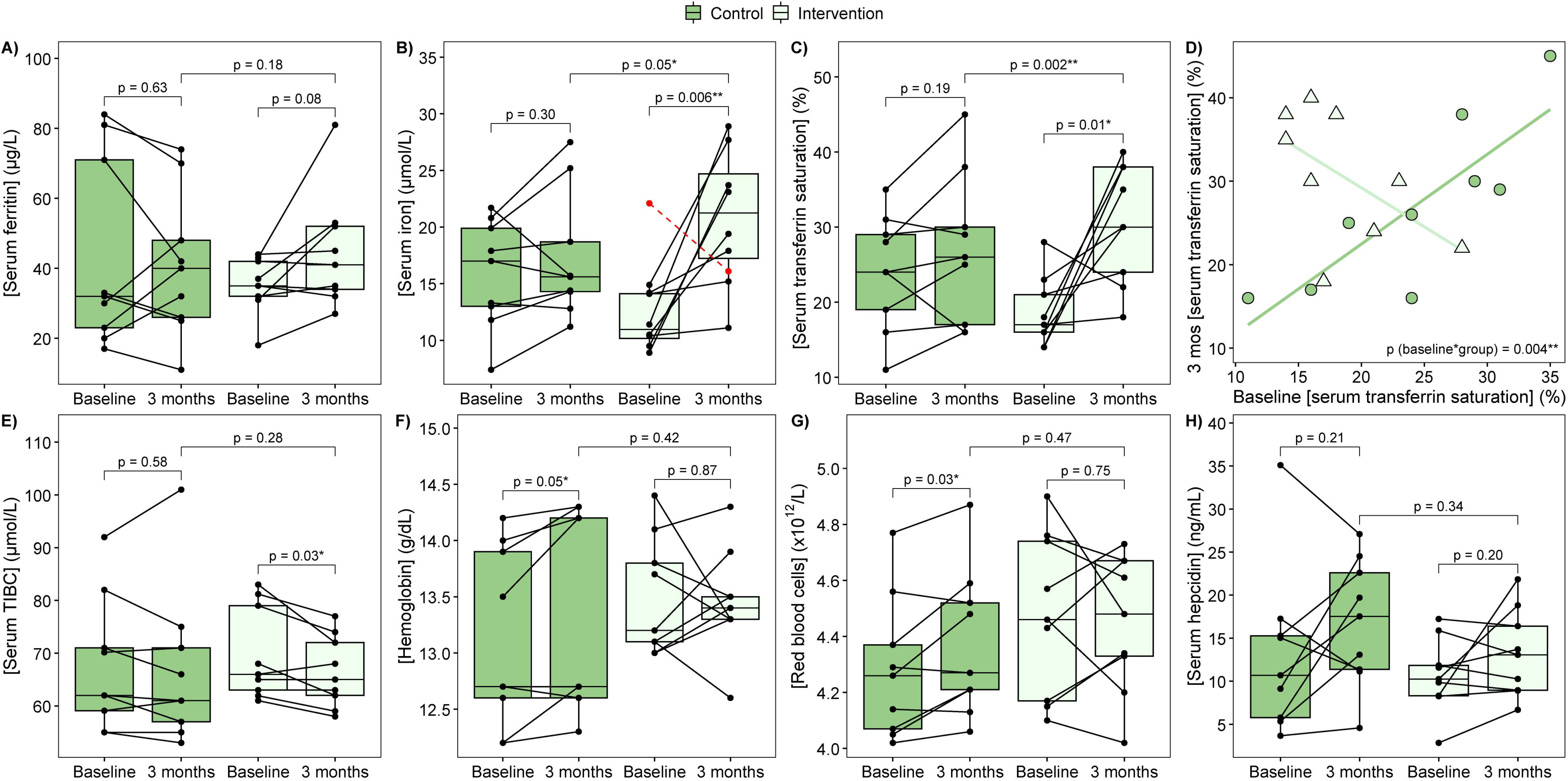
Effects of low-dose iron supplementation on iron status biomarkers. **(A)** Serum ferritin, **(B)** serum iron (participant marked in red was excluded from the statistical analysis), **(C–D)** serum transferrin saturation (significant baseline × group interaction: *P*_group_ reported in C, *P*_interaction_ reported in D), **(E)** serum TIBC, **(F)** hemoglobin concentration, **(G)** red blood cell counts and **(H)** serum hepcidin. Each dot represents one sample, with lines connecting samples from the same participant. The *P*-values are from per protocol ANCOVA with baseline adjustment (between-group comparison) and paired samples t-tests (within-group comparison). Mos, months; TIBC, total iron-binding capacity.

At 3-month follow-up, we found significantly higher serum iron concentrations (Figure 2B) in the intervention group compared to the control group (*P*_group_ = 0.05). Additionally, the within-group analysis showed a significant increase in the intervention group from baseline to follow-up (*P* = 0.006), while no change was found in the control group (*P* = 0.30). Of note, one participant in the intervention group showed a marked decrease in serum iron from baseline to follow-up, clearly deviating from the otherwise consistent increases observed in the group. When this participant was included, the between-group difference was no longer significant (*P*_group_ = 0.17), although the within-group increase in the intervention group remained significant (*P* = 0.02).

Serum ferritin concentrations (Figure 2A) did not differ significantly between the groups at 3-month follow-up (*P*_group_ = 0.18), but the data suggested a non-significant increase in the intervention group (*P* = 0.08). Given this trend for increase and the higher serum iron concentrations in the intervention group at follow-up, we conducted an exploratory post hoc analysis to investigate whether the proportion of participants experiencing declines in serum ferritin from baseline to follow-up was lower in the intervention group. This analysis showed that a higher proportion of control participants had a decline in serum ferritin compared with the intervention group when defining declines as either > 1, ≥ 2, ≥ 3 µg/L (all *P* = 0.05), ≥ 4, ≥ 5, ≥ 6 µg/L (all *P* = 0.009) or ≥ 7 µg/L (*P* = 0.03). As an example, **Supplementary Figure S21** (Supplemental Digital Content 1) shows the proportion of participants in each group with or without a decline in serum ferritin of ≥ 5 µg/L. It should be noted that the difference in proportions between groups did not remain statistically significant for declines ≥ 8 µg/L (*P* > 0.05, data not shown).

For serum transferrin saturation (Figure 2C), we found a significant group effect (*P*_group_ = 0.002) and a significant interaction between baseline values and group (*P*_interaction_ = 0.004), indicating that participants with lower transferrin saturation at baseline experienced greater increases during the iron intervention than participants with higher baseline transferrin saturation (Figure 2D). In addition, transferrin saturation increased significantly from baseline to 3-month follow-up in the intervention group (*P* = 0.01) but not in controls (*P* = 0.19). Serum TIBC (Figure 2E) did not differ significantly between groups at follow-up, but a significant within-group decrease was observed in the intervention group only (*P* = 0.03). Lastly, no significant between-group differences were found for hemoglobin concentration (Figure 2F), red blood cells (Figure 2G) or serum hepcidin (Figure 2H).

### Effects of low-dose iron supplementation on systemic inflammation and liver damage

The effects of iron supplementation on biomarkers of inflammation and liver damage were assessed using ANCOVA with baseline adjustment. Results are presented in Table 2 as β estimates with 95% CI for between-group comparisons and visualized in **Figure 3**. Within-group changes are reported in Supplementary Table S19 (Supplemental Digital Content 1).

**Figure 3.**
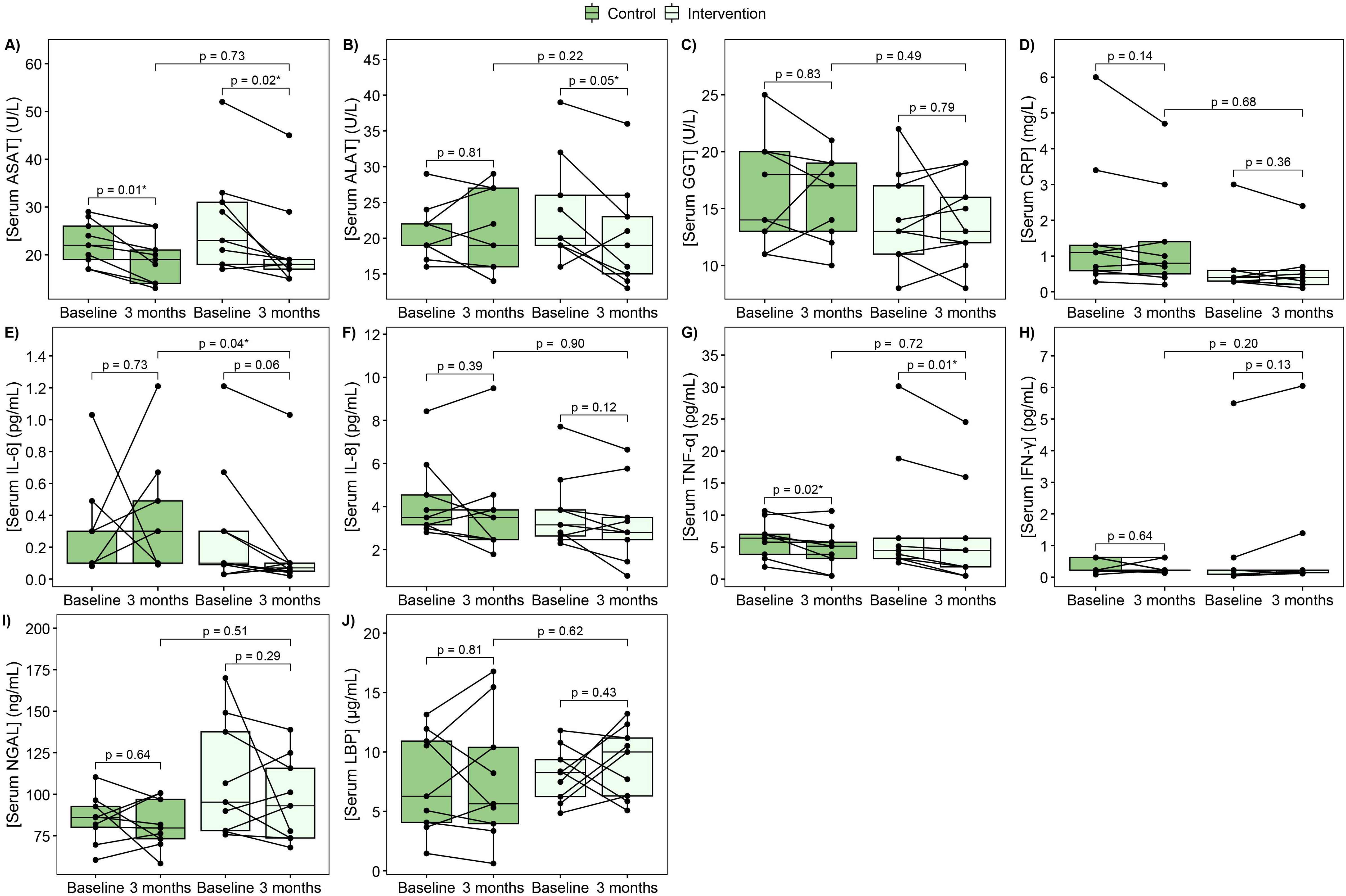
Effects of low-dose iron supplementation on liver damage and inflammatory biomarkers. **(A)** Serum ASAT, **(B)** serum ALAT, **(C)** serum GGT, **(D)** serum CRP, **(E)** serum IL-6 (data were log-transformed prior to analysis but untransformed values are plotted), **(F)** serum IL-8, **(G)** serum TNF-α, **(H)** serum IFN-γ, **(I)** serum NGAL and **(J)** serum LBP. Each dot represents one sample, with lines connecting samples from the same participant. The *P*-values are from per protocol ANCOVA with baseline adjustment (between-group comparison) and paired samples t-tests (within-group comparison). ALAT, alanine aminotransferase; ASAT, aspartate aminotransferase; CRP, C-reactive protein; GGT, gamma-glutamyl transferase; IFN-γ, interferon-gamma; IL, interleukin; LBP, lipopolysaccharide-binding protein; NGAL, neutrophil gelatinase-associated lipocalin; TNF-α, tumor necrosis factor-alpha.

Of all the markers, only serum IL-6 (Figure 3E) differed significantly between the groups at 3-month follow-up, where the intervention group had lower levels than the control group (*P*_group_ = 0.04). For the remaining serum inflammatory markers (CRP, IL-8, TNF-α, IFN-γ, NGAL, LBP; Figures 3D and 3F–J) and the liver damage biomarkers (ASAT, ALAT, GGT; Figures 3A–C), no significant between-group differences were observed. However, serum ASAT decreased significantly from baseline to follow-up in both groups (control, *P* = 0.01; intervention, *P* = 0.02) and serum ALAT decreased significantly in the intervention group only (*P* = 0.05).

### Health-related complaints at follow-up

Apart from one participant reporting posterior knee inflammation and one reporting ankle pain, none of the participants reported any other health-related complaints, including GI-related issues, at 3-month follow-up. However, one participant contacted the study center several months after the study had ended, to report GI-related symptoms (constipation). It is uncertain whether these symptoms were a delayed effect of the iron intervention or occurred independently of the study. We also examined differences between groups in reported BSFS type at follow-up, adjusting for baseline values, and found no significant differences (*P*_group_ = 0.56) (**Supplementary Figure S22**, Supplemental Digital Content 1).

### Effects of low-dose iron supplementation on gut microbiota, microbial metabolites and gut inflammation

The effects of iron supplementation on gut microbiota (diversity and taxa relative abundances) and fecal biomarkers are presented in **Table 3** as β estimates with 95% CI for between-group comparisons and visualized in **Figures 4 and 5**, and **Supplementary Figures S25 and S27** (Supplemental Digital Content 1). Within-group changes are reported in **Supplementary Table S20** (Supplemental Digital Content 1).

**Figure 4.**
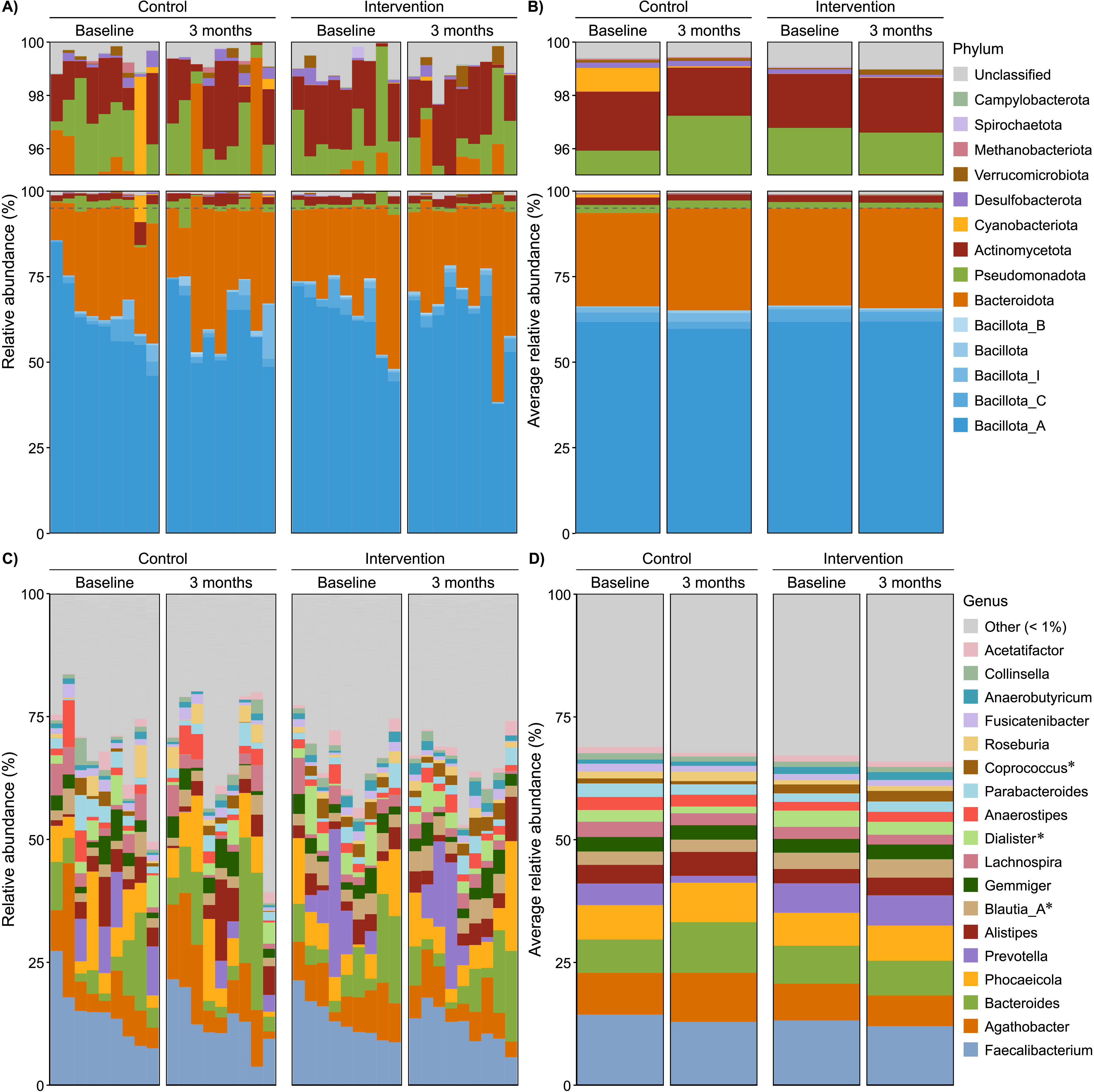
Fecal relative abundances (%) of detected (A, B) phyla and (C, D) genera at baseline and 3-month follow-up. **(A, C)** Each bar represents one participant. Within groups, bars are sorted in descending order according to the most abundant taxon at baseline (phylum, *Bacillota_A*; genus, *Faecalibacterium*), and this order is preserved at follow-up. **(B, D)** Group means are displayed as average relative abundances at each visit. At phylum level, the upper section of the plot (indicated by a dashed horizontal line) is enlarged to improve visualization of less abundant phyla. OTUs with taxonomy confidence < 0.8 are shown as “Unclassified”. Genera with mean relative abundance < 1% are shown as “Other.” Taxa are ordered by overall mean abundance, except “Unclassified” and “Other” which appear at the top regardless of abundance. Taxa that were significantly different between groups at follow-up (per protocol ANCOVA with baseline adjustment; uncorrected for multiple testing) are marked with an asterisk (*).

**Figure 5.**
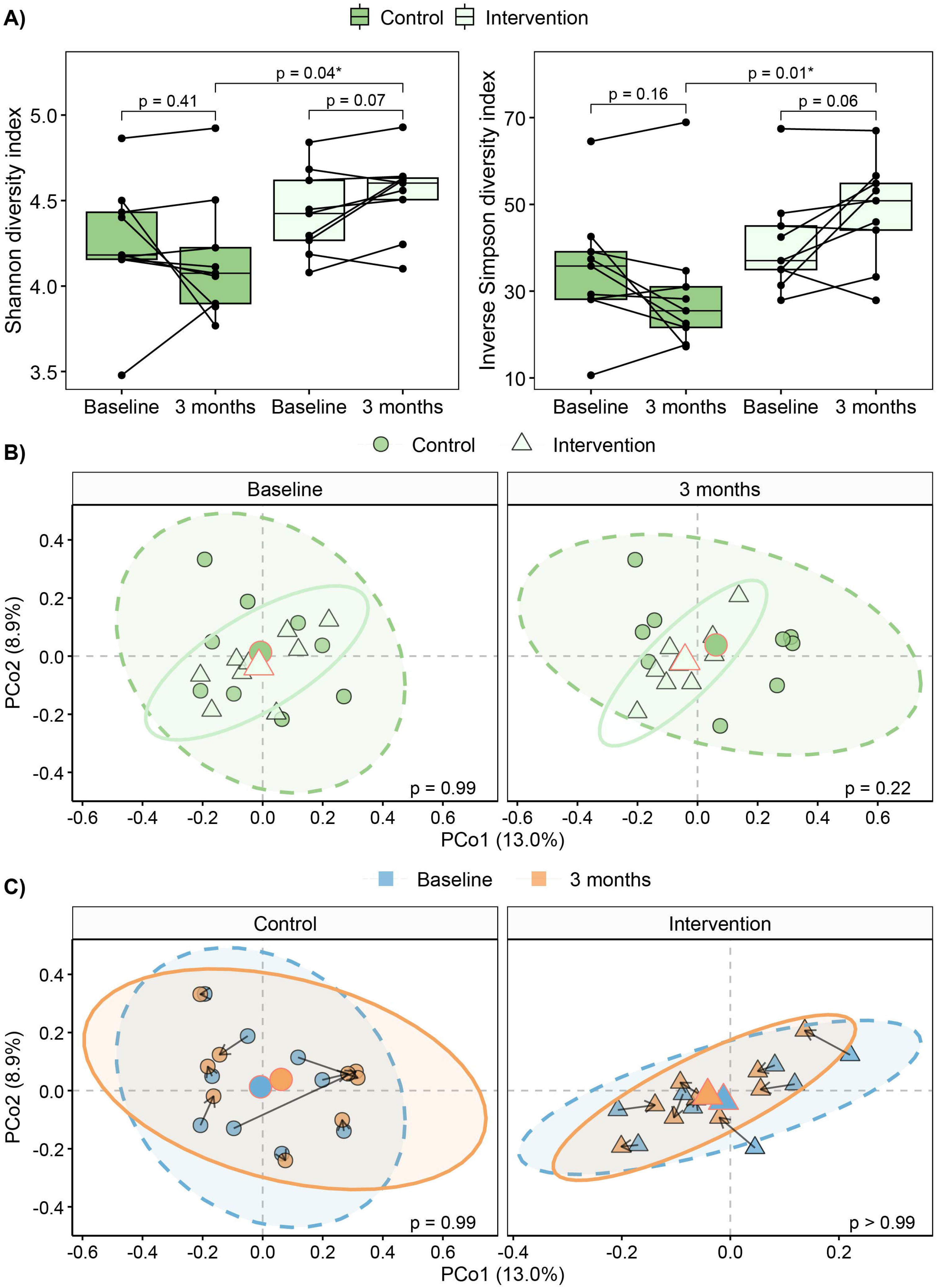
Effects of low-dose iron supplementation on gut microbial diversity. **(A)** Alpha diversity (left, Shannon; right, Inverse Simpson). Each dot represents one sample, with lines connecting samples from the same participant. The *P*-values are from per protocol ANCOVA with baseline adjustment (between-group comparison) and paired samples t-tests (within-group comparisons). **(B)** Beta diversity (PCoA based on Bray-Curtis dissimilarity) at baseline and 3-month follow-up, colored by group (control, dark green; intervention, light green). Each dot represents the overall microbial composition of one sample, and samples that are closer together have more similar microbial communities. Shaded ellipses indicate 95% CI for each group, and group centroids are colored according to group with a red border for distinction. The axes (PCo1 and PCo2) show the first two principal coordinate axes, with percentage of explained variance denoted in parentheses. The *P*-values are from per protocol PERMANOVA performed separately at each visit. **(C)** Beta diversity (PCoA based on Bray-Curtis dissimilarity) in each group, colored by visit (baseline, blue; 3-month follow-up, orange). Arrows indicate individual trajectories from baseline to 3-month follow-up. The *P*-values are from per protocol PERMANOVA performed separately for each group. PCo, principal coordinate.

**Table 3.**
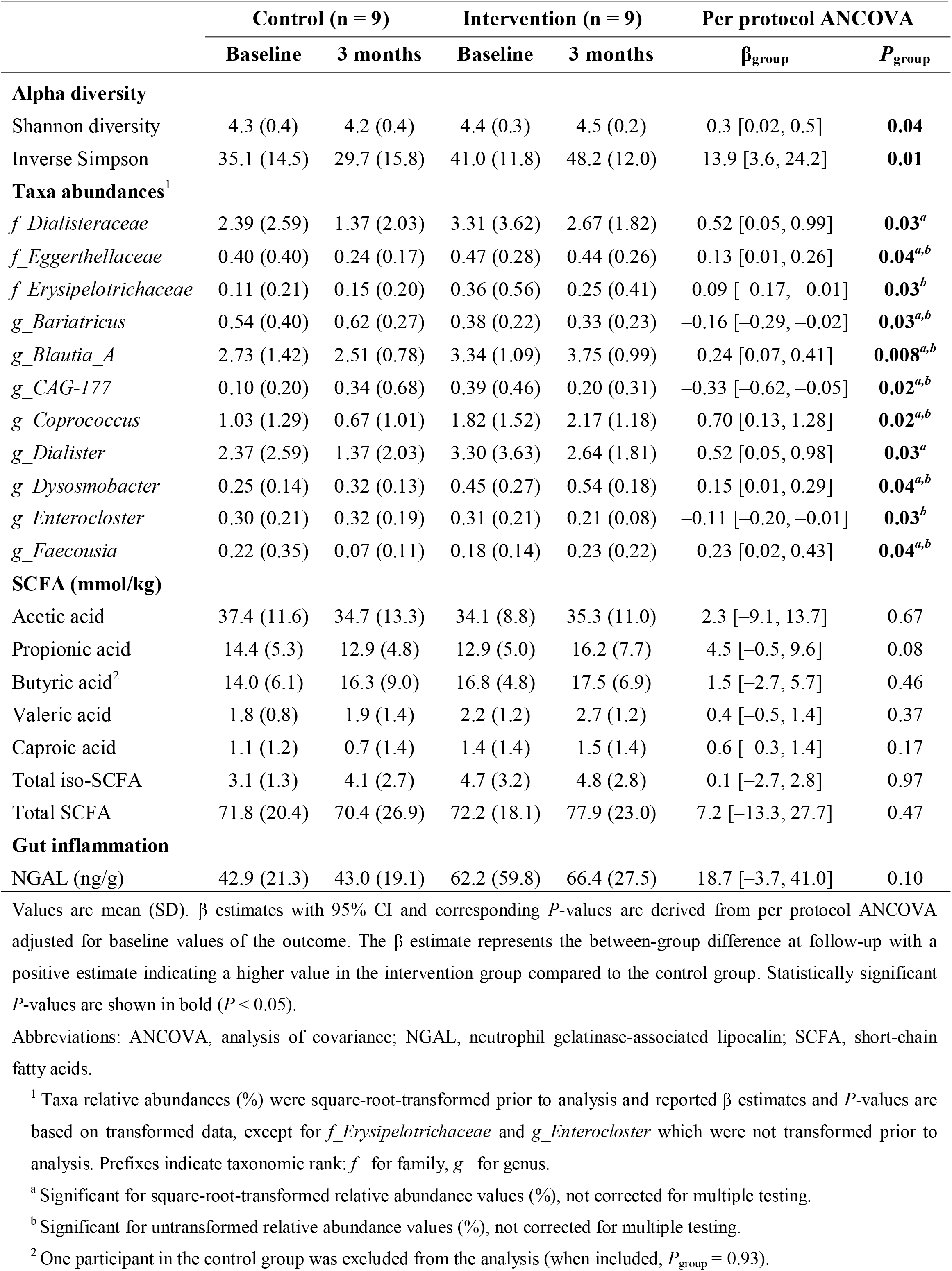
Fecal microbial diversity, taxa relative abundances and biomarkers at baseline and 3-month follow-up with estimated between-group differences at follow-up.

#### Overview of taxa abundances on different taxonomic levels

The gut microbiota profiles at phylum and genus level are visualized for each participant (Figures 4A and 4C) and as group means at each study visit (Figures 4B and 4D). Similar visualizations are presented at family level in **Supplementary Figure S23** (Supplemental Digital Content 1). The phyla with highest mean (SD) relative abundance across samples were *Bacillota_A* (61.3 (9.7)%), *Bacteroidota* (28.7 (10.1)%), *Bacillota_C* (2.8 (2.4)%), *Pseudomonadota* (2.0 (1.7)%) and *Actinomycetota* (2.0 (1.3)%). Also, some highly individual patterns were seen. For example, one participant displayed a high relative abundance of *Cyanobacteriota* at baseline (7.8%), which was largely absent from other samples (Figure 4A).

#### Microbial diversity and taxa relative abundances

The alpha diversity indices Shannon and Inverse Simpson (Figure 5A, Table 3), which reflect both richness (number of different taxa) and evenness (distribution of taxa abundances) within a microbial community, were higher in the intervention group than in the control group at 3-month follow-up when adjusting for baseline values (*P*_group_ = 0.04 and *P*_group_ = 0.01, respectively).

Beta diversity, reflecting overall differences in microbial community composition, was visualized using PCoA of Bray-Curtis dissimilarities (Figures 5B and 5C). At baseline, the centroids for the groups (i.e. the average position of each group in the multivariate space) overlapped, indicating similar composition between groups. By the 3-month follow-up, the group centroids shifted in different directions (rightward in the control group, leftward in the intervention group), but without clear separation (Figure 5B). Consistently, neither the overall PERMANOVA (group × visit interaction) nor the stratified PERMANOVAs at each visit showed significant between-group differences (**Supplementary Table S24**, Supplemental Digital Content 1). Also, no significant within-group shifts from baseline to follow-up were observed (Figure 5C, Supplementary Table S24). Taken together, the results indicate no clear systematic changes in overall microbial communities.

ANCOVA with baseline adjustment showed no significant between-group differences for the 171 taxa that were analyzed, when correcting for multiple testing using FDR. However, since FDR correction may become overly conservative or unreliable when sample sizes are small and the number of comparisons is high (38), the 11 taxa with nominal *P*_group_ < 0.05 were investigated further (Table 3, Supplementary Figure S25, Supplemental Digital Content 1).

Compared to controls, the intervention group displayed higher relative abundances of two families (*Dialisteraceae*, *Eggerthellaceae*) and five genera (*Blautia_A*, *Coprococcus*, *Dialister*, *Dysosmobacter*, *Faecousia*), and lower relative abundances of one family (*Erysipelotrichaceae*) and three genera (*Bariatricus*, *CAG-177*, *Enterocloster*) (**Supplementary Table S26**, Supplemental Digital Content 1, nominal and FDR-corrected *P*-values).

#### Microbial metabolites and gut inflammation

Neither of the individual SCFA nor the total SCFA concentrations were significantly affected by the low-dose iron supplementation (Table 3, Supplementary Figure S27B–H, Supplemental Digital Content 1). For fecal NGAL, we found a tendency toward higher levels in the intervention group compared to the control group, although the difference did not reach statistical significance (*P*_group_ = 0.10) (Supplementary Figure S27A, Supplemental Digital Content 1).

#### Exploratory correlation analyses

To explore potential associations between gut microbiota (i.e. taxa relative abundances) and other study variables, including blood and fecal biomarkers, we generated heatmaps of Spearman’s rank correlations at each taxonomic rank (**Supplementary Figures S28–S39**, Supplemental Digital Content 1). Heatmaps were created separately for baseline and follow-up samples, including all participants in the per protocol analysis (n = 18), as well as for calculated changes from baseline to follow-up for each group. Changes were calculated by subtracting the baseline value from the follow-up value. Hierarchical clustering was applied in both dimensions to visualize patterns across taxa and variables. As expected, physiologically related variables clustered together, such as serum ferritin with hepcidin and red blood cells with hemoglobin concentration. Apart from a negative association with *Verrucomicrobiota* (Spearman’s rank correlation coefficient*, r*_s_ = –0.71, *P* < 0.001), dietary iron intake showed no significant correlations with any of the assessed taxa at baseline (Supplementary Figures S28–S30). Among the iron-related biomarkers, hemoglobin and red blood cells correlated positively with *Bacillota_A*, *Clostridia* (both *r*_s_ = 0.68 and 0.69; *P* = 0.002 and 0.001, respectively), *Lachnospirales* and *Lachnospiraceae* (both *r*_s_ = 0.51 and 0.60; *P* = 0.03 and 0.009, respectively) at baseline (Supplementary Figures S28 and S29). These associations did not persist at follow-up. Instead, serum iron and transferrin saturation showed strong positive associations with *Actinomycetes*, *Actinomycetales* (both *r*_s_ = 0.61 and 0.69; *P* = 0.007 and 0.002, respectively) and *Bifidobacteriaceae* (*r*_s_ = 0.60 and 0.66; *P* = 0.009 and 0.003, respectively) and a strong negative association with *Enterobacteriaceae* (*r*_s_ = –0.76 and –0.65; *P* < 0.001 and 0.003, respectively) at follow-up (Supplementary Figures S31 and S32).

## DISCUSSION

The primary objectives of the current randomized controlled study were to characterize the dietary iron intake and iron status of young female soccer players and to investigate the effects of a 3-month low-dose iron supplementation (27 mg/day) on iron status, safety-related outcomes and gut microbiota. At baseline, we found that the majority (91%) of participants had inadequate dietary iron intake and 43% had serum ferritin concentrations indicative of depleted iron stores (< 35 µg/L (15)). The low-dose iron supplementation appeared to attenuate decreases in iron stores during the competitive soccer season, impacting several iron-related biomarkers. Furthermore, the supplementation was not associated with any GI symptoms, nor did it increase any of the safety-related biomarkers or induce any obvious unbeneficial effects on gut microbiota, indicating no adverse effects of the supplementation. On the contrary, we found higher microbial diversity and lower IL-6 levels in the iron-supplemented players compared to controls at 3-month follow-up.

To our knowledge, this is the first randomized controlled trial to investigate the effects of low-dose iron supplementation in female athletes while also assessing changes in the gut microbiota.

### Baseline dietary iron intake and iron status

Ensuring adequate iron intake is particularly important for female athletes who experience increased iron demands and losses related to menstruation and exercise (2). Consistent with previous reports of inadequate dietary iron intake (39, 40) and suboptimal serum ferritin levels (10, 11, 41) in female soccer players, 91% of the participants in the current study did not meet the RI for iron, and 43% had ferritin levels indicative of iron depletion (serum ferritin < 35 µg/L (15)). Similar findings were reported in young German elite female soccer players (n = 56), where 69% had inadequate iron intake (< 15 mg/day) and 59% had low serum ferritin levels (< 35 µg/L) (42). In our previous work, we also found a high prevalence (67%) of iron stores below 35 µg/L among Norwegian female soldiers (43), underscoring that suboptimal iron status is a common concern among physically active women. Importantly, the definition of iron depletion or deficiency in athletes is complicated by the lack of consensus on an appropriate ferritin threshold (3, 27). While guidelines have been proposed to increase uniformity around how iron status is assessed and reported in athletic populations, with ferritin < 35 µg/L typically used to indicate iron depletion (44), values below this threshold do not necessarily reflect compromised iron status or impaired performance (2). As we did not assess physical performance, we cannot draw conclusions about whether participants with ferritin below 35 µg/L experienced any functional consequences.

Dietary recommendations for iron also differ between countries (45). For example, the U.S. RDA for females is 18 mg iron per day (7), which would make the proportion of participants below recommendations in our study even higher. Comparisons across studies are further complicated by the use of different reference values to define adequacy, as some apply the RDA/RI while others use the estimated AR. In our study, the estimated median iron intake exceeded the AR of 9 mg/day according to NNR-2023 (5), with 96% of participants having an intake above this level. However, athletes may have 30–70% higher iron requirements (6), which would correspond to an AR of ∼12–15 mg/day. Based on these suggested athlete-specific requirements, only 8 (35%) and 2 (9%) participants in our study met the respective thresholds of 12 and 15 mg/day.

In the current study, grain products were the main contributor to total dietary iron intake (44%) followed by vegetables, fruits and berries (21%), suggesting that nonheme iron was the major iron constituent. Similar was reported in Polish female soccer players (n = 38) where cereal products contributed with 32% of the dietary iron (46). However, nonheme iron is absorbed less efficiently (∼2–20%) than heme iron (∼15–35%), which is derived from hemoglobin and myoglobin in animal foods, because the uptake of nonheme iron is strongly influenced by dietary enhancers (e.g. ascorbic acid) and inhibitors (e.g. phytates, polyphenols) (4). Of note, participants in the current study with high intakes of grain products had lower intakes of red meat (*r_p_*= –0.44, *P* = 0.04, Supplementary Figure S11, Supplemental Digital Content 1), which may partly explain the relatively high share of participants with suboptimal baseline iron status. In line with this, a recent study in a subset of the U.S. population reported that adherence to the American dietary guidelines, assessed using the Healthy Eating Index, supported dietary iron intake but not iron status (47). In that study, whole grains scores were positively associated with dietary iron intake but not with iron status, likely due to poor bioavailability, and dietary iron intake alone showed no association with iron status. Similarly, in competitive runners, diets with higher iron bioavailability were associated with better iron status, underscoring the importance of meal composition rather than total iron intake alone in maintaining iron balance (48). Thus, although sufficient iron intake can be achieved through diets rich in whole grains and iron-containing vegetables, it may require greater attention to dietary composition and iron bioavailability, particularly when intake of traditional iron-rich foods such as red meat is limited.

### Changes in iron status in response to iron supplementation

The significant increase in serum iron in the iron-supplemented group suggests good bioavailability of the low-dose iron supplementation. Moreover, participants with low baseline transferrin saturation experienced the largest increases at 3-month follow-up, indicating enhanced iron absorption in those with lower initial iron status. This pattern is consistent with the physiological regulation of iron homeostasis, where absorption is upregulated when the body’s iron stores are low (19), reflecting the central role of absorption rather than excretion in maintaining iron balance. It should be noted, however, that the increases in serum iron and transferrin saturation may partially reflect recent supplement intake, as we cannot exclude that some participants took their capsule on the morning of blood collection despite fasting instructions.

Previous studies have reported declines in ferritin in female soccer players during the competitive season (12). While the daily consumption of low-dose iron supplements in our study did not significantly increase serum ferritin, the share of participants having declines from baseline to follow-up were significantly lower in the iron-supplemented group compared to the control group. This finding suggests that low-dose iron supplementation may help attenuate reductions in iron stores during the competitive season, consistent with a study in female soldiers where eight weeks of low-dose iron supplementation (15 mg/day) attenuated reductions in iron stores during military training (49). Similarly, male soccer players showed decreased serum ferritin at the end of the season after discontinuing preventive iron supplementation (50 mg/day) (50). In contrast, Stefan et al. (51) reported that 27 mg of iron daily for eight weeks increased serum ferritin in nonanemic, iron-deficient premenopausal women (baseline ferritin < 30 µg/L) while Kang and Matsuo (30) found that 40 mg of iron daily for four weeks increased plasma ferritin in nonanemic, iron-deficient female soccer players (baseline ferritin < 30 µg/L). Such divergent results align with a recent systematic review and meta-analysis concluding that oral iron supplementation increases serum ferritin in athletes with low initial ferritin, but has minimal effects in those with higher baseline stores (27). The absence of a significant effect on ferritin in our study may also be due to limited statistical power, as discussed under limitations.

Although serum TIBC levels did not differ significantly between groups, the observed decrease in the iron-supplemented group may reflect an early regulation of iron homeostasis, with downregulated transferrin expression in response to increased iron availability (27). In contrast, hepcidin, the liver-derived peptide hormone that regulates iron absorption and release from enterocytes (dietary iron), macrophages (recycled iron) and hepatocytes (stored iron) (20), was not significantly altered by the intervention. This may indicate that a larger or more rapid increase in iron availability is required to trigger a measurable hepcidin response, which is consistent with previous studies showing that high (> 60 mg) but not low (≤ 40 mg) iron doses significantly increased hepcidin levels (29). Also, as hepcidin synthesis increases in response to inflammation (primarily via IL-6) and elevated iron stores (20), the lack of significant changes in hepcidin in our study aligns with the absence of significant increases in both ferritin and inflammatory biomarkers. However, in exploratory correlation analyses, we observed a strong positive relationship between changes in serum hepcidin and ferritin from baseline to follow-up (*r_s_*= 0.67, *P* = 0.002), consistent with physiological expectations (**Supplementary Figure S41**, Supplemental Digital Content 1).

As for hemoglobin and red blood cell counts, the lack of significant between-group differences suggests that iron supplementation was not required to support erythropoiesis and that the participants’ iron stores were sufficient to maintain normal red blood cell production during the competitive season. Indeed, all participants had hemoglobin levels within the normal range at both baseline and 3-month follow-up. In line with our findings, a previous trial in iron-deficient athletes (n = 20) with normal baseline hemoglobin reported no increase in hemoglobin concentrations after six weeks of 30 mg iron daily compared with placebo (52). However, low-dose iron therapy (6 mg twice daily for eight weeks) increased hemoglobin in nonathletic, iron-deficient women with normal baseline hemoglobin levels (n = 36) (53). These contradictory findings between athletic and nonathletic populations may be explained by exercise-induced plasma volume expansion, which dilutes circulating hemoglobin and red blood cell concentrations (54) and has been observed in soccer players (55). Therefore, measurement of total hemoglobin mass would provide a more reliable assessment of true erythropoietic changes, while estimation of plasma volume would aid the interpretation of concentration-based markers (56). Additional exercise-related factors, such as foot-strike hemolysis, may also influence hemoglobin and red blood cell concentrations in athletes (8), particularly when iron recycling from senescent erythrocytes is constrained by elevated hepcidin levels (20).

### Adverse effects of iron supplementation

In our study, none of the participants reported GI-related symptoms at 3-month follow-up and no significant differences in stool consistency were found, indicating that the daily dose of 27 mg was well tolerated. This finding aligns with a trial in nonanemic, iron-deficient women using the same iron dose (27 mg) provided as a liquid fermented iron-bisglycinate supplement (51). Given that fractional iron absorption is greater at lower doses, the low dose applied in our study likely limited the amount of unabsorbed iron reaching the gut lumen, which may have reduced GI irritation (28). Moreover, we used ferrous bisglycinate, a chelated form of iron that has been shown to cause fewer side effects compared to ferrous sulphate (4). It should be noted that we assessed adverse symptoms using a non-systematic open-ended question asking about health-related issues during the past week rather than a validated questionnaire, which is discussed further under limitations.

A rise in biomarkers of oxidative stress and/or inflammation could also indicate potential adverse effects related to excessive iron intake (22). However, in the current study, we found no adverse changes in the safety-related biomarkers, including no increases in inflammatory markers or serum GGT, a biomarker of liver diseases also related to oxidative stress (57). On the contrary, the iron-supplemented group showed lower serum IL-6 levels than controls at 3-month follow-up. This finding is relevant as exercise can trigger an inflammatory response that increases cytokines such as IL-6 (8), and elevated IL-6 levels have been observed during the soccer season in female players (12). However, while cytokines are often associated with inflammation and illness, in the context of exercise, muscle-derived IL-6 also plays an important regulatory role that contributes to physiological adaptations (58). Although the clinical relevance of the reduced IL-6 levels in response to iron supplementation is unclear, one could speculate that it may reflect enhanced adaptation to accumulated training load, or relate to iron’s involvement in the immune system where it supports immune cell growth, differentiation and function (59). The within-group decreases in ALAT and ASAT may also relate to potential training adaptation.

### Changes in gut microbiota and gut inflammation in response to iron supplementation

Because free iron in the host is tightly regulated and scarce, alterations in the availability of unbound iron in the gut lumen may strongly influence the microbial ecosystem (60). Despite increasing interest in the interplay between diet, exercise and the gut microbiota, evidence from clinical trials regarding iron supplementation effects remains limited and inconsistent (26, 60). Specifically, to our knowledge, there are no studies investigating the impact of iron supplementation on gut microbiota in athletes. In the current study, low-dose iron supplementation over three months led to higher alpha diversity compared to controls, who showed a pattern of decreasing diversity. This contrasts with findings from Cambodian women receiving either 18 mg ferrous bisglycinate or 60 mg ferrous sulfate daily, where iron supplementation did not alter gut microbial diversity compared with placebo (61). Similarly, Celis et al. (62) reported that supplementing healthy adults with 65 mg iron daily did not alter alpha diversity. However, these studies were conducted in non-athletic participants, limiting the comparability to our study population. Interestingly, Celis et al. (62) also performed *in vitro* experiments using fecal samples from the study participants and found that iron deprivation caused marked reductions in community richness and loss of species, suggesting that iron limitation may potentially induce adverse effects on gut microbial diversity.

Despite the increase in alpha diversity, we detected no significant differences in beta diversity and only modest shifts in individual taxa, without effects on fecal SCFA concentrations. These findings, together with the absence of GI-related adverse events and stable hepcidin levels, suggest that the low-dose iron supplementation did not lead to luminal iron accumulation and thus had minimal impact on overall gut community structure. Consistent with this, a retrospective study of middle-aged women (n = 56) that compared no iron supplementation, low-dose (6–10 mg/day) and high-dose (> 100 mg/day) iron intake reported a dose-dependent effect on gut microbiota composition (63). While the strongest impact was observed in the high-dose group, the authors also reported significant differences in beta diversity between the no-supplementation and low-dose groups (63). It is possible that the lack of significant effects on beta diversity in our study is due to the limited sample size, as the opposite shifts in group centroids observed in the PCoA plots at 3-month follow-up may indicate an early or subtle divergence.

The work of others also supports our observation of minimal effects of iron supplementation on taxa relative abundances. Elms et al. (64) reported that 65.7 mg of iron daily for 21 days did not alter the gut microbiota of females of reproductive age compared with placebo. Dekker Nitert et al. (65) likewise found that neither low-dose (≤ 10 mg) nor high-dose (≥ 60 mg) iron supplements significantly changed the gut microbiota composition in pregnant women with overweight or obesity. In contrast, one study by Finlayson-Trick et al. (61) observed that a similar dose as the current trial (18 mg/day as ferrous bisglycinate) for 12 weeks increased the relative abundance of *Enterobacteriaceae*, a family that includes several enteric pathogens, although neither alpha nor beta diversity differed significantly from controls.

In our study, prior to multiple testing correction, a few taxa differed between the groups at 3-month follow-up, including the family *Dialisteraceae* and its genus *Dialister*, which were more abundant in the iron-supplemented group. Moreover, changes in their abundance from baseline to follow-up showed positive, albeit non-significant, associations with systemic iron-related biomarkers (Supplementary Figures S33–S36, Supplemental Digital Content 1). One possible explanation could be that *Dialister* bacteria may be less competitive when luminal iron is scarce. The finding also aligns with a recent review reporting that the proportion of *Dialister* increased in response to iron supplementation and decreased during iron deficiency (60). Another peculiar finding in our study was the high relative abundance of *Cyanobacteriota* in one of the participants at baseline who also reported the highest intake of fish and shellfish (**Supplementary Figure S40**, Supplemental Digital Content 1), mostly from sushi. This may indicate a transient diet-derived increase due to seaweed (nori) intake (66); however, the potential use of *Cyanobacteriota* as a biomarker for sushi intake remains to be established.

Bacteria use several strategies to acquire iron in iron-limited environments, one of which is the secretion of siderophores (67). In the current study, we found that iron supplementation tended to increase fecal NGAL, a small glycoprotein expressed in several tissues including the intestinal epithelium (68). NGAL works by binding bacterial ferric-siderophore complexes, thereby limiting iron availability for the bacteria and restricting their growth (67). Because fecal NGAL increases in response to bacterial infection, gut inflammation and epithelial injury, it has emerged as a promising biomarker of gut inflammation (32, 68). The tendency toward higher fecal NGAL in our study in the iron-supplemented group could therefore reflect mild gut inflammation. However, as we did not observe a systemic inflammatory response (serum CRP, NGAL, LBP, cytokines), and only modest shifts in the gut microbiota composition, it is more likely that its increase reflects altered iron availability. In line with this, NGAL has been suggested to play a role in iron homeostasis, possibly acting as a protective mechanism against iron overload and associated adverse effects (69). In exploratory analyses, we observed a positive, albeit non-significant, correlation between changes in fecal NGAL and ferritin (*r_s_* = 0.38, *P* = 0.12, Supplementary Figure S41, Supplemental Digital Content 1). While this observation does not support a definitive association, it may warrant further investigation of potential links between intestinal iron handling and systemic iron markers.

### Limitations

While the current study followed a randomized controlled trial design, considered the gold standard for evaluating intervention effects, it was not placebo-controlled. Therefore, we cannot exclude potential psychological effects, such as participants’ expectations regarding the benefits or side effects of iron supplementation, or the possibility that lack of blinding affected eating behavior. However, participants were not informed of any anticipated benefits or adverse effects, and the outcomes assessed (blood and fecal biomarkers) are less likely to be influenced by placebo effects. Follow-up questionnaires indicated no major dietary changes, and the observed group differences in serum iron and transferrin saturation suggest that the control group did not substantially alter their iron intake. Furthermore, we did not provide standardized instructions on when to take the iron supplements, which may have influenced absorption efficiency given the exercise-induced fluctuations in hepcidin (4). The use of an open-ended question to assess adverse symptoms also has some limitations. While this approach may reduce overreporting that can occur when participants are asked about specific symptoms via checklists (70), it may also underestimate milder or less noticeable symptoms.

Another limitation of this study is the relatively small sample size. The estimates used in the power calculation were optimistic, particularly given the relatively low iron dose, and based on much larger expected increase in ferritin (55 µg/L) than what we observed (9.2 µg/L). Thus, it is likely that the study was underpowered to detect subtle, yet potentially meaningful effects. Also, while we collected information on the timing and duration of the last menstrual cycle at baseline and follow-up, we did not record the number of cycles during the study period.

The digital food diary used in the current study had not been previously validated. However, it was based on a validated paper version, and we evaluated its performance using serum carotenoids which indicated good performance in terms of intake of vegetables, fruits and berries. Furthermore, the digital form was basically paper-on-a-screen with no database and minimal prompts. Thus, compared to the paper version, the digital diary is unlikely to systematically over- or underestimate iron intake just because it is digital. In addition, the option to upload photographs of foods and ingredients may have improved the accuracy of iron intake estimation. However, the dietary assessment tool did not allow differentiation between heme and nonheme iron, limiting our ability to account for differences in dietary iron bioavailability. Finally, we did not have detailed training data which could have helped account for exercise’s known effects on iron status and inflammation (4). However, the participating soccer clubs followed a strict training schedule, and it is likely that the workload was similar among participants.

## CONCLUSION

In conclusion, the current study population of female soccer players had inadequate baseline iron intake, with 91% of the participants reporting intakes below the recommended level. Our findings suggest that the low-dose iron supplementation (27 mg/day) may aid in maintaining iron status during the competitive season in female soccer players without causing adverse effects. On the contrary, increased microbial alpha diversity and decreased level of the systemic inflammatory biomarker IL-6 might reflect potential beneficial effects.

The findings of the current study underscore the need for larger-scale studies to confirm the efficacy of low-dose iron supplementation on iron status and to comprehensively evaluate the safety of a longer-term supplementation regimen, particularly in terms of the gut microbiota. If shown to be both effective and safe, a steady low-dose iron supplementation could represent a valuable strategy for maintaining iron status and performance in female athletes throughout the competitive season, particularly in circumstances when it is difficult to achieve sufficient iron intake through diet alone. Whether higher iron doses will lead to greater increases in overall iron status without increase in adverse effects remains to be investigated.

## Supporting information

Supplemental Digital Content 1

## ABBREVIATIONS

ALAT: alanine aminotransferase
ANCOVA: analysis of covariance
ASAT: aspartate aminotransferase
AR: average requirement
BMI: body mass index
BSFS: Bristol stool form scale
CI: confidence interval
CRP: C-reactive protein
CV: coefficient of variation
DPBS: Dulbecco’s phosphate buffered saline
ELISA: enzyme-linked immunosorbent assay
FDR: false discovery rate
GGT: gamma-glutamyl transferase
GM-CSF: granulocyte-macrophage colony-stimulating factor
GTDB: genome taxonomy database
IFN-γ: interferon-gamma
IL: interleukin
LBP: lipopolysaccharide-binding protein
NGAL: neutrophil gelatinase-associated lipocalin (also known as lipocalin-2)
NNR: Nordic nutrition recommendations
OTU: operational taxonomic unit
PCoA: principal coordinate analysis
PERMANOVA: permutational multivariate analysis of variance
Q: quartile
RDA: recommended dietary allowance
RI: recommended intake
rRNA: ribosomal RNA
SCFA: short-chain fatty acids
SD: standard deviation
S.T.A.R.: stool transport and recovery
TIBC: total iron-binding capacity
TNF-α: tumor necrosis factor-alpha.

## ACKNOWLEDGEMENTS

The authors would like to thank Lars Fredrik Moen for assistance with blood sampling; Haakon Breien Benestad for serving as medical supervisor during the trial; Gunn Helen Malmstrøm for performing SCFA analysis; and Tonje Nilsen and Inga Leena Angell for assistance and technical support during sequencing. We sincerely thank the study participants for their valuable contribution and commitment to the study.

The sequencing service was provided by the Norwegian Sequencing Centre (www.sequencing.uio.no), a national technology platform hosted by the University of Oslo and Oslo University Hospital and supported by the Research Council of Norway and the Southeastern Regional Health Authorities.

## Author contributions

**SSS**, **GP**, **TR** and **SKB** designed the research; **SSS**, **TEA**, **GP**, **TR** and **SKB** conducted the research; **MHC**, **KR**, **JV** and **SKB** provided essential materials; **SSS**, **TEA**, **LMG**, **AMH** and **NEB** performed the laboratory analyses; **MCH** and **SKB** developed the dietary assessment method; **SSS** performed the statistical analyses and data visualizations, processed the sequencing data and coded the food diaries; **MCH** performed the food and nutrient estimations; **AKM** contributed to interpretation of the nutritional reports to the participants; and **SSS** and **SKB** wrote the paper. **SKB** had primary responsibility for the final content. All authors commented on draft manuscripts and approved the final version of the manuscript.

## DATA AVAILABILITY STATEMENT

Data collected and produced in the present study may be available upon reasonable request pending approval by relevant data access committees, ethical committees and privacy regulation authorities. Requests to access the datasets should be directed to the corresponding author.

## DECLARATION OF AI USE IN THE WRITING PROCESS

During the preparation of this manuscript, the authors used ChatGPT (chat.openai.com) to improve clarity and readability of the text. All content was subsequently reviewed and edited by the authors, who take full responsibility for the content of the publication.

## SUPPLEMENTAL DIGITAL CONTENT

Supplemental digital content 1. pdf

