## Supplemental Digital Content 1 for "Effects of low-dose iron supplementation on iron status, safety outcomes and gut microbiota in female soccer players: a randomized controlled study"

Stine Sofie Strømland[1], Trude Elise Aspholm[1], Gøran Paulsen[2], Monica Hauger Carlsen[3], Lisa Medina Grimestad[1], Anne Mari Herfindal[1], Anu Koivisto-Mørk[4], Nasser Ezzatkhah Bastani[3], Knut Rudi[1], Jørgen Valeur[5,6], Truls Raastad[2], Siv Kjølsrud Bøhn[1]

[1] Faculty of Chemistry, Biotechnology and Food Science, Norwegian University of Life Sciences, Ås, Norway

[2] Department of Physical Performance, Norwegian School of Sport Sciences, Oslo, Norway

[3] Department of Nutrition, Institute of Basic Medical Sciences, University of Oslo, Oslo, Norway

[4] The Norwegian Football Association's Sports Medicine Center (Idrettens Helsesenter), Oslo, Norway

[5] Unger-Vetlesens Institute, Lovisenberg Diaconal Hospital, Oslo, Norway

[6] Department of Gastroenterology and Hepatology, Institute of Clinical Medicine, University of Oslo, Oslo, Norway

Effects of low-dose iron supplementation on iron status, safety outcomes and gut microbiota in female soccer players: a randomized controlled study, Strømland et al.

### 1. STUDY DESIGN

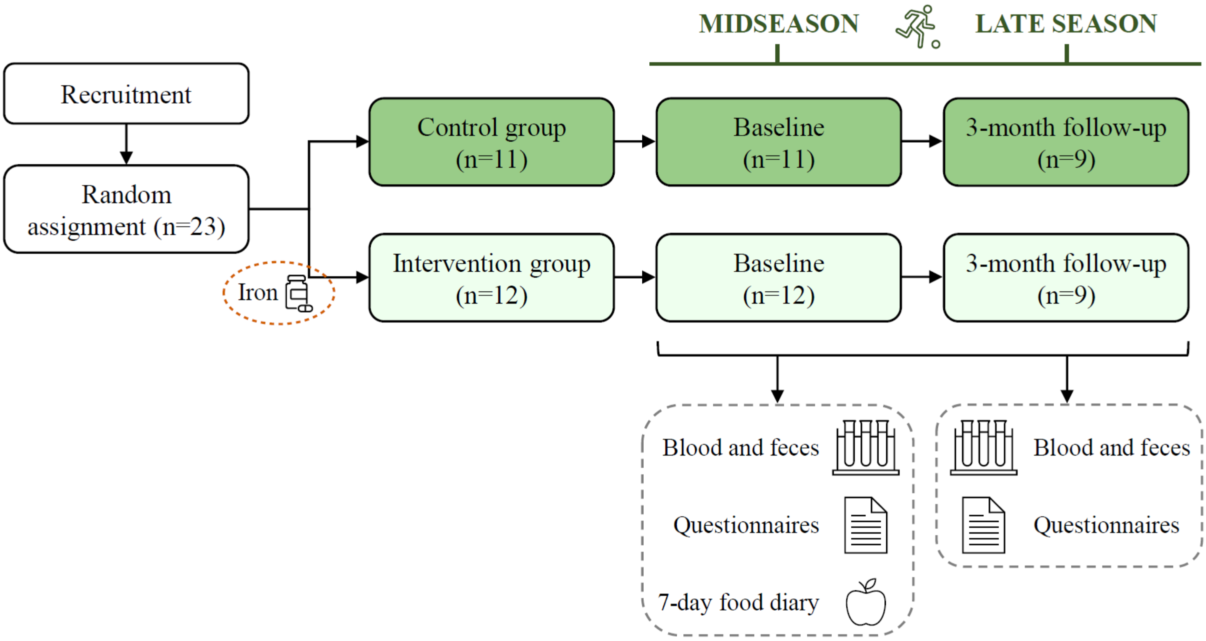

**Supplementary Figure S1**. Overview of the study design. Participants were followed from midseason (baseline) to late season (3-month follow-up). At each study visit, participants delivered biological samples (blood and feces) and filled out questionnaires (background information, menstrual cycle and Bristol stool form scale). Additionally, 7-day food diaries were submitted at baseline. Nine participants per group were included in per protocol analysis.

### 2. MULTIPLEX ASSAY

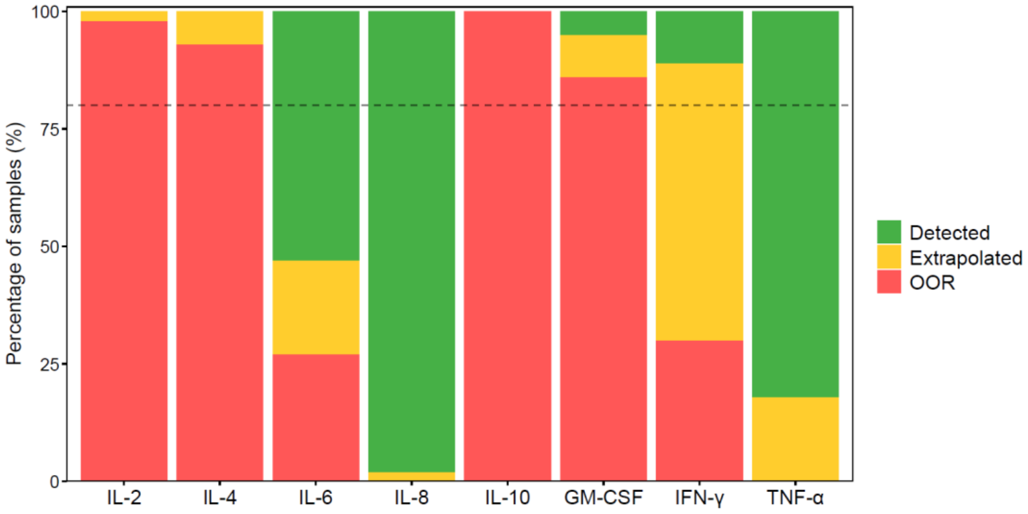

**Supplementary Figure S2**. Sample detectability. A total of 44 samples (baseline, n = 23; follow-up, n = 21) were analyzed. For each analyte (bar), green indicates the proportion of samples with detectable concentrations, yellow indicates extrapolated values and red indicates samples below the limit of detection. Analytes with more than 80% OOR values (IL-2, IL-4, IL-10, and GM-CSF) were excluded from further analysis. GM-CSF, granulocyte-macrophage colony-stimulating factor; IL, interleukin; IFN-γ, interferon-gamma; OOR, out-of-range; TNF-α, tumor necrosis factor-alpha.

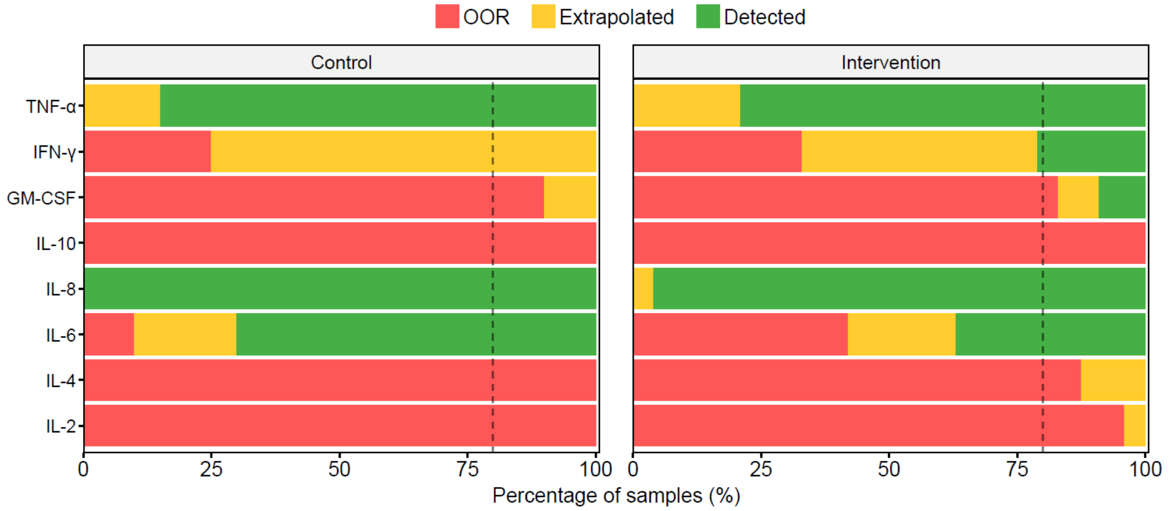

**Supplementary Figure S3**. Sample detectability by study group. The number of samples analyzed was 20 (n = 11 participants) in the control group and 24 (n = 12 participants) in the intervention group. For each analyte (bar), green indicates the proportion of samples with detectable concentrations, yellow indicates extrapolated values and red indicates samples below the limit of detection. GM-CSF, granulocyte-macrophage colony-stimulating factor; IL, interleukin; IFN-γ, interferon-gamma; OOR, out-of-range; TNF-α, tumor necrosis factor-alpha.

**Supplementary Table S4**. Overview of imputed samples in the multiplex cytokine analysis.

| Analyte | Lowest detectable concentration (pg/mL) | Number of imputed samples (n) | Percentage of imputed samples (%) |
|---|---|---|---|
| IL-6 | 0.10 | 12 | 27 |
| IFN-γ | 0.22 | 13 | 30 |

Imputation was performed using maximum likelihood estimation, assuming a log-normal distribution. The lowest measured concentration was set as the limit of quantification. Imputation was not performed for analytes with more than 80% out-of-range values (Supplementary Figures S2 and S3).
Abbreviations: IFN-γ, interferon-gamma; IL, interleukin.

### 3. SCFA ANALYSIS

For determination of SCFA concentrations, approximately 0.5 g of fecal material was homogenized in distilled water containing 3 mmol/L 2-ethylbutyric acid ($C_6H_{12}O_2$) as an internal standard and 0.5 mmol/L sulfuric acid ($H_2SO_4$). Then, 2.5 mL of the homogenate was vacuum distilled according to the method described by Zijlstra et al. (1), as modified by Høverstad et al. (2). The distillate was analyzed with gas chromatography (Agilent 6850, CA, USA) using a capillary column (Serial No. USE400311H, Agilent J&W GC columns, CA, USA) and quantified by internal standardization. Flame ionization detection was employed.

### 4. 16S rRNA SEQUENCING

#### 4.1 Library preparation

##### DNA extraction

DNA extraction was performed using a combination of mechanical (bead-beating) and chemical lysis. Fecal aliquots (approximately 0.2 g mixed with 600 µL S.T.A.R. buffer) were thawed and homogenized by vortexing before 300 µL of each suspension was transferred to tubes containing acid-washed glass beads (approximately 0.2 g of < 106 µm and 425–600 µm beads along with two beads of 2.5–3.5 mm, Sigma-Aldrich). Samples were processed twice on a FastPrep-96 instrument at 1,800 rpm for 40 seconds, with a 5-minute rest between runs, to obtain cell lysis. After centrifugation (15,871 × g, 10 minutes, 4°C), 50 µL of each supernatant was transferred to a 96-well microtiter plate for proteinase K treatment followed by DNA extraction using the Mag Midi LGC kit (Cat. No. NAP40420, Biosearch technologies, LGC Genomics GmbH, Germany). The procedure was conducted according to the manufacturer's instructions using a KingFisher Flex automated system (Thermo Scientific). To verify successful extraction, DNA concentrations were quantified using the Qubit dsDNA HS kit (Cat. No. Q33231) and a Qubit 2.0 Fluorometer (Invitrogen).

**Amplicon PCR**

Extracted DNA was diluted 1:10 in nuclease-free water prior to PCR amplification to obtain the appropriate DNA amount (0.1–10 ng). The 16S rRNA gene was amplified using primers targeting the V3–V4 region: forward primer PRK341F, 5'-CCTACGGGRBGCASCAG-3' and reverse primer PRK806R, 5'-GGACTACYVGGGTATCTAAT-3' (3). PCR amplification was performed on a 2720 Thermal Cycler (Applied Biosystems) under the following conditions: initial denaturation at 95°C for 15 minutes, 25 cycles of denaturation at 95°C for 30 seconds, annealing at 55°C for 30 seconds and extension at 72°C for 45 seconds, and a final extension at 72°C for 7 minutes. PCR amplicon quality was assessed by gel electrophoresis (1.5% agarose gel with 2 µL peQ green per 50 mL gel mix; 100 bp DNA ladder, Solis BioDyne) to confirm fragment size (~466 bp). PCR products were purified using AMPure XP beads (Sera Mag Beads), including two ethanol wash steps prior to elution in nuclease-free water.

**Index PCR**

Unique combinations of index primers modified with Illumina adapters (**Supplementary Table S5**, primer sequences) were added to each sample using an epMotion 5070 automated pipetting system (eppendorf). PCR amplification was performed under the following conditions: initial denaturation at 95°C for 5 minutes, 10 cycles of denaturation at 95°C for 30 seconds, annealing at 55°C for 1 minute and extension at 72°C for 45 seconds, and a final extension at 72°C for 7 minutes. Fragment size (~594 bp) was confirmed by gel electrophoresis (1.5% agarose gel).

**Illumina sequencing**

All samples were pooled based on DNA concentration (normalized to 60 ng) followed by purification (AMPure XP beads) and quantification of DNA concentration (Qubit dsDNA HS kit and Qubit 2.0 fluorometer). The pooled library was diluted to ~18 ng/µL in nuclease-free water before amplicon size (~550–600 bp) was determined by gel electrophoresis (2.0% agarose gel). The final amplicon library was stored at –20°C until shipment on ice to the Norwegian Sequencing Centre (Oslo, Norway) for 16S rRNA gene sequencing on the Illumina MiSeq v3 platform (300 bp paired-end reads).

**4.2 Sequence processing**

Raw Illumina sequencing reads were processed using the open-source tool VSEARCH (v.2.28.1--h6a68c12_0) (4) as an alternative to USEARCH (5). Reads were demultiplexed based on unique barcode sequences, trimmed (R2, 60 bp; R1, 20 bp), merged and quality filtered (average error prob tolerated, P = 0.01, corresponds to Q = 20). Denoising was performed to generate zero-radius operational taxonomic units (zOTUs) (6), hereby referred to as 'OTUs', which represent unique, high-resolution sequence variants analogous to amplicon sequence variants (ASVs). Unlike traditional OTU clustering, this approach resolves sequences at single-nucleotide resolution without applying a predefined similarity threshold (7).

To account for variation in sequencing depth across samples, total sum scaling was applied to convert raw read counts to relative abundances (%) prior to downstream analyses. Samples were not rarefied (i.e. subsampled to equal sequencing depth) to preserve statistical power and avoid introducing artificial noise (8). Alpha diversity indices (Shannon and Inverse Simpson) assessed in this paper did not correlate significantly with library size ($P > 0.05$, data not shown).

Taxonomic classification of OTUs was performed using the SINTAX algorithm (9) with the Genome Taxonomy Database (GTDB) release 2.20 as reference (10). In contrast to conventional 16S-based taxonomies, GTDB is based on whole-genome phylogenetic reconstruction and provides a more accurate representation of microbial evolutionary relationships (11). Taxonomy was assigned using a confidence threshold of 0.8, and classifications below this threshold were categorized as unclassified at each respective rank. Excluding unclassified taxa, a total of 14 phyla, 17 classes, 35 orders, 77 families and 240 genera were identified across the 36 samples. Of note, one OTU was inconsistently classified across taxonomic databases, being assigned to the domain *Bacteria* (genus *Ruminococcus*) by GTDB but to *Archaea* (genus *Methanobrevibacter*) by all other databases evaluated, including RDP, SILVA and GSR (data not shown). Given the consensus among alternative databases, and that an *Archaea* lineage was also among the top GTDB hits (genus *Methanobrevibacter_A*, ranked third), this OTU was manually reassigned to the *Archaea* domain.

#### 4.3 Index primers

**Supplementary Table S5**. Index primers modified with Illumina adapters used during library preparation for 16S rRNA sequencing. A unique combination of forward and reverse primers was added to each sample for downstream demultiplexing of pooled samples.

| Primer name | Primer sequence (5' → 3') | Target gene (region) | Direction |
|---|---|---|---|
| F1 | aatgatacggcgaccaccgagatctacactctttccctacacgacgctcttccgatctagtcaaCCTACGGGRBGCASCAG | 16S rRNA (V3–V4) | Forward |
| F2 | aatgatacggcgaccaccgagatctacactctttccctacacgacgctcttccgatctagttccCCTACGGGRBGCASCAG | 16S rRNA (V3–V4) | Forward |
| F3 | aatgatacggcgaccaccgagatctacactctttccctacacgacgctcttccgatctatgtcaCCTACGGGRBGCASCAG | 16S rRNA (V3–V4) | Forward |
| F4 | aatgatacggcgaccaccgagatctacactctttccctacacgacgctcttccgatctccgtccCCTACGGGRBGCASCAG | 16S rRNA (V3–V4) | Forward |
| F5 | aatgatacggcgaccaccgagatctacactctttccctacacgacgctcttccgatctgtagagCCTACGGGRBGCASCAG | 16S rRNA (V3–V4) | Forward |
| F6 | aatgatacggcgaccaccgagatctacactctttccctacacgacgctcttccgatctgtccgcCCTACGGGRBGCASCAG | 16S rRNA (V3–V4) | Forward |
| F7 | aatgatacggcgaccaccgagatctacactctttccctacacgacgctcttccgatctgtgaaaCCTACGGGRBGCASCAG | 16S rRNA (V3–V4) | Forward |
| F8 | aatgatacggcgaccaccgagatctacactctttccctacacgacgctcttccgatctgtggccCCTACGGGRBGCASCAG | 16S rRNA (V3–V4) | Forward |
| R1 | caagcagaagacggcatacgagatCGTGATgtgactggagttcagacgtgtgctcttccgatctGGACTACYVGGGTATCTAAT | 16S rRNA (V3–V4) | Reverse |
| R2 | caagcagaagacggcatacgagatACATCGgtgactggagttcagacgtgtgctcttccgatctGGACTACYVGGGTATCTAAT | 16S rRNA (V3–V4) | Reverse |
| R3 | caagcagaagacggcatacgagatGCCTAAgtgactggagttcagacgtgtgctcttccgatctGGACTACYVGGGTATCTAAT | 16S rRNA (V3–V4) | Reverse |
| R4 | caagcagaagacggcatacgagatTGGTCAgtgactggagttcagacgtgtgctcttccgatctGGACTACYVGGGTATCTAAT | 16S rRNA (V3–V4) | Reverse |
| R5 | caagcagaagacggcatacgagatCACTCTgtgactggagttcagacgtgtgctcttccgatctGGACTACYVGGGTATCTAAT | 16S rRNA (V3–V4) | Reverse |
| R6 | caagcagaagacggcatacgagatATTGGCgtgactggagttcagacgtgtgctcttccgatctGGACTACYVGGGTATCTAAT | 16S rRNA (V3–V4) | Reverse |

#### 4.4 Library sizes

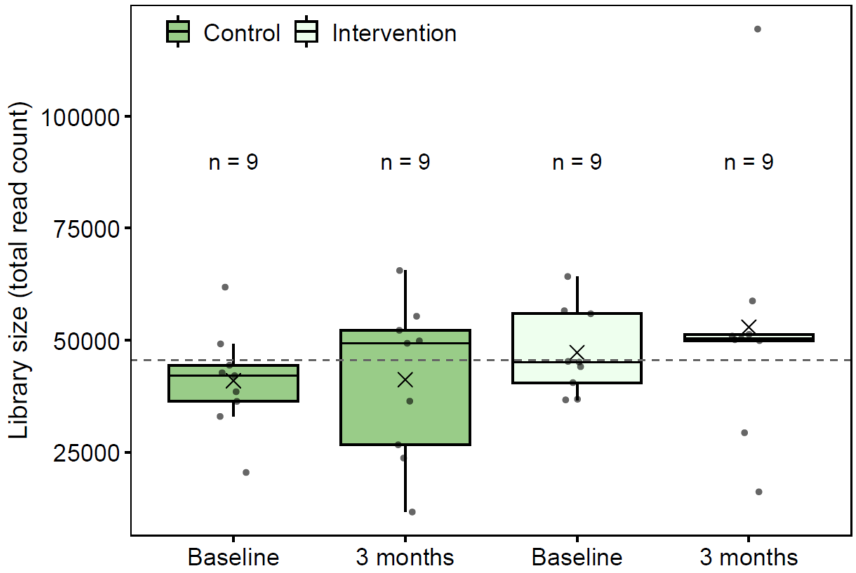

**Supplementary Figure S6**. Library sizes (total read counts per sample) at baseline and 3-month follow-up in the control (dark green) and intervention (light green) group. Each dot represents one participant, and the number of participants is shown above each box. The dashed horizontal line indicates the overall mean library size, and the cross within each box indicates the group mean at each visit.

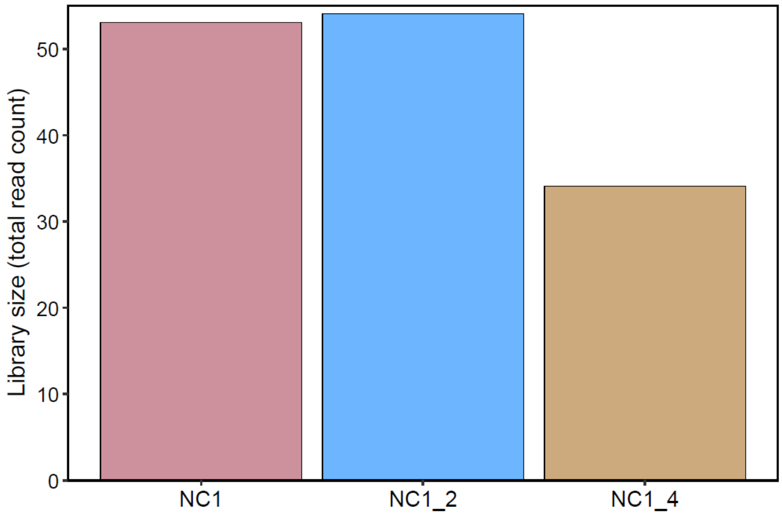

**Supplementary Figure S7**. Library sizes (total read counts) of the negative control samples included in the 16S rRNA sequencing. NC, negative control.

#### 4.5 Principal coordinate analysis

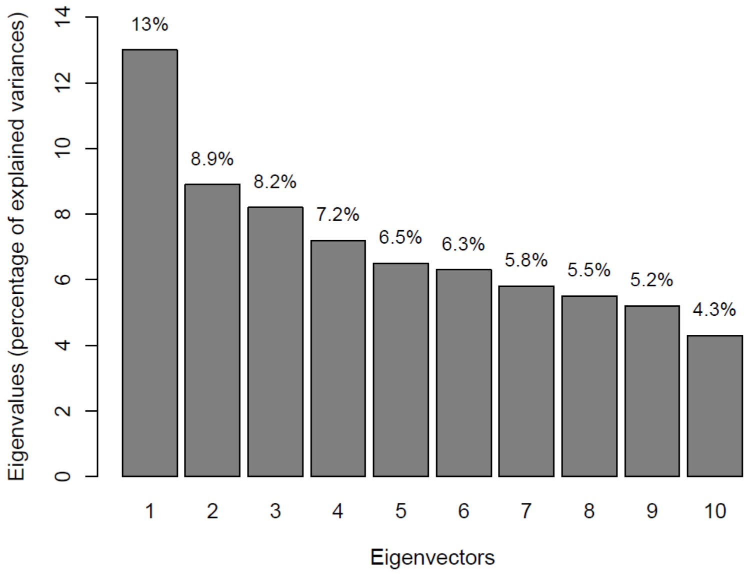

**Supplementary Figure S8**. Scree plot showing the proportion of variation explained by the first 10 principal coordinate axes from the PCoA based on Bray-Curtis dissimilarities. The plot illustrates how much variation each axis accounts for in the beta diversity analysis.

#### 4.6 Differential abundance analysis

**Supplementary Table S9**. Summary statistics for taxa (n = 171) included in the differential abundance analysis (having a median relative abundance ≥ 0.01% in at least one of the two study groups), arranged alphabetically within each rank. For each study group, the mean and median relative abundance (%) across both visits are reported, along with the number and percentage of samples in which the taxon was present.

| Taxa | Rank | Group | Present (%) | Present (n) | Absent (n) | Mean | Median |
|---|---|---|---|---|---|---|---|
| *Actinomycetota* | Phylum | Control | 100 | 18 | 0 | 2.013 | 1.928 |
| *Actinomycetota* | Phylum | Intervention | 100 | 18 | 0 | 2.043 | 2.196 |
| *Bacillota* | Phylum | Control | 100 | 18 | 0 | 0.514 | 0.294 |
| *Bacillota* | Phylum | Intervention | 100 | 18 | 0 | 0.373 | 0.374 |
| *Bacillota_A* | Phylum | Control | 100 | 18 | 0 | 60.759 | 58.866 |
| *Bacillota_A* | Phylum | Intervention | 100 | 18 | 0 | 61.864 | 64.027 |
| *Bacillota_B* | Phylum | Control | 61 | 11 | 7 | 0.054 | 0.007 |
| *Bacillota_B* | Phylum | Intervention | 89 | 16 | 2 | 0.128 | 0.082 |
| *Bacillota_C* | Phylum | Control | 100 | 18 | 0 | 2.422 | 1.774 |
| *Bacillota_C* | Phylum | Intervention | 100 | 18 | 0 | 3.185 | 2.522 |
| *Bacillota_I* | Phylum | Control | 100 | 18 | 0 | 1.971 | 0.361 |
| *Bacillota_I* | Phylum | Intervention | 100 | 18 | 0 | 0.591 | 0.466 |
| *Bacteroidota* | Phylum | Control | 100 | 18 | 0 | 28.521 | 28.055 |
| *Bacteroidota* | Phylum | Intervention | 100 | 18 | 0 | 28.800 | 26.112 |
| *Campylobacterota* | Phylum | Control | 83 | 15 | 3 | 0.010 | 0.007 |
| *Campylobacterota* | Phylum | Intervention | 89 | 16 | 2 | 0.011 | 0.010 |
| *Desulfobacterota* | Phylum | Control | 78 | 14 | 4 | 0.201 | 0.113 |
| *Desulfobacterota* | Phylum | Intervention | 78 | 14 | 4 | 0.130 | 0.080 |

| | | | | | | | |
|---|---|---|---|---|---|---|---|
| *Pseudomonadota* | Phylum | Control | 100 | 18 | 0 | 2.339 | 1.905 |
| *Pseudomonadota* | Phylum | Intervention | 100 | 18 | 0 | 1.747 | 1.302 |
| *Verrucomicrobiota* | Phylum | Control | 56 | 10 | 8 | 0.097 | 0.013 |
| *Verrucomicrobiota* | Phylum | Intervention | 50 | 9 | 9 | 0.128 | 0.002 |
| *Actinomycetes* | Class | Control | 89 | 16 | 2 | 0.505 | 0.532 |
| *Actinomycetes* | Class | Intervention | 94 | 17 | 1 | 0.477 | 0.444 |
| *Bacilli* | Class | Control | 100 | 18 | 0 | 0.514 | 0.294 |
| *Bacilli* | Class | Intervention | 100 | 18 | 0 | 0.373 | 0.374 |
| *Bacilli_A* | Class | Control | 100 | 18 | 0 | 1.971 | 0.361 |
| *Bacilli_A* | Class | Intervention | 100 | 18 | 0 | 0.591 | 0.466 |
| *Bacteroidia* | Class | Control | 100 | 18 | 0 | 28.521 | 28.055 |
| *Bacteroidia* | Class | Intervention | 100 | 18 | 0 | 28.800 | 26.112 |
| *Campylobacteria* | Class | Control | 83 | 15 | 3 | 0.010 | 0.007 |
| *Campylobacteria* | Class | Intervention | 89 | 16 | 2 | 0.011 | 0.010 |
| *Clostridia* | Class | Control | 100 | 18 | 0 | 60.759 | 58.866 |
| *Clostridia* | Class | Intervention | 100 | 18 | 0 | 61.864 | 64.027 |
| *Coriobacteriia* | Class | Control | 100 | 18 | 0 | 1.502 | 1.080 |
| *Coriobacteriia* | Class | Intervention | 100 | 18 | 0 | 1.561 | 1.494 |
| *Desulfovibrionia* | Class | Control | 78 | 14 | 4 | 0.201 | 0.113 |
| *Desulfovibrionia* | Class | Intervention | 78 | 14 | 4 | 0.130 | 0.080 |
| *Gammaproteobacteria* | Class | Control | 100 | 18 | 0 | 1.739 | 1.322 |
| *Gammaproteobacteria* | Class | Intervention | 100 | 18 | 0 | 1.290 | 1.096 |
| *Negativicutes* | Class | Control | 100 | 18 | 0 | 2.422 | 1.774 |
| *Negativicutes* | Class | Intervention | 100 | 18 | 0 | 3.185 | 2.522 |
| *Peptococcia* | Class | Control | 61 | 11 | 7 | 0.054 | 0.007 |
| *Peptococcia* | Class | Intervention | 89 | 16 | 2 | 0.128 | 0.082 |
| *Actinomycetales* | Order | Control | 89 | 16 | 2 | 0.505 | 0.532 |
| *Actinomycetales* | Order | Intervention | 94 | 17 | 1 | 0.477 | 0.444 |
| *Bacteroidales* | Order | Control | 100 | 18 | 0 | 28.521 | 28.055 |
| *Bacteroidales* | Order | Intervention | 100 | 18 | 0 | 28.782 | 26.112 |
| *Burkholderiales* | Order | Control | 94 | 17 | 1 | 0.928 | 1.024 |
| *Burkholderiales* | Order | Intervention | 100 | 18 | 0 | 0.893 | 0.862 |
| *Campylobacterales* | Order | Control | 83 | 15 | 3 | 0.010 | 0.007 |
| *Campylobacterales* | Order | Intervention | 89 | 16 | 2 | 0.011 | 0.010 |
| *Christensenellales* | Order | Control | 100 | 18 | 0 | 0.906 | 0.542 |
| *Christensenellales* | Order | Intervention | 100 | 18 | 0 | 1.299 | 0.902 |
| *Clostridiales* | Order | Control | 78 | 14 | 4 | 0.143 | 0.046 |
| *Clostridiales* | Order | Intervention | 89 | 16 | 2 | 0.257 | 0.031 |
| *Coriobacteriales* | Order | Control | 100 | 18 | 0 | 1.502 | 1.080 |
| *Coriobacteriales* | Order | Intervention | 100 | 18 | 0 | 1.561 | 1.494 |
| *Desulfovibrionales* | Order | Control | 78 | 14 | 4 | 0.201 | 0.113 |
| *Desulfovibrionales* | Order | Intervention | 78 | 14 | 4 | 0.130 | 0.080 |
| *Enterobacterales* | Order | Control | 100 | 18 | 0 | 0.812 | 0.318 |
| *Enterobacterales* | Order | Intervention | 100 | 18 | 0 | 0.397 | 0.087 |
| *Erysipelotrichales* | Order | Control | 100 | 18 | 0 | 0.174 | 0.098 |
| *Erysipelotrichales* | Order | Intervention | 100 | 18 | 0 | 0.441 | 0.329 |
| *Lachnospirales* | Order | Control | 100 | 18 | 0 | 33.109 | 31.306 |
| *Lachnospirales* | Order | Intervention | 100 | 18 | 0 | 32.540 | 32.435 |
| *Lactobacillales* | Order | Control | 100 | 18 | 0 | 0.436 | 0.225 |
| *Lactobacillales* | Order | Intervention | 100 | 18 | 0 | 0.362 | 0.358 |
| *MOL361* | Order | Control | 83 | 15 | 3 | 0.075 | 0.039 |
| *MOL361* | Order | Intervention | 72 | 13 | 5 | 0.009 | 0.005 |
| *Oscillospirales* | Order | Control | 100 | 18 | 0 | 23.947 | 24.658 |
| *Oscillospirales* | Order | Intervention | 100 | 18 | 0 | 25.454 | 25.292 |
| *Peptococcales* | Order | Control | 61 | 11 | 7 | 0.054 | 0.007 |
| *Peptococcales* | Order | Intervention | 89 | 16 | 2 | 0.128 | 0.082 |
| *Peptostreptococcales* | Order | Control | 100 | 18 | 0 | 0.482 | 0.231 |
| *Peptostreptococcales* | Order | Intervention | 100 | 18 | 0 | 0.279 | 0.235 |
| *RF39* | Order | Control | 33 | 6 | 12 | 0.316 | 0.000 |
| *RF39* | Order | Intervention | 78 | 14 | 4 | 0.104 | 0.053 |
| *TANB77* | Order | Control | 89 | 16 | 2 | 1.635 | 0.543 |
| *TANB77* | Order | Intervention | 94 | 17 | 1 | 1.468 | 0.929 |
| *UBA1381* | Order | Control | 89 | 16 | 2 | 0.191 | 0.130 |
| *UBA1381* | Order | Intervention | 100 | 18 | 0 | 0.327 | 0.340 |
| *Veillonellales* | Order | Control | 100 | 18 | 0 | 2.183 | 1.774 |

| | | | | | | | |
|---|---|---|---|---|---|---|---|
| *Veillonellales* | Order | Intervention | 100 | 18 | 0 | 3.035 | 2.463 |
| *Acutalibacteraceae* | Family | Control | 100 | 18 | 0 | 2.020 | 2.174 |
| *Acutalibacteraceae* | Family | Intervention | 100 | 18 | 0 | 2.852 | 3.078 |
| *Anaerotignaceae* | Family | Control | 100 | 18 | 0 | 0.158 | 0.065 |
| *Anaerotignaceae* | Family | Intervention | 100 | 18 | 0 | 0.132 | 0.102 |
| *Anaerovoracaceae* | Family | Control | 94 | 17 | 1 | 0.108 | 0.075 |
| *Anaerovoracaceae* | Family | Intervention | 100 | 18 | 0 | 0.137 | 0.111 |
| *Aristaeellaceae* | Family | Control | 100 | 18 | 0 | 0.711 | 0.424 |
| *Aristaeellaceae* | Family | Intervention | 100 | 18 | 0 | 1.031 | 0.632 |
| *Bacteroidaceae* | Family | Control | 100 | 18 | 0 | 19.571 | 17.217 |
| *Bacteroidaceae* | Family | Intervention | 100 | 18 | 0 | 21.279 | 18.276 |
| *Barnesiellaceae* | Family | Control | 61 | 11 | 7 | 0.601 | 0.213 |
| *Barnesiellaceae* | Family | Intervention | 94 | 17 | 1 | 0.908 | 0.595 |
| *Bifidobacteriaceae* | Family | Control | 89 | 16 | 2 | 0.503 | 0.527 |
| *Bifidobacteriaceae* | Family | Intervention | 94 | 17 | 1 | 0.475 | 0.438 |
| *Burkholderiaceae* | Family | Control | 94 | 17 | 1 | 0.928 | 1.024 |
| *Burkholderiaceae* | Family | Intervention | 100 | 18 | 0 | 0.893 | 0.862 |
| *Butyricicoccaceae* | Family | Control | 100 | 18 | 0 | 0.402 | 0.335 |
| *Butyricicoccaceae* | Family | Intervention | 100 | 18 | 0 | 0.625 | 0.552 |
| *CAG-508* | Family | Control | 89 | 16 | 2 | 1.626 | 0.543 |
| *CAG-508* | Family | Intervention | 94 | 17 | 1 | 1.468 | 0.928 |
| *Christensenellaceae* | Family | Control | 61 | 11 | 7 | 0.005 | 0.004 |
| *Christensenellaceae* | Family | Intervention | 89 | 16 | 2 | 0.021 | 0.014 |
| *Clostridiaceae* | Family | Control | 78 | 14 | 4 | 0.143 | 0.046 |
| *Clostridiaceae* | Family | Intervention | 89 | 16 | 2 | 0.257 | 0.031 |
| *Coprobacillaceae* | Family | Control | 89 | 16 | 2 | 0.041 | 0.022 |
| *Coprobacillaceae* | Family | Intervention | 100 | 18 | 0 | 0.141 | 0.101 |
| *Coprobacteraceae* | Family | Control | 67 | 12 | 6 | 0.090 | 0.024 |
| *Coprobacteraceae* | Family | Intervention | 56 | 10 | 8 | 0.013 | 0.003 |
| *Coriobacteriaceae* | Family | Control | 94 | 17 | 1 | 1.162 | 0.811 |
| *Coriobacteriaceae* | Family | Intervention | 94 | 17 | 1 | 1.078 | 0.997 |
| *Desulfovibrionaceae* | Family | Control | 78 | 14 | 4 | 0.201 | 0.113 |
| *Desulfovibrionaceae* | Family | Intervention | 78 | 14 | 4 | 0.130 | 0.080 |
| *Dialisteraceae* | Family | Control | 94 | 17 | 1 | 1.878 | 0.631 |
| *Dialisteraceae* | Family | Intervention | 100 | 18 | 0 | 2.990 | 2.441 |
| *Eggerthellaceae* | Family | Control | 100 | 18 | 0 | 0.317 | 0.262 |
| *Eggerthellaceae* | Family | Intervention | 100 | 18 | 0 | 0.456 | 0.433 |
| *Enterobacteriaceae* | Family | Control | 78 | 14 | 4 | 0.050 | 0.026 |
| *Enterobacteriaceae* | Family | Intervention | 78 | 14 | 4 | 0.031 | 0.012 |
| *Erysipelotrichaceae* | Family | Control | 100 | 18 | 0 | 0.133 | 0.057 |
| *Erysipelotrichaceae* | Family | Intervention | 100 | 18 | 0 | 0.300 | 0.089 |
| *Lachnospiraceae* | Family | Control | 100 | 18 | 0 | 32.931 | 31.118 |
| *Lachnospiraceae* | Family | Intervention | 100 | 18 | 0 | 32.404 | 32.328 |
| *Marinifilaceae* | Family | Control | 83 | 15 | 3 | 0.336 | 0.303 |
| *Marinifilaceae* | Family | Intervention | 100 | 18 | 0 | 0.336 | 0.226 |
| *Muribaculaceae* | Family | Control | 78 | 14 | 4 | 0.347 | 0.009 |
| *Muribaculaceae* | Family | Intervention | 83 | 15 | 3 | 0.293 | 0.023 |
| *Oscillospiraceae* | Family | Control | 100 | 18 | 0 | 1.996 | 1.742 |
| *Oscillospiraceae* | Family | Intervention | 100 | 18 | 0 | 2.990 | 3.253 |
| *Pasteurellaceae* | Family | Control | 100 | 18 | 0 | 0.752 | 0.288 |
| *Pasteurellaceae* | Family | Intervention | 94 | 17 | 1 | 0.366 | 0.055 |
| *Peptococcaceae* | Family | Control | 61 | 11 | 7 | 0.054 | 0.007 |
| *Peptococcaceae* | Family | Intervention | 89 | 16 | 2 | 0.128 | 0.082 |
| *Peptostreptococcaceae* | Family | Control | 94 | 17 | 1 | 0.374 | 0.104 |
| *Peptostreptococcaceae* | Family | Intervention | 100 | 18 | 0 | 0.131 | 0.085 |
| *Rikenellaceae* | Family | Control | 100 | 18 | 0 | 4.516 | 3.548 |
| *Rikenellaceae* | Family | Intervention | 100 | 18 | 0 | 3.489 | 3.143 |
| *Ruminococcaceae* | Family | Control | 100 | 18 | 0 | 18.798 | 18.524 |
| *Ruminococcaceae* | Family | Intervention | 100 | 18 | 0 | 17.916 | 17.943 |
| *Streptococcaceae* | Family | Control | 100 | 18 | 0 | 0.421 | 0.212 |
| *Streptococcaceae* | Family | Intervention | 100 | 18 | 0 | 0.289 | 0.278 |
| *Sulfurovaceae* | Family | Control | 83 | 15 | 3 | 0.010 | 0.007 |
| *Sulfurovaceae* | Family | Intervention | 89 | 16 | 2 | 0.011 | 0.010 |
| *Tannerellaceae* | Family | Control | 100 | 18 | 0 | 2.428 | 1.956 |
| *Tannerellaceae* | Family | Intervention | 100 | 18 | 0 | 1.902 | 1.375 |

| | | | | | | | |
|---|---|---|---|---|---|---|---|
| *Turicibacteraceae* | Family | Control | 83 | 15 | 3 | 0.075 | 0.039 |
| *Turicibacteraceae* | Family | Intervention | 72 | 13 | 5 | 0.009 | 0.005 |
| *UBA1381* | Family | Control | 89 | 16 | 2 | 0.191 | 0.130 |
| *UBA1381* | Family | Intervention | 100 | 18 | 0 | 0.324 | 0.340 |
| *UBA660* | Family | Control | 33 | 6 | 12 | 0.316 | 0.000 |
| *UBA660* | Family | Intervention | 78 | 14 | 4 | 0.104 | 0.053 |
| *Veillonellaceae* | Family | Control | 100 | 18 | 0 | 0.305 | 0.056 |
| *Veillonellaceae* | Family | Intervention | 94 | 17 | 1 | 0.045 | 0.037 |
| *Acetatifactor* | Genus | Control | 100 | 18 | 0 | 1.015 | 0.513 |
| *Acetatifactor* | Genus | Intervention | 100 | 18 | 0 | 1.142 | 0.874 |
| *Adlercreutzia* | Genus | Control | 94 | 17 | 1 | 0.099 | 0.037 |
| *Adlercreutzia* | Genus | Intervention | 94 | 17 | 1 | 0.123 | 0.107 |
| *Agathobacter* | Genus | Control | 100 | 18 | 0 | 9.275 | 7.712 |
| *Agathobacter* | Genus | Intervention | 100 | 18 | 0 | 6.898 | 7.232 |
| *Agathobaculum* | Genus | Control | 100 | 18 | 0 | 0.392 | 0.310 |
| *Agathobaculum* | Genus | Intervention | 100 | 18 | 0 | 0.590 | 0.471 |
| *Alistipes* | Genus | Control | 100 | 18 | 0 | 4.332 | 3.287 |
| *Alistipes* | Genus | Intervention | 100 | 18 | 0 | 3.278 | 2.887 |
| *Alistipes_A* | Genus | Control | 89 | 16 | 2 | 0.074 | 0.071 |
| *Alistipes_A* | Genus | Intervention | 100 | 18 | 0 | 0.159 | 0.075 |
| *Alitiscatomonas* | Genus | Control | 100 | 18 | 0 | 0.409 | 0.335 |
| *Alitiscatomonas* | Genus | Intervention | 100 | 18 | 0 | 0.379 | 0.422 |
| *Alloprevotella* | Genus | Control | 67 | 12 | 6 | 0.164 | 0.016 |
| *Alloprevotella* | Genus | Intervention | 61 | 11 | 7 | 0.514 | 0.020 |
| *Anaerobutyricum* | Genus | Control | 100 | 18 | 0 | 0.857 | 0.741 |
| *Anaerobutyricum* | Genus | Intervention | 100 | 18 | 0 | 1.542 | 1.622 |
| *Anaerostipes* | Genus | Control | 100 | 18 | 0 | 2.543 | 1.425 |
| *Anaerostipes* | Genus | Intervention | 100 | 18 | 0 | 1.918 | 1.623 |
| *Anaerotardibacter* | Genus | Control | 61 | 11 | 7 | 0.171 | 0.053 |
| *Anaerotardibacter* | Genus | Intervention | 100 | 18 | 0 | 0.292 | 0.230 |
| *Anaerotignum* | Genus | Control | 72 | 13 | 5 | 0.120 | 0.031 |
| *Anaerotignum* | Genus | Intervention | 94 | 17 | 1 | 0.096 | 0.063 |
| *Anthropogastromicrobium* | Genus | Control | 94 | 17 | 1 | 0.325 | 0.290 |
| *Anthropogastromicrobium* | Genus | Intervention | 100 | 18 | 0 | 0.534 | 0.418 |
| *Aristaeella* | Genus | Control | 33 | 6 | 12 | 0.022 | 0.000 |
| *Aristaeella* | Genus | Intervention | 67 | 12 | 6 | 0.025 | 0.028 |
| *Bacteroides* | Genus | Control | 100 | 18 | 0 | 8.560 | 4.960 |
| *Bacteroides* | Genus | Intervention | 100 | 18 | 0 | 7.380 | 4.143 |
| *Bariatricus* | Genus | Control | 100 | 18 | 0 | 0.581 | 0.520 |
| *Bariatricus* | Genus | Intervention | 100 | 18 | 0 | 0.350 | 0.295 |
| *Barnesiella* | Genus | Control | 61 | 11 | 7 | 0.601 | 0.213 |
| *Barnesiella* | Genus | Intervention | 94 | 17 | 1 | 0.908 | 0.595 |
| *Bifidobacterium* | Genus | Control | 89 | 16 | 2 | 0.503 | 0.527 |
| *Bifidobacterium* | Genus | Intervention | 94 | 17 | 1 | 0.475 | 0.438 |
| *Bilophila* | Genus | Control | 78 | 14 | 4 | 0.154 | 0.106 |
| *Bilophila* | Genus | Intervention | 78 | 14 | 4 | 0.073 | 0.063 |
| *Blautia_A* | Genus | Control | 100 | 18 | 0 | 2.620 | 2.325 |
| *Blautia_A* | Genus | Intervention | 100 | 18 | 0 | 3.544 | 3.893 |
| *Brotaphodocola* | Genus | Control | 100 | 18 | 0 | 0.236 | 0.204 |
| *Brotaphodocola* | Genus | Intervention | 100 | 18 | 0 | 0.174 | 0.150 |
| *Butyribacter* | Genus | Control | 89 | 16 | 2 | 0.413 | 0.294 |
| *Butyribacter* | Genus | Intervention | 100 | 18 | 0 | 0.762 | 0.646 |
| *Butyricimonas* | Genus | Control | 67 | 12 | 6 | 0.254 | 0.208 |
| *Butyricimonas* | Genus | Intervention | 67 | 12 | 6 | 0.164 | 0.126 |
| *CAG-170* | Genus | Control | 67 | 12 | 6 | 0.183 | 0.074 |
| *CAG-170* | Genus | Intervention | 100 | 18 | 0 | 0.279 | 0.239 |
| *CAG-177* | Genus | Control | 33 | 6 | 12 | 0.220 | 0.000 |
| *CAG-177* | Genus | Intervention | 61 | 11 | 7 | 0.295 | 0.050 |
| *CAG-238* | Genus | Control | 61 | 11 | 7 | 0.034 | 0.018 |
| *CAG-238* | Genus | Intervention | 72 | 13 | 5 | 0.025 | 0.023 |
| *CAG-269* | Genus | Control | 78 | 14 | 4 | 0.639 | 0.110 |
| *CAG-269* | Genus | Intervention | 94 | 17 | 1 | 0.724 | 0.671 |
| *Choladousia* | Genus | Control | 100 | 18 | 0 | 0.047 | 0.036 |
| *Choladousia* | Genus | Intervention | 100 | 18 | 0 | 0.049 | 0.047 |
| *Clostridium* | Genus | Control | 78 | 14 | 4 | 0.143 | 0.046 |

| | | | | | | | |
|---|---|---|---|---|---|---|---|
| *Clostridium* | Genus | Intervention | 89 | 16 | 2 | 0.257 | 0.031 |
| *Collinsella* | Genus | Control | 94 | 17 | 1 | 1.162 | 0.811 |
| *Collinsella* | Genus | Intervention | 94 | 17 | 1 | 1.078 | 0.997 |
| *Coprobacter* | Genus | Control | 67 | 12 | 6 | 0.090 | 0.024 |
| *Coprobacter* | Genus | Intervention | 56 | 10 | 8 | 0.013 | 0.003 |
| *Coprococcus* | Genus | Control | 89 | 16 | 2 | 0.848 | 0.349 |
| *Coprococcus* | Genus | Intervention | 83 | 15 | 3 | 1.993 | 2.085 |
| *Coprococcus_A* | Genus | Control | 100 | 18 | 0 | 0.148 | 0.154 |
| *Coprococcus_A* | Genus | Intervention | 89 | 16 | 2 | 0.160 | 0.142 |
| *Dialister* | Genus | Control | 94 | 17 | 1 | 1.871 | 0.631 |
| *Dialister* | Genus | Intervention | 100 | 18 | 0 | 2.967 | 2.441 |
| *Dorea* | Genus | Control | 100 | 18 | 0 | 0.176 | 0.102 |
| *Dorea* | Genus | Intervention | 100 | 18 | 0 | 0.232 | 0.233 |
| *Dorea_A* | Genus | Control | 100 | 18 | 0 | 0.915 | 0.796 |
| *Dorea_A* | Genus | Intervention | 100 | 18 | 0 | 0.784 | 0.765 |
| *Dorea_D* | Genus | Control | 89 | 16 | 2 | 0.045 | 0.029 |
| *Dorea_D* | Genus | Intervention | 100 | 18 | 0 | 0.121 | 0.098 |
| *Dysosmobacter* | Genus | Control | 100 | 18 | 0 | 0.287 | 0.281 |
| *Dysosmobacter* | Genus | Intervention | 100 | 18 | 0 | 0.499 | 0.462 |
| *Enterocloster* | Genus | Control | 100 | 18 | 0 | 0.311 | 0.282 |
| *Enterocloster* | Genus | Intervention | 100 | 18 | 0 | 0.261 | 0.218 |
| *Eubacterium_F* | Genus | Control | 72 | 13 | 5 | 0.416 | 0.202 |
| *Eubacterium_F* | Genus | Intervention | 100 | 18 | 0 | 0.821 | 0.620 |
| *Eubacterium_G* | Genus | Control | 100 | 18 | 0 | 0.435 | 0.304 |
| *Eubacterium_G* | Genus | Intervention | 100 | 18 | 0 | 0.337 | 0.315 |
| *Eubacterium_I* | Genus | Control | 78 | 14 | 4 | 0.080 | 0.042 |
| *Eubacterium_I* | Genus | Intervention | 100 | 18 | 0 | 0.070 | 0.044 |
| *Eubacterium_R* | Genus | Control | 67 | 12 | 6 | 0.229 | 0.177 |
| *Eubacterium_R* | Genus | Intervention | 83 | 15 | 3 | 0.583 | 0.371 |
| *Evtepia* | Genus | Control | 67 | 12 | 6 | 0.053 | 0.038 |
| *Evtepia* | Genus | Intervention | 83 | 15 | 3 | 0.091 | 0.104 |
| *Faecalibacillus* | Genus | Control | 56 | 10 | 8 | 0.028 | 0.003 |
| *Faecalibacillus* | Genus | Intervention | 61 | 11 | 7 | 0.109 | 0.045 |
| *Faecalibacterium* | Genus | Control | 100 | 18 | 0 | 13.631 | 13.239 |
| *Faecalibacterium* | Genus | Intervention | 100 | 18 | 0 | 12.609 | 12.423 |
| *Faecousia* | Genus | Control | 61 | 11 | 7 | 0.146 | 0.037 |
| *Faecousia* | Genus | Intervention | 94 | 17 | 1 | 0.206 | 0.158 |
| *Fusicatenibacter* | Genus | Control | 100 | 18 | 0 | 1.392 | 1.203 |
| *Fusicatenibacter* | Genus | Intervention | 100 | 18 | 0 | 1.203 | 1.168 |
| *Gallintestinimicrobium* | Genus | Control | 100 | 18 | 0 | 0.292 | 0.239 |
| *Gallintestinimicrobium* | Genus | Intervention | 100 | 18 | 0 | 0.365 | 0.309 |
| *Gemmiger* | Genus | Control | 100 | 18 | 0 | 2.947 | 2.770 |
| *Gemmiger* | Genus | Intervention | 100 | 18 | 0 | 2.905 | 2.504 |
| *Haemophilus_D* | Genus | Control | 100 | 18 | 0 | 0.743 | 0.286 |
| *Haemophilus_D* | Genus | Intervention | 94 | 17 | 1 | 0.360 | 0.054 |
| *Hominenteromicrobium* | Genus | Control | 89 | 16 | 2 | 0.256 | 0.188 |
| *Hominenteromicrobium* | Genus | Intervention | 100 | 18 | 0 | 0.241 | 0.168 |
| *Hominicoprocola* | Genus | Control | 94 | 17 | 1 | 0.476 | 0.384 |
| *Hominicoprocola* | Genus | Intervention | 100 | 18 | 0 | 0.696 | 0.577 |
| *Hominilimicola* | Genus | Control | 89 | 16 | 2 | 0.191 | 0.130 |
| *Hominilimicola* | Genus | Intervention | 100 | 18 | 0 | 0.324 | 0.340 |
| *Hominimerdicola* | Genus | Control | 78 | 14 | 4 | 0.991 | 0.716 |
| *Hominimerdicola* | Genus | Intervention | 100 | 18 | 0 | 0.761 | 0.462 |
| *Hominiventricola* | Genus | Control | 100 | 18 | 0 | 0.309 | 0.248 |
| *Hominiventricola* | Genus | Intervention | 100 | 18 | 0 | 0.396 | 0.371 |
| *Intestinibacter* | Genus | Control | 78 | 14 | 4 | 0.187 | 0.012 |
| *Intestinibacter* | Genus | Intervention | 67 | 12 | 6 | 0.038 | 0.012 |
| *JAGTTR01* | Genus | Control | 83 | 15 | 3 | 0.018 | 0.016 |
| *JAGTTR01* | Genus | Intervention | 83 | 15 | 3 | 0.022 | 0.018 |
| *Lachnoclostridium_B* | Genus | Control | 100 | 18 | 0 | 0.057 | 0.032 |
| *Lachnoclostridium_B* | Genus | Intervention | 100 | 18 | 0 | 0.062 | 0.047 |
| *Lachnospira* | Genus | Control | 100 | 18 | 0 | 2.764 | 1.923 |
| *Lachnospira* | Genus | Intervention | 100 | 18 | 0 | 2.211 | 1.857 |
| *Laedolimicola* | Genus | Control | 94 | 17 | 1 | 0.063 | 0.053 |
| *Laedolimicola* | Genus | Intervention | 100 | 18 | 0 | 0.092 | 0.094 |

| | | | | | | | |
|---|---|---|---|---|---|---|---|
| *Lawsonibacter* | Genus | Control | 100 | 18 | 0 | 0.117 | 0.103 |
| *Lawsonibacter* | Genus | Intervention | 100 | 18 | 0 | 0.085 | 0.077 |
| *Lentihominibacter* | Genus | Control | 89 | 16 | 2 | 0.035 | 0.023 |
| *Lentihominibacter* | Genus | Intervention | 100 | 18 | 0 | 0.047 | 0.044 |
| *Limivivens* | Genus | Control | 100 | 18 | 0 | 0.103 | 0.092 |
| *Limivivens* | Genus | Intervention | 100 | 18 | 0 | 0.138 | 0.133 |
| *Mediterraneibacter* | Genus | Control | 100 | 18 | 0 | 0.482 | 0.413 |
| *Mediterraneibacter* | Genus | Intervention | 100 | 18 | 0 | 0.734 | 0.483 |
| *Mediterraneibacter_A* | Genus | Control | 89 | 16 | 2 | 0.013 | 0.012 |
| *Mediterraneibacter_A* | Genus | Intervention | 100 | 18 | 0 | 0.018 | 0.018 |
| *Muricoprocola* | Genus | Control | 44 | 8 | 10 | 0.014 | 0.000 |
| *Muricoprocola* | Genus | Intervention | 61 | 11 | 7 | 0.041 | 0.022 |
| *Odoribacter* | Genus | Control | 67 | 12 | 6 | 0.082 | 0.005 |
| *Odoribacter* | Genus | Intervention | 78 | 14 | 4 | 0.172 | 0.010 |
| *Oliverpabstia* | Genus | Control | 94 | 17 | 1 | 0.252 | 0.230 |
| *Oliverpabstia* | Genus | Intervention | 89 | 16 | 2 | 0.285 | 0.216 |
| *Onthenecus* | Genus | Control | 67 | 12 | 6 | 0.110 | 0.004 |
| *Onthenecus* | Genus | Intervention | 83 | 15 | 3 | 0.163 | 0.022 |
| *Otoolea* | Genus | Control | 100 | 18 | 0 | 0.386 | 0.282 |
| *Otoolea* | Genus | Intervention | 100 | 18 | 0 | 0.454 | 0.342 |
| *Parabacteroides* | Genus | Control | 100 | 18 | 0 | 2.428 | 1.956 |
| *Parabacteroides* | Genus | Intervention | 100 | 18 | 0 | 1.902 | 1.375 |
| *Parasutterella* | Genus | Control | 83 | 15 | 3 | 0.123 | 0.032 |
| *Parasutterella* | Genus | Intervention | 78 | 14 | 4 | 0.148 | 0.026 |
| *Phocaeicola* | Genus | Control | 100 | 18 | 0 | 7.503 | 6.017 |
| *Phocaeicola* | Genus | Intervention | 100 | 18 | 0 | 6.942 | 5.165 |
| *Prevotella* | Genus | Control | 83 | 15 | 3 | 2.892 | 0.009 |
| *Prevotella* | Genus | Intervention | 78 | 14 | 4 | 6.047 | 2.210 |
| *Romboutsia* | Genus | Control | 94 | 17 | 1 | 0.085 | 0.061 |
| *Romboutsia* | Genus | Intervention | 100 | 18 | 0 | 0.071 | 0.039 |
| *Roseburia* | Genus | Control | 100 | 18 | 0 | 1.656 | 0.357 |
| *Roseburia* | Genus | Intervention | 100 | 18 | 0 | 0.959 | 0.722 |
| *Roseburia_C* | Genus | Control | 61 | 11 | 7 | 0.053 | 0.014 |
| *Roseburia_C* | Genus | Intervention | 100 | 18 | 0 | 0.328 | 0.122 |
| *Ruminiclostridium_E* | Genus | Control | 78 | 14 | 4 | 0.464 | 0.036 |
| *Ruminiclostridium_E* | Genus | Intervention | 100 | 18 | 0 | 0.289 | 0.121 |
| *Ruminococcus_C* | Genus | Control | 67 | 12 | 6 | 0.313 | 0.173 |
| *Ruminococcus_C* | Genus | Intervention | 61 | 11 | 7 | 0.385 | 0.002 |
| *Ruminococcus_E* | Genus | Control | 89 | 16 | 2 | 0.820 | 0.309 |
| *Ruminococcus_E* | Genus | Intervention | 94 | 17 | 1 | 1.072 | 0.969 |
| *Ruthenibacterium* | Genus | Control | 100 | 18 | 0 | 0.017 | 0.012 |
| *Ruthenibacterium* | Genus | Intervention | 94 | 17 | 1 | 0.008 | 0.006 |
| *Schaedlerella* | Genus | Control | 83 | 15 | 3 | 0.047 | 0.030 |
| *Schaedlerella* | Genus | Intervention | 72 | 13 | 5 | 0.019 | 0.004 |
| *Streptococcus* | Genus | Control | 100 | 18 | 0 | 0.421 | 0.212 |
| *Streptococcus* | Genus | Intervention | 100 | 18 | 0 | 0.289 | 0.278 |
| *Sulfurovum* | Genus | Control | 83 | 15 | 3 | 0.010 | 0.007 |
| *Sulfurovum* | Genus | Intervention | 89 | 16 | 2 | 0.011 | 0.010 |
| *Sutterella* | Genus | Control | 89 | 16 | 2 | 0.782 | 0.844 |
| *Sutterella* | Genus | Intervention | 83 | 15 | 3 | 0.667 | 0.697 |
| *Tidjanibacter* | Genus | Control | 50 | 9 | 9 | 0.110 | 0.021 |
| *Tidjanibacter* | Genus | Intervention | 44 | 8 | 10 | 0.053 | 0.000 |
| *Turicibacter* | Genus | Control | 83 | 15 | 3 | 0.075 | 0.039 |
| *Turicibacter* | Genus | Intervention | 72 | 13 | 5 | 0.009 | 0.005 |
| *UBA3402* | Genus | Control | 83 | 15 | 3 | 0.073 | 0.056 |
| *UBA3402* | Genus | Intervention | 89 | 16 | 2 | 0.068 | 0.071 |
| *UBA7185* | Genus | Control | 44 | 8 | 10 | 0.040 | 0.000 |
| *UBA7185* | Genus | Intervention | 78 | 14 | 4 | 0.110 | 0.055 |
| *Veillonella* | Genus | Control | 100 | 18 | 0 | 0.305 | 0.056 |
| *Veillonella* | Genus | Intervention | 94 | 17 | 1 | 0.045 | 0.037 |
| *Vescimonas* | Genus | Control | 89 | 16 | 2 | 0.416 | 0.251 |
| *Vescimonas* | Genus | Intervention | 100 | 18 | 0 | 0.631 | 0.457 |
| *Wujia* | Genus | Control | 50 | 9 | 9 | 0.435 | 0.019 |
| *Wujia* | Genus | Intervention | 50 | 9 | 9 | 0.262 | 0.000 |

### 5. DIETARY INTAKE OF SELECTED MICRONUTRIENTS AND FOOD GROUPS

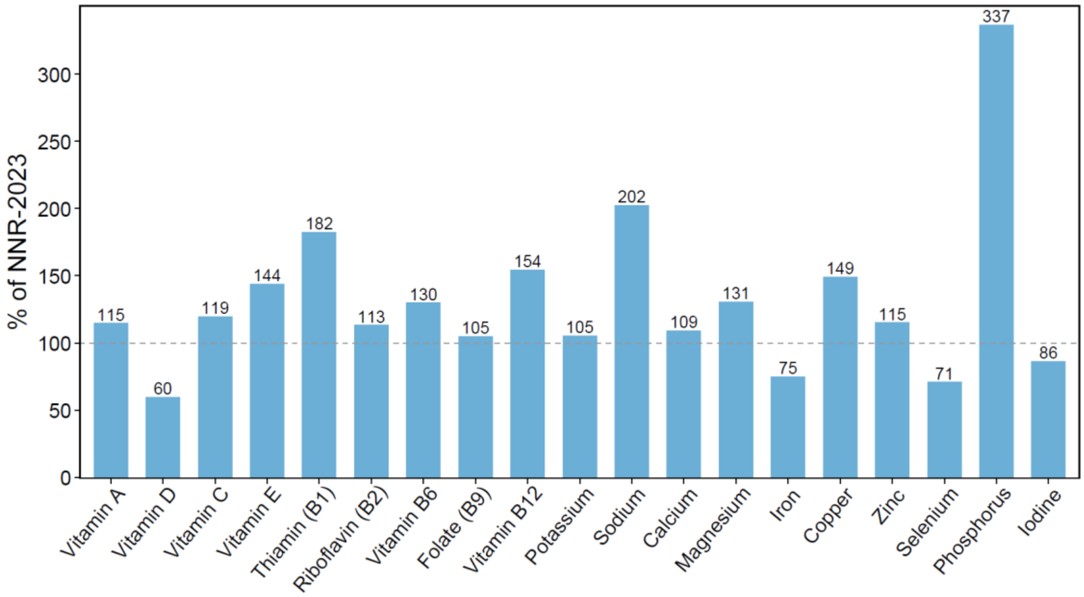

**Supplementary Figure S10**. Dietary intake of selected micronutrients at baseline (n = 23) expressed as percentages of the recommended intakes from NNR-2023 (12). Values represent mean or median percentage, depending on data distribution. For nutrients without established recommended intakes, adequate intake values were used as reference. The dashed grey line indicates 100% of the recommended lower intake values from NNR-2023, with values below this line indicating intakes below the recommendations. For several micronutrients, the recommended intake represents a range with an established upper intake level; thus, intakes exceeding 100% of the recommended value do not necessarily reflect intakes beyond the recommended range. NNR, Nordic Nutrition Recommendations.

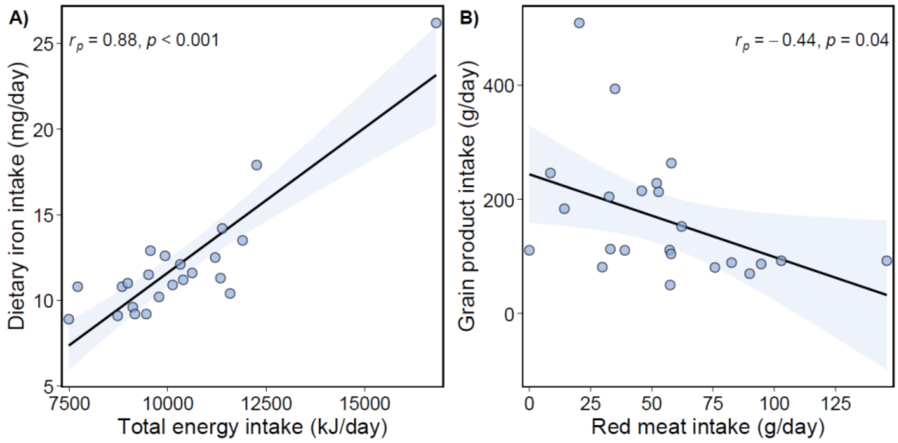

**Supplementary Figure S11**. Scatterplots showing Pearson's correlation at baseline (n = 23) between (A) daily intake of dietary iron (mg/day) and total energy (kJ/day) and (B) daily intake of red meat (g/day) and grain products (g/day). Pearson's correlation coefficients ($r_p$) with corresponding $P$-values are shown in the upper left or right corner. Each dot represents one participant.

**Supplementary Table S12**. Estimated daily intakes of selected food groups at baseline (n = 23).

| Food group (g/day) | Mean (SD) | Median (Q1, Q3) |
|---|---|---|
| Grain products[1] | 345 (155) | 336 (259, 371) |
| Whole grains | 101 (78) | 84 (62, 107) |
| Dietary fiber | 31 (12) | 29 (24, 33) |
| Dairy products[2] | 338 (164) | 335 (238, 454) |
| Fish and shellfish | 59 (47) | 54 (31, 81) |
| Total meat | 106 (44) | 104 (81, 131) |
| Red meat | 54 (34) | 53 (33, 69) |
| White meat | 52 (39) | 43 (28, 66) |
| Total vegetables, fruits and berries[3] | 588 (206) | 572 (410, 739) |
| Vegetables | 207 (70) | 192 (166, 255) |
| Fruits and berries | 381 (180) | 357 (245, 540) |

Estimates are based on digital 7-day food diaries submitted prior to the baseline visit.
[1] Includes bread and other grain products, excluding cake.
[2] Includes milk, yogurt, cultured milk, ice cream and cheese (white, low-fat, brown).
[3] Does not include potatoes.

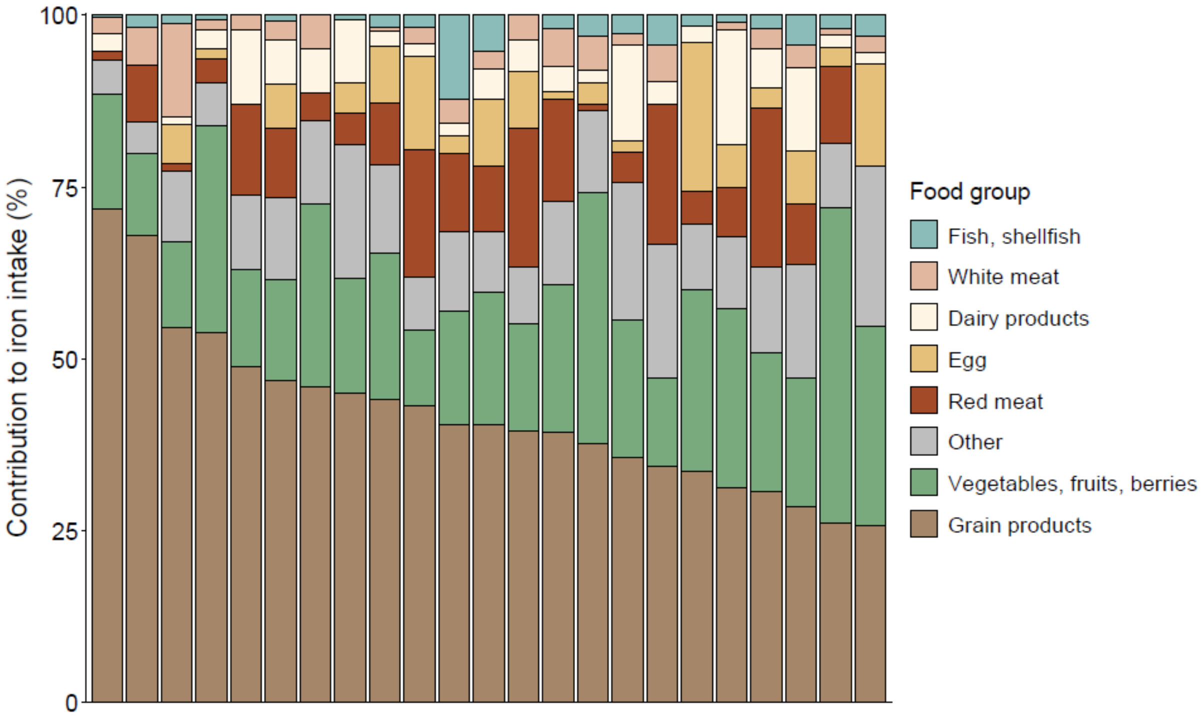

**Supplementary Figure S13**. Contribution of selected food groups to daily dietary iron intake (%) for each participant at baseline (n = 23). Each bar represents one participant, sorted in descending order based on the food group with the largest average contribution (grain products). The "Other" category includes iron from ungrouped foods, cakes, potatoes, butter, margarine, oil, sugar and sweets, beverages, mixed dishes and spices/salt.

### 6. EVALUATION OF THE FOOD DIARY

The serum carotenoids lutein, zeaxanthin, β-cryptoxanthin, β-carotene, α-carotene and lycopene were measured by high-performance liquid chromatography using a previously described method (13). To assess the performance of the web-based food diary in estimating the intake of vegetables, fruits and berries, we correlated reported intakes with serum carotenoid concentrations (**Supplementary Table S14**, **Figures S15–S17**). Total daily intake of vegetables, fruits and berries correlated positively with total serum carotenoid concentration ($r_p$ = 0.45, $P$ = 0.03), with the strongest relationship observed for β-carotene ($r_p$ = 0.47, $P$ = 0.02). In addition, β-carotene correlated positively with intake of fruits and berries ($r_p$ = 0.43, $P$ = 0.04), and α-carotene correlated positively with both vegetable intake ($r_p$ = 0.42, $P$ = 0.05) and total intake of vegetables, fruits and berries ($r_p$ = 0.42, $P$ = 0.04).

**Supplementary Table S14**. Correlations between serum carotenoids and dietary intake of fruits, berries and vegetables at baseline (n = 23).

| Carotenoid (µmol/L) | Fruits and berries (g/day) | | Vegetables (g/day) | | Total fruits, berries and vegetables (g/day) | |
|---|---|---|---|---|---|---|
| | $r_p$ | P-value | $r_p$ | P-value | $r_p$ | P-value |
| α-carotene | 0.32 [–0.11, 0.65] | 0.14 | 0.42 [0.01, 0.71] | **0.05** | 0.42 [0.01, 0.71] | **0.04** |
| β-carotene | 0.43 [0.02, 0.72] | **0.04** | 0.27 [–0.16, 0.61] | 0.22 | 0.47 [0.07, 0.74] | **0.02** |
| β-cryptoxanthin | 0.38 [–0.03, 0.69] | 0.07 | –0.21 [–0.57, 0.23] | 0.35 | 0.27 [–0.16, 0.61] | 0.22 |
| Lycopene | –0.21 [–0.57, 0.22] | 0.33 | 0.12 [–0.31, 0.51] | 0.58 | –0.14 [–0.52, 0.29] | 0.51 |
| Lutein | 0.03 [–0.39, 0.44] | 0.89 | –0.19 [–0.56, 0.25] | 0.40 | –0.04 [–0.44, 0.38] | 0.87 |
| Zeaxanthin | –0.06 [–0.46, 0.36] | 0.77 | –0.47 [–0.74, –0.07] | **0.03** | –0.21 [–0.58, 0.22] | 0.33 |
| Total carotenoids | 0.39 [–0.03, 0.69] | 0.07 | 0.33 [–0.10, 0.65] | 0.12 | 0.45 [0.05, 0.73] | **0.03** |

Values are Pearson's correlation coefficients ($r_p$) with 95% CI and corresponding *P*-values. Statistically significant *P*-values are shown in bold ($P < 0.05$), and cell shading reflects the strength and direction of the relationship (red gradient = positive, blue gradient = negative).

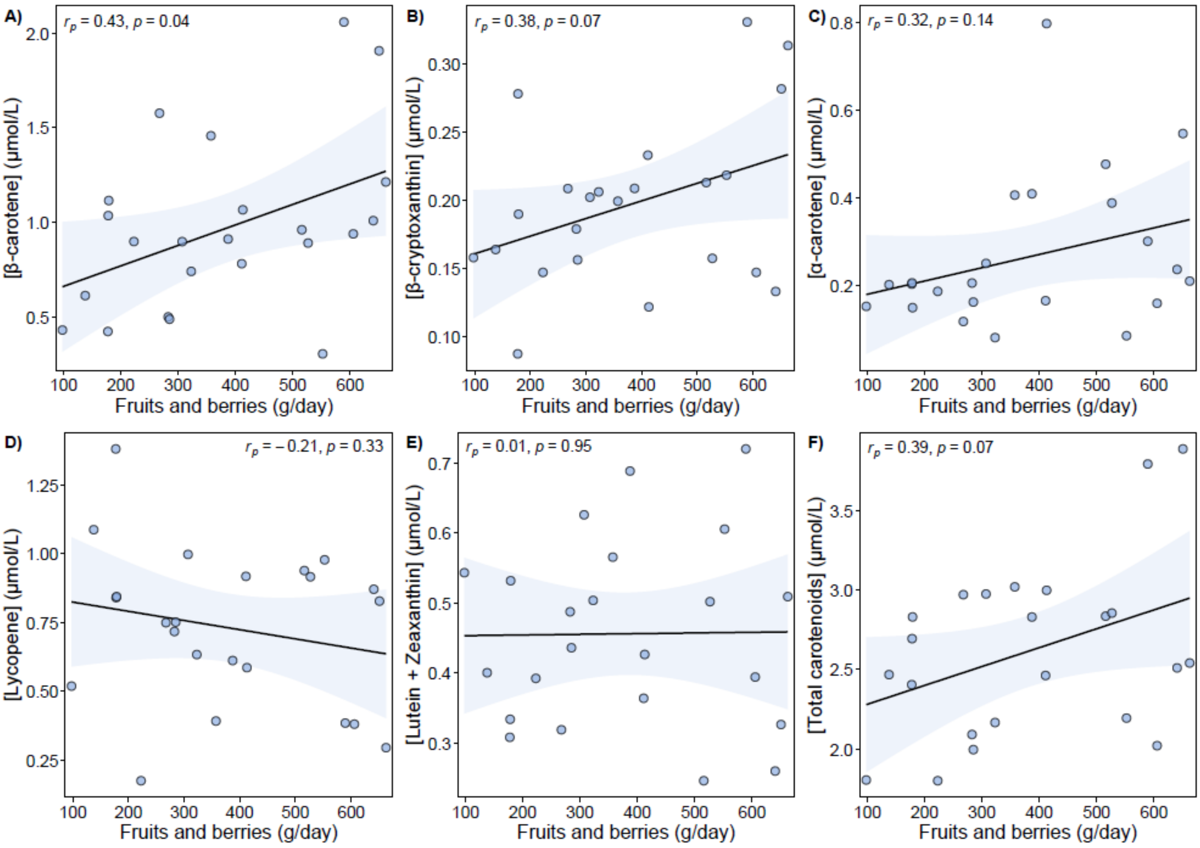

**Supplementary Figure S15**. Scatterplots showing Pearson's correlation at baseline (n = 23) between daily intake of fruits and berries and serum concentrations of (A) β-carotene, (B) β-cryptoxanthin, (C) α-carotene, (D) lycopene, (E) the sum of lutein and zeaxanthin and (F) total carotenoids. Pearson's correlation coefficients ($r_p$) with corresponding $P$-values are shown in the upper left or right corner. Each dot represents one participant.

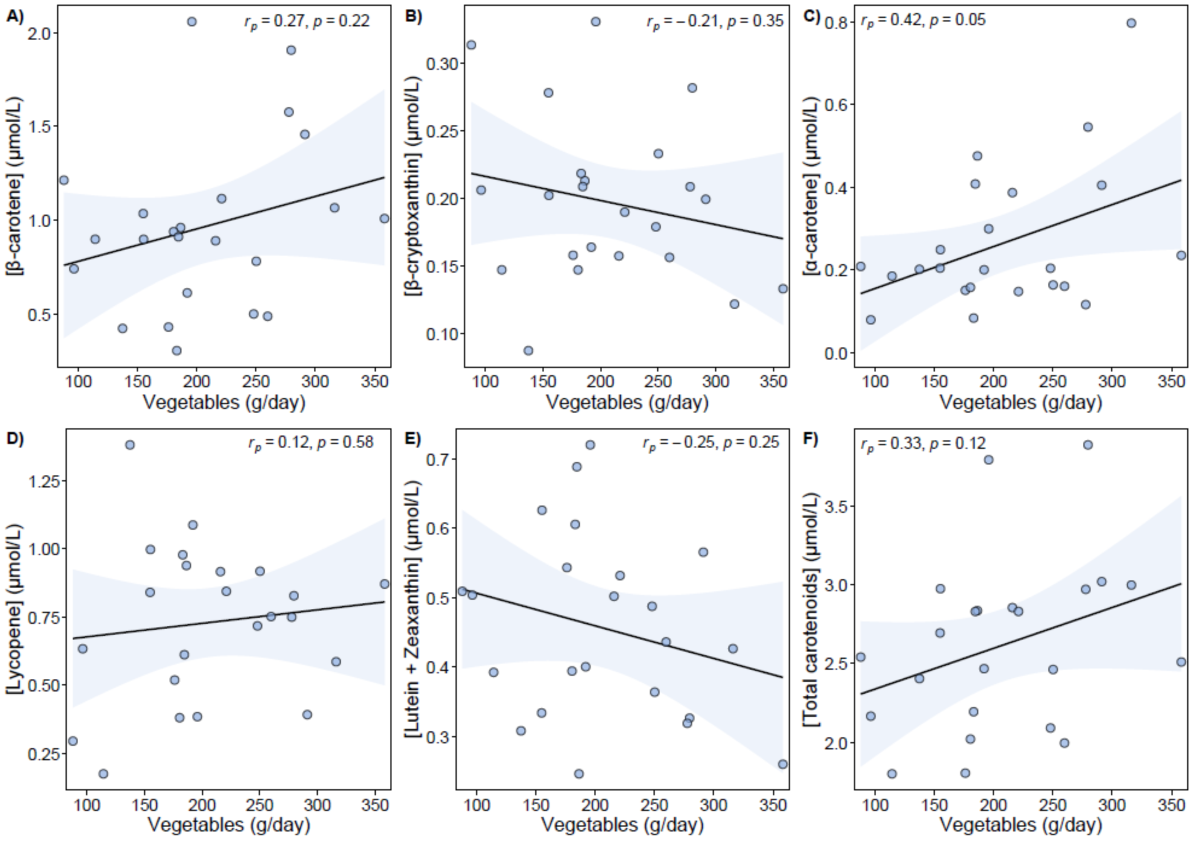

**Supplementary Figure S16**. Scatterplots showing Pearson's correlation at baseline (n = 23) between daily intake of vegetables and serum concentrations of (A) β-carotene, (B) β-cryptoxanthin, (C) α-carotene, (D) lycopene, (E) the sum of lutein and zeaxanthin and (F) total carotenoids. Pearson's correlation coefficients ($r_p$) with corresponding *P*-values are shown in the upper left or right corner. Each dot represents one participant.

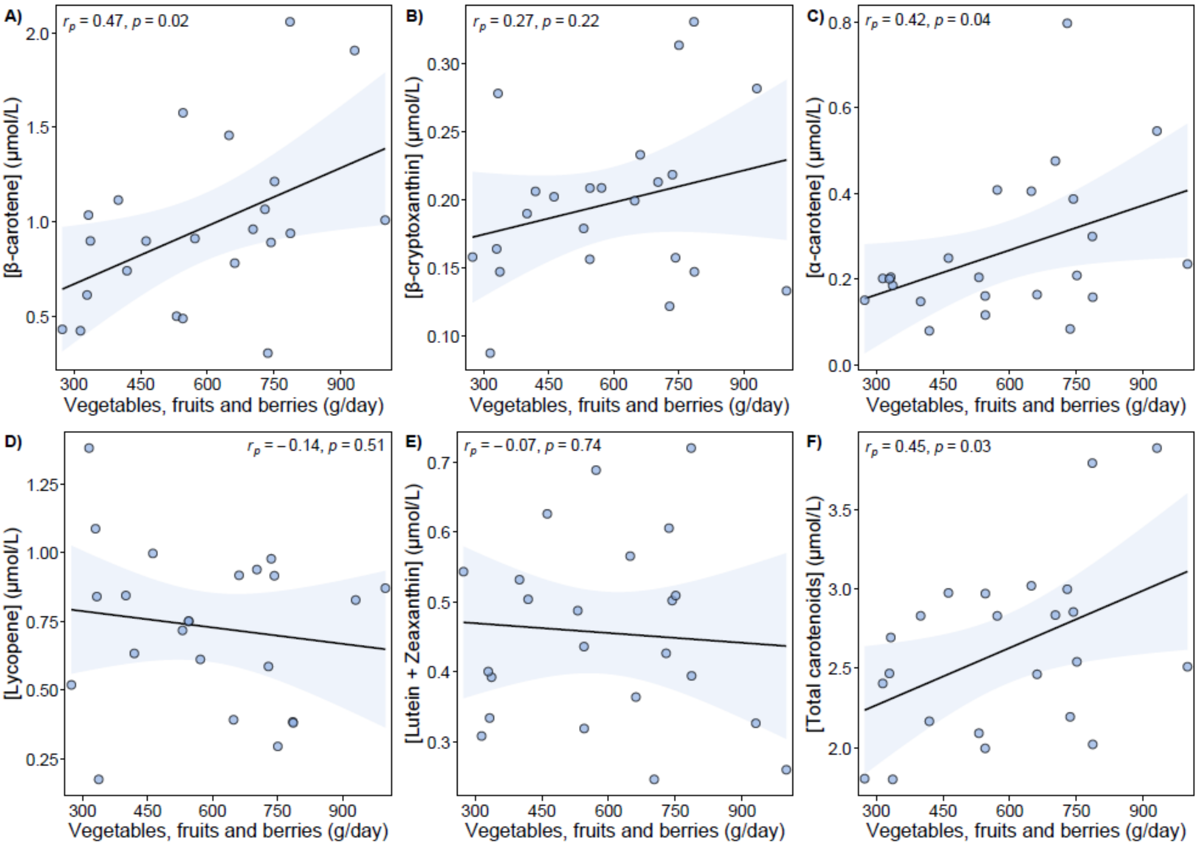

**Supplementary Figure S17**. Scatterplots showing Pearson's correlation at baseline (n = 23) between total daily intake of fruits, berries and vegetables and serum concentrations of (A) β-carotene, B) β-cryptoxanthin, (C) α-carotene, (D) lycopene, (E) the sum of lutein and zeaxanthin and (F) total carotenoids. Pearson's correlation coefficients ($r_p$) with corresponding *P*-values are shown in the upper left or right corner. Each dot represents one participant.

### 7. EFFECTS OF THE INTERVENTION

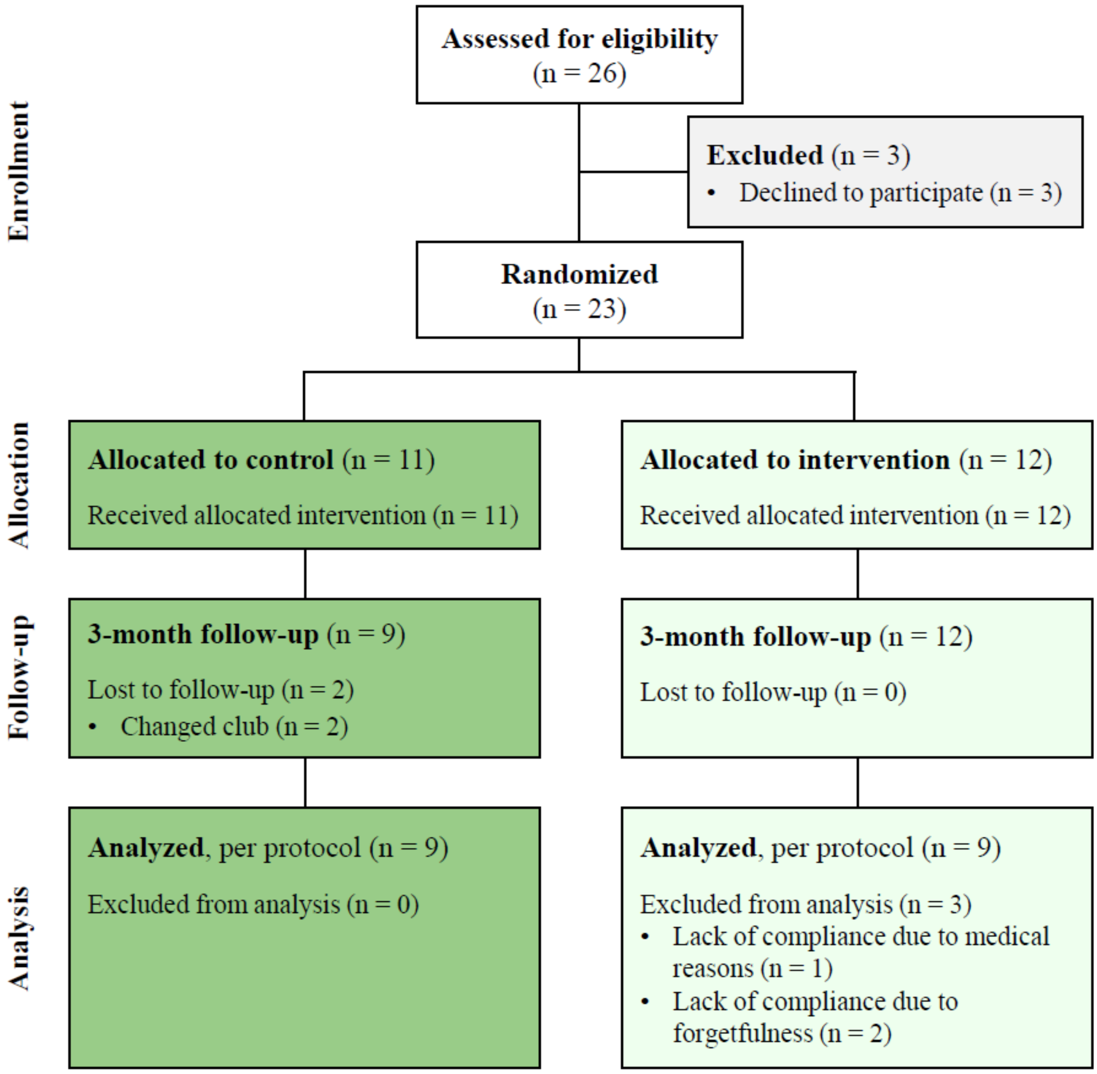

**Supplementary Figure S18**. Participant flow diagram. A total of 23 female soccer players volunteered to participate and were randomized to either a control group (n = 11) without iron supplementation or an intervention group (n = 12) with iron supplementation. Prior to the 3-month follow-up, two participants resigned from the control group. For the per protocol analysis, three participants were excluded from the intervention group due to lack of compliance with the intervention, leaving nine participants in each group.

**Supplementary Table S19**. Estimated within-group changes in serum biomarkers from baseline to 3-month follow-up.

| | Control (n = 9) | | Intervention (n = 9) | |
|---|---|---|---|---|
| | **β estimate** | ***P*-value** | **β estimate** | ***P*-value** |
| **Iron status** | | | | |
| Ferritin (µg/L) | –2.6 [–14.5, 9.3] | 0.63 | 9.2 [–1.2, 19.7] | 0.08 |
| Iron[1] (µmol/L) | 1.4 [–1.5, 4.3] | 0.30 | 9.1 [3.7, 14.6] | **0.006** |
| Transferrin saturation (%) | 2.8 [–1.7, 7.2] | 0.19 | 12.0 [3.5, 20.5] | **0.01** |
| TIBC (µmol/L) | –0.9 [–4.6, 2.7] | 0.58 | –3.3 [–6.3, –0.4] | **0.03** |
| Hemoglobin (g/dL) | 0.2 [0.01, 0.4] | **0.05** | –0.03 [–0.5, 0.4] | 0.87 |
| Red blood cells (x$10^{12}$/L) | 0.1 [0.01, 0.2] | **0.03** | –0.03 [–0.2, 0.1] | 0.75 |
| Hepcidin (ng/mL) | 3.8 [–2.7, 10.3] | 0.21 | 2.5 [–1.6, 6.6] | 0.20 |
| **Liver damage** | | | | |
| ASAT (U/L) | –3.4 [–5.8, –1.1] | **0.01** | –5.2 [–9.5, –0.9] | **0.02** |
| ALAT (U/L) | –0.3 [–3.4, 2.8] | 0.81 | –3.4 [–6.9, –0.004] | **0.05** |
| GGT (U/L) | –0.2 [–2.6, 2.1] | 0.83 | –0.3 [–3.2, 2.5] | 0.79 |
| **Inflammation** | | | | |
| CRP (mg/L) | –0.3 [–0.6, 0.1] | 0.14 | –0.1 [–0.3, 0.1] | 0.36 |
| IL-6 (pg/mL) | 0.1 [–0.3, 0.5] | 0.73 | –0.1 [–0.3, 0.01] | 0.06 |
| IL-8 (pg/mL) | –0.4 [–1.5, 0.7] | 0.39 | –0.5 [–1.2, 0.2] | 0.12 |
| TNF-α (pg/mL) | –1.4 [–2.5, –0.2] | **0.02** | –1.9 [–3.3, –0.6] | **0.01** |
| IFN-γ (pg/mL) | –0.04 [–0.2, 0.2] | 0.64 | 0.2 [–0.1, 0.4] | 0.13 |
| NGAL (ng/mL) | –2.9 [–16.6, 10.8] | 0.64 | –12.6 [–38.2, 13.1] | 0.29 |
| LBP (µg/mL) | 0.3 [–2.5, 3.1] | 0.81 | 1.0 [–1.8, 3.9] | 0.43 |

β estimates with 95% CI and corresponding *P*-values are derived from per protocol paired samples t-tests. The β estimate represents the change from baseline to 3-month follow-up with a positive estimate indicating a higher value at follow-up compared to baseline. Statistically significant *P*-values are shown in bold ($P < 0.05$).

Abbreviations: ALAT, alanine aminotransferase; ASAT, aspartate aminotransferase; CRP, C-reactive protein; GGT, gamma-glutamyl transferase; IFN-γ, interferon-gamma; IL, interleukin; LBP, lipopolysaccharide-binding protein; NGAL, neutrophil gelatinase-associated lipocalin; TIBC, total iron-binding capacity; TNF-α, tumor necrosis factor-alpha.

[1] One participant in the intervention group was excluded from the analysis (when included. $P = 0.02$).

**Supplementary Table S20**. Estimated within-group changes in fecal microbial diversity, taxa relative abundances and biomarkers from baseline to 3-month follow-up.

| | Control (n = 9) | | Intervention (n = 9) | |
|---|---|---|---|---|
| | **β estimate** | ***P*-value** | **β estimate** | ***P*-value** |
| **Alpha diversity** | | | | |
| Shannon diversity | –0.1 [–0.4, 0.2] | 0.41 | 0.1 [–0.01, 0.2] | 0.07 |
| Inverse Simpson | –5.3 [–13.2, 2.5] | 0.16 | 7.2 [–0.5, 14.9] | 0.06 |
| **Taxa abundances**[1] | | | | |
| *f_Dialisteraceae* | –0.44 [–0.73, –0.15] | **0.008** | –0.02 [–0.41, 0.37] | 0.90 |
| *f_Eggerthellaceae* | –0.12 [–0.20, –0.03] | **0.01** | –0.02 [–0.06, 0.03] | 0.38 |
| *f_Erysipelotrichaceae* | 0.03 [–0.01, 0.13] | 0.10 | –0.07 [–0.25, 0.005] | 0.06 |
| *g_Bariatricus* | 0.07 [–0.002, 0.14] | 0.06 | –0.04 [–0.08, 0.001] | 0.06 |
| *g_Blautia_A* | –0.04 [–0.19, 0.11] | 0.54 | 0.12 [0.02, 0.21] | **0.02** |
| *g_CAG-177* | 0.15 [–0.05, 0.34] | 0.12 | –0.18 [–0.32, –0.04] | **0.02** |
| *g_Coprococcus* | –0.25 [–0.46, –0.04] | **0.03** | 0.26 [–0.10, 0.62] | 0.13 |
| *g_Dialister* | –0.44 [–0.72, –0.15] | **0.008** | –0.02 [–0.42, 0.38] | 0.92 |
| *g_Dysosmobacter* | 0.07 [0.03, 0.11] | **0.006** | 0.08 [0.005, 0.15] | **0.04** |
| *g_Enterocloster* | 0.02 [–0.05, 0.08] | 0.57 | –0.07 [–0.21, 0.01] | 0.06 |
| *g_Faecousia* | –0.15 [–0.26, –0.03] | **0.02** | 0.05 [–0.02, 0.11] | 0.13 |
| **SCFA (mmol/kg)** | | | | |
| Acetic acid | –2.7 [–13.4, 8.0] | 0.58 | 1.1 [–6.0, 8.2] | 0.73 |
| Propionic acid | –1.5 [–4.6, 1.6] | 0.30 | 3.3 [–1.0, 7.7] | 0.12 |
| Butyric acid[2] | –1.0 [–4.9, 3.0] | 0.59 | 0.7 [–1.6, 2.9] | 0.52 |
| Valeric acid | 0.1 [–0.7, 0.8] | 0.85 | 0.5 [–0.2, 1.1] | 0.13 |
| Caproic acid | –0.5 [–1.1, 0.2] | 0.16 | 0.1 [–0.5, 0.6] | 0.79 |
| Total iso-SCFA | 0.9 [–1.1, 3.0] | 0.34 | 0.1 [–2.2, 2.4] | 0.90 |
| Total SCFA | –1.4 [–19.8, 17.1] | 0.87 | 5.7 [–6.0, 17.5] | 0.29 |
| **Gut inflammation** | | | | |
| NGAL (ng/g) | 0.2 [–17.1, 17.5] | 0.98 | 4.2 [–35.6, 44.0] | 0.81 |

β estimates with 95% CI and corresponding *P*-values are derived from per protocol paired samples t-tests. The β estimate represents the change from baseline to 3-month follow-up with a positive estimate indicating a higher value at follow-up compared to baseline. Statistically significant *P*-values are shown in bold ($P < 0.05$).

Abbreviations: NGAL, neutrophil gelatinase-associated lipocalin; SCFA, short-chain fatty acids.

[1] Taxa relative abundances (%) were square-root-transformed prior to analysis and reported β estimates and *P*-values are based on transformed data, except for *f_Erysipelotrichaceae* and *g_Enterocloster* which were not transformed prior to analysis. These were analyzed using paired Wilcoxon signed rank test. Prefixes indicate taxonomic rank: *f_* for family, *g_* for genus.

[2] One participant in the control group was excluded from the analysis (when included, $P = 0.54$).

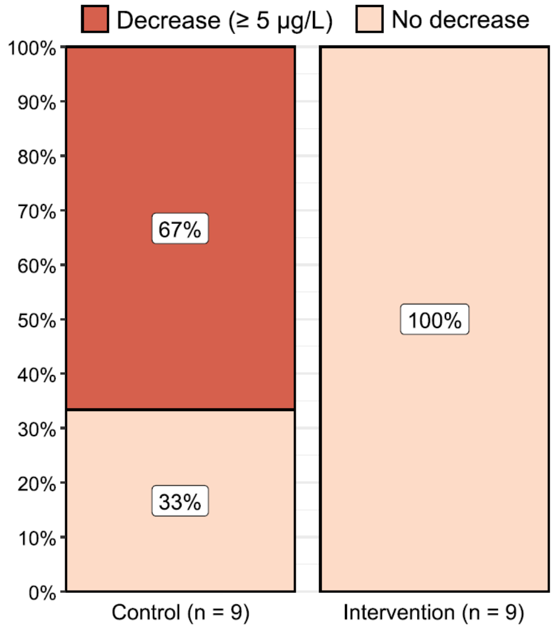

**Supplementary Figure S21**. Percentage of participants (n = 18) in each group showing a decline in serum ferritin of ≥ 5 µg/L (dark red) or not (light red) from baseline to 3-month follow-up.

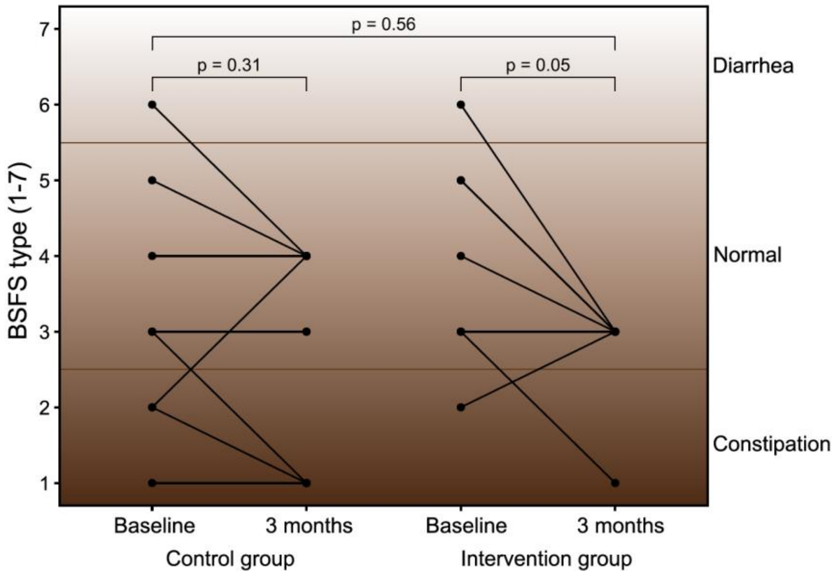

**Supplementary Figure S22**. Effects of low-dose iron supplementation on BSFS type (n = 17). Background shading reflects stool consistency with darker brown indicating firmer stools (constipation) and lighter brown indicating looser stools (diarrhea). The fecal category associated with each BSFS type is shown to the right (types 1–2, constipation; types 3–5, normal; types 6–7, diarrhea) (14). The *P*-values are from per protocol ANCOVA with baseline adjustment (between-group comparison) and paired samples t-tests (within-group comparison). One participant in the intervention group was excluded from the analysis because two different BSFS types were reported at follow-up. BSFS, Bristol Stool Form Scale.

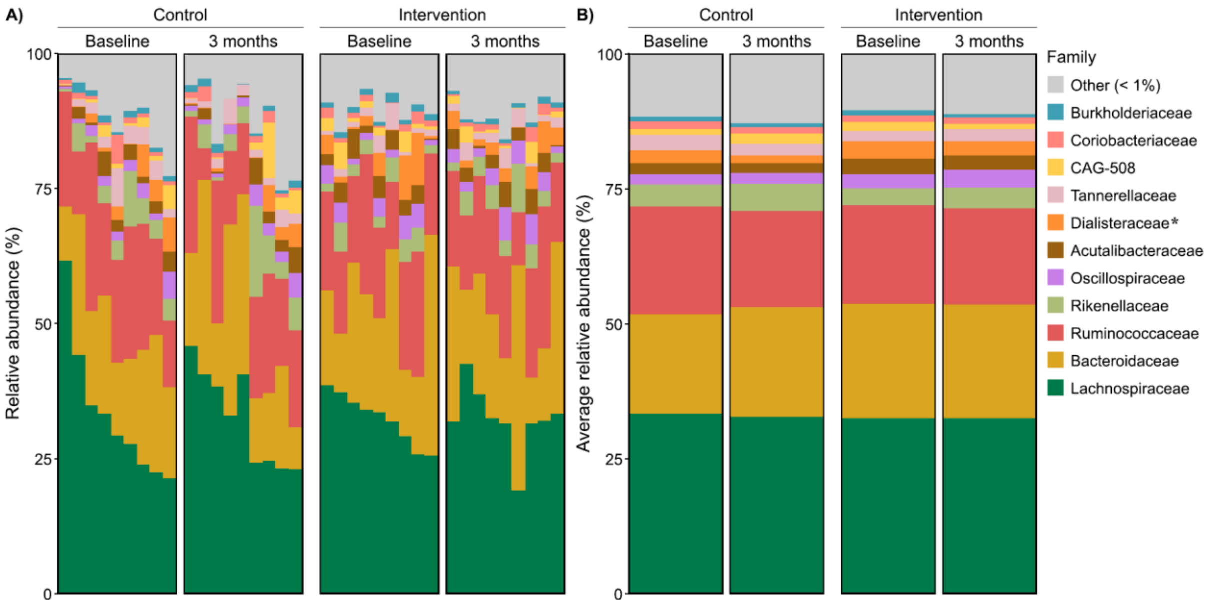

**Supplementary Figure S23**. Fecal relative abundances (%) of bacterial families at baseline and 3-month follow-up. (A) Each bar represents one participant. Within groups, bars are sorted in descending order according to the most abundant family at baseline (*Lachnospiraceae*), and this order is preserved at follow-up. (B) Group means are displayed as average relative abundances at each visit. Families with mean relative abundance < 1% are shown as "Other." Taxa are ordered by overall mean abundance, except "Other" which appear at the top regardless of abundance. Taxa that were significantly different between groups at follow-up (per protocol ANCOVA with baseline adjustment; uncorrected for multiple testing) are marked with an asterisk (*).

**Supplementary Table S24**. PERMANOVA results on Bray-Curtis dissimilarities (n = 18). Analyses were performed on the full dataset (including an interaction term), separately at each visit to assess between-group differences, and separately for each study group to assess within-group differences.

| | Sum of squares | $R^2$ | F statistic | *P*-value |
|---|---|---|---|---|
| **Full model**[1] | | | | |
| Study group | 0.34 | 0.04 | 1.52 | 0.24 |
| Visit | 0.06 | 0.01 | 0.26 | 0.45 |
| Study group × Visit | 0.07 | 0.01 | 0.31 | 0.16 |
| **Separately at each visit**[2] | | | | |
| Baseline, Study group | 0.16 | 0.04 | 0.70 | 0.99 |
| Follow-up, Study group | 0.25 | 0.07 | 1.13 | 0.22 |
| **Separately for each study group**[3] | | | | |
| Control, Visit | 0.09 | 0.02 | 0.34 | 0.99 |
| Intervention, Visit | 0.04 | 0.01 | 0.21 | > 0.99 |

[1] Model: distance matrix ~ study group × visit.

[2] Model: distance matrix ~ study group, performed separately at baseline and 3-month follow-up.

[3] Model: distance matrix ~ visit, performed separately for the control and intervention group.

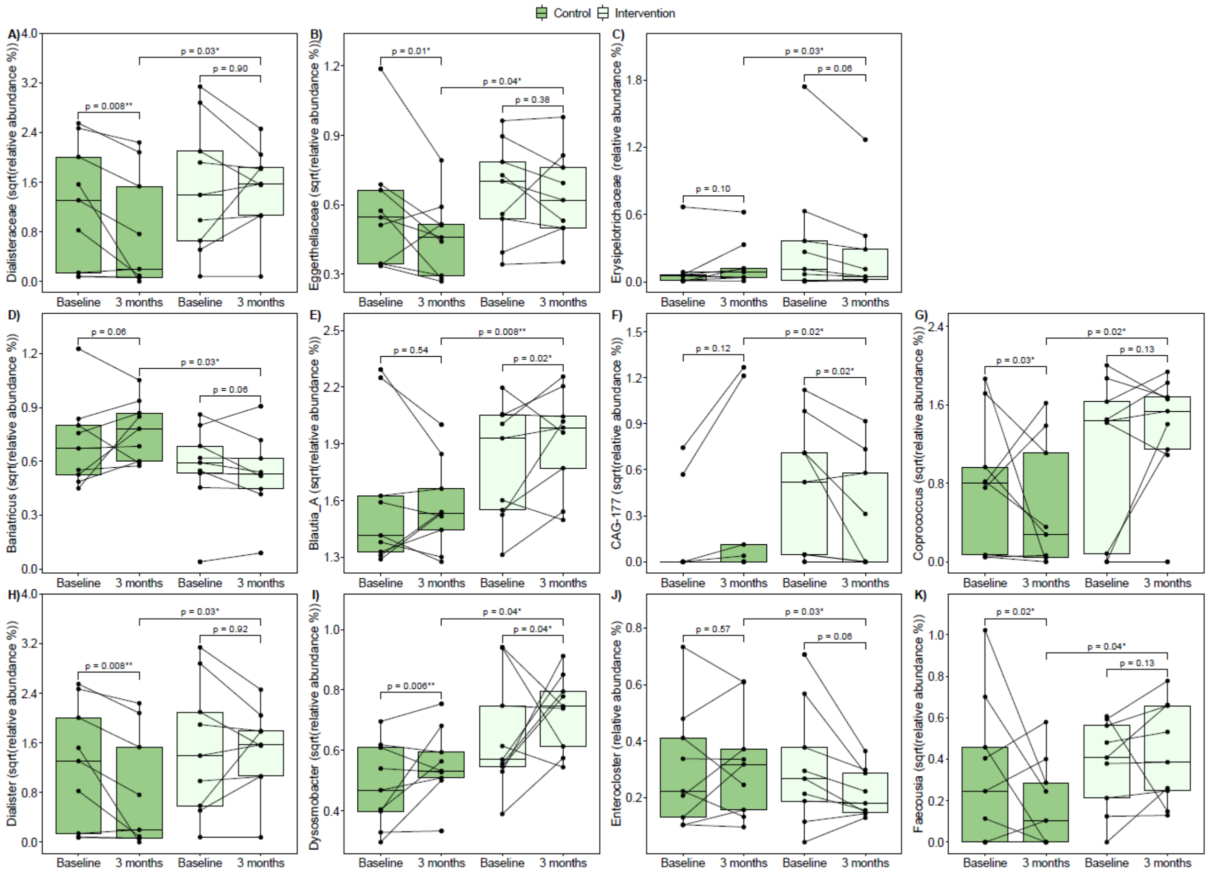

**Supplementary Figure S25**. Effects of low-dose iron supplementation on taxa relative abundances (%). (A) *Dialisteraceae* family, (B) *Eggerthellaceae* family, (C) *Erysipelotrichaceae* family, (D) *Bariatricus* genus, (E) *Blautia_A* genus, (F) *CAG-177* genus, (G) *Coprococcus* genus, (H) *Dialister* genus, (I) *Dysosmobacter* genus, (J) *Enterocloster* genus and (K) *Faecousia* genus. Each dot represents one sample, with lines connecting samples from the same participant. The *P*-values are from per protocol ANCOVA with baseline adjustment (between-group comparison) and paired samples t-tests (within-group comparison). Taxa relative abundances were square-root-transformed (sqrt) prior to analysis except for *Erysipelotrichaceae* and *Enterocloster* in which untransformed relative abundances were used.

**Supplementary Table S26**. Taxa that differed significantly between study groups at 3-month follow-up prior to correction for multiple testing, based on untransformed and square-root-transformed data.

| | **$\beta_{group}$** | **$P_{group}$** | **FDR-corrected $P_{group}$** |
|---|---|---|---|
| **Untransformed relative abundance (%)** | | | |
| *f_Eggerthellaceae* | 0.17 [0.01, 0.32] | **0.04** | 0.58 |
| *f_Erysipelotrichaceae* | –0.09 [–0.17, –0.01] | **0.03** | 0.58 |
| *g_Bariatricus* | –0.20 [–0.38, –0.02] | **0.04** | 0.49 |
| *g_Blautia_A* | 0.91 [0.30, 1.52] | **0.006** | 0.49 |
| *g_CAG-177* | –0.44 [–0.87, –0.01] | **0.05** | 0.49 |
| *g_Coprococcus* | 1.25 [0.17, 2.33] | **0.03** | 0.49 |
| *g_Dysosmobacter* | 0.22 [0.04, 0.41] | **0.02** | 0.49 |
| *g_Enterocloster* | –0.11 [–0.20, –0.01] | **0.03** | 0.49 |
| **Sqrt-transformed relative abundance (%)** | | | |
| *f_Dialisteraceae* | 0.52 [0.05, 0.99] | **0.03** | 0.55 |
| *f_Eggerthellaceae* | 0.13 [0.01, 0.26] | **0.04** | 0.55 |
| *g_Bariatricus* | –0.16 [–0.29, –0.02] | **0.03** | 0.48 |
| *g_Blautia_A* | 0.24 [0.07, 0.41] | **0.008** | 0.48 |
| *g_CAG-177* | –0.33 [–0.62, –0.05] | **0.02** | 0.48 |
| *g_Coprococcus* | 0.70 [0.13, 1.28] | **0.02** | 0.48 |
| *g_Dialister* | 0.52 [0.05, 0.98] | **0.03** | 0.48 |
| *g_Dysosmobacter* | 0.15 [0.01, 0.30] | **0.04** | 0.48 |
| *g_Faecousia* | 0.23 [0.02, 0.44] | **0.04** | 0.48 |

β estimates with 95% CI and corresponding *P*-values are derived from per protocol ANCOVA adjusted for baseline values of the outcome. The β estimate represents the between-group difference at follow-up with a positive estimate indicating a higher value in the intervention group compared to the control group. Prefixes indicate taxonomic rank: *f_* for family, *g_* for genus. Statistically significant *P*-values are shown in bold ($P < 0.05$).

Abbreviations: FDR, false discovery rate; sqrt, square root.

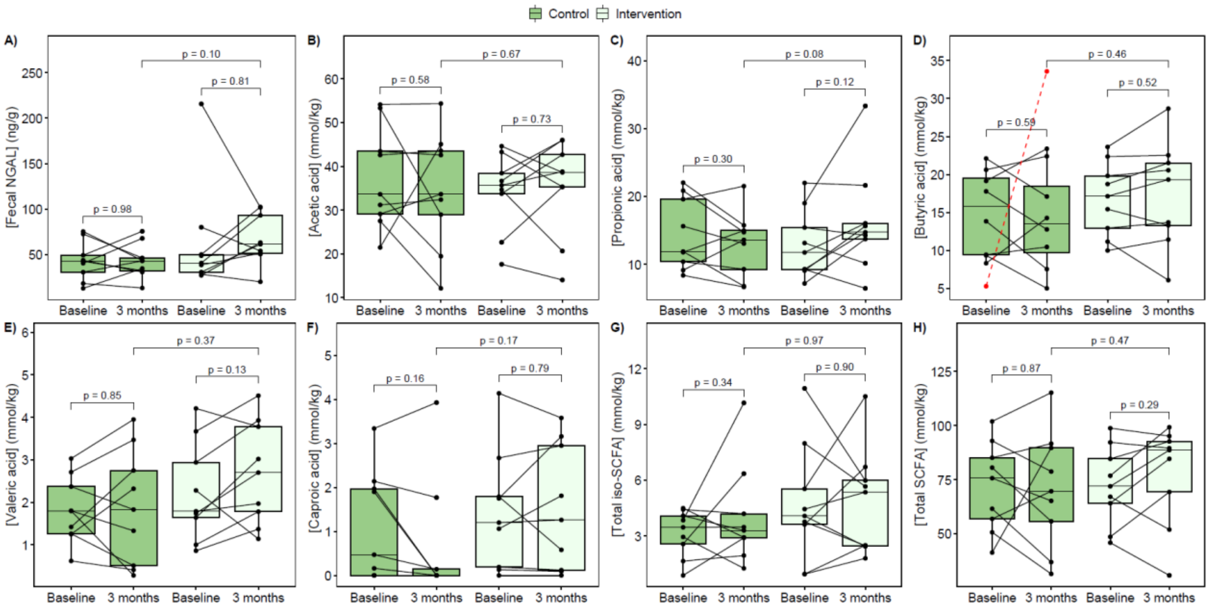

**Supplementary Figure S27**. Effects of low-dose iron supplementation on gut inflammation and microbial metabolites. (A) Fecal NGAL, (B) acetic acid, C) propionic acid, (D) butyric acid (participant marked in red was excluded from the statistical analysis), (E) valeric acid, (F) caproic acid, (G) total iso-SCFA (iso-butyric, iso-valeric and iso-caproic acid) and (H) total SCFA concentration. Each dot represents one sample, with lines connecting samples from the same participant. The *P*-values are from per protocol ANCOVA with baseline adjustment (between-group comparison) and paired samples t-tests (within-group comparison). NGAL, neutrophil gelatinase-associated lipocalin; SCFA, short-chain fatty acids.

### 8. CORRELATION BETWEEN GUT MICROBIOTA AND OTHER STUDY OUTCOMES

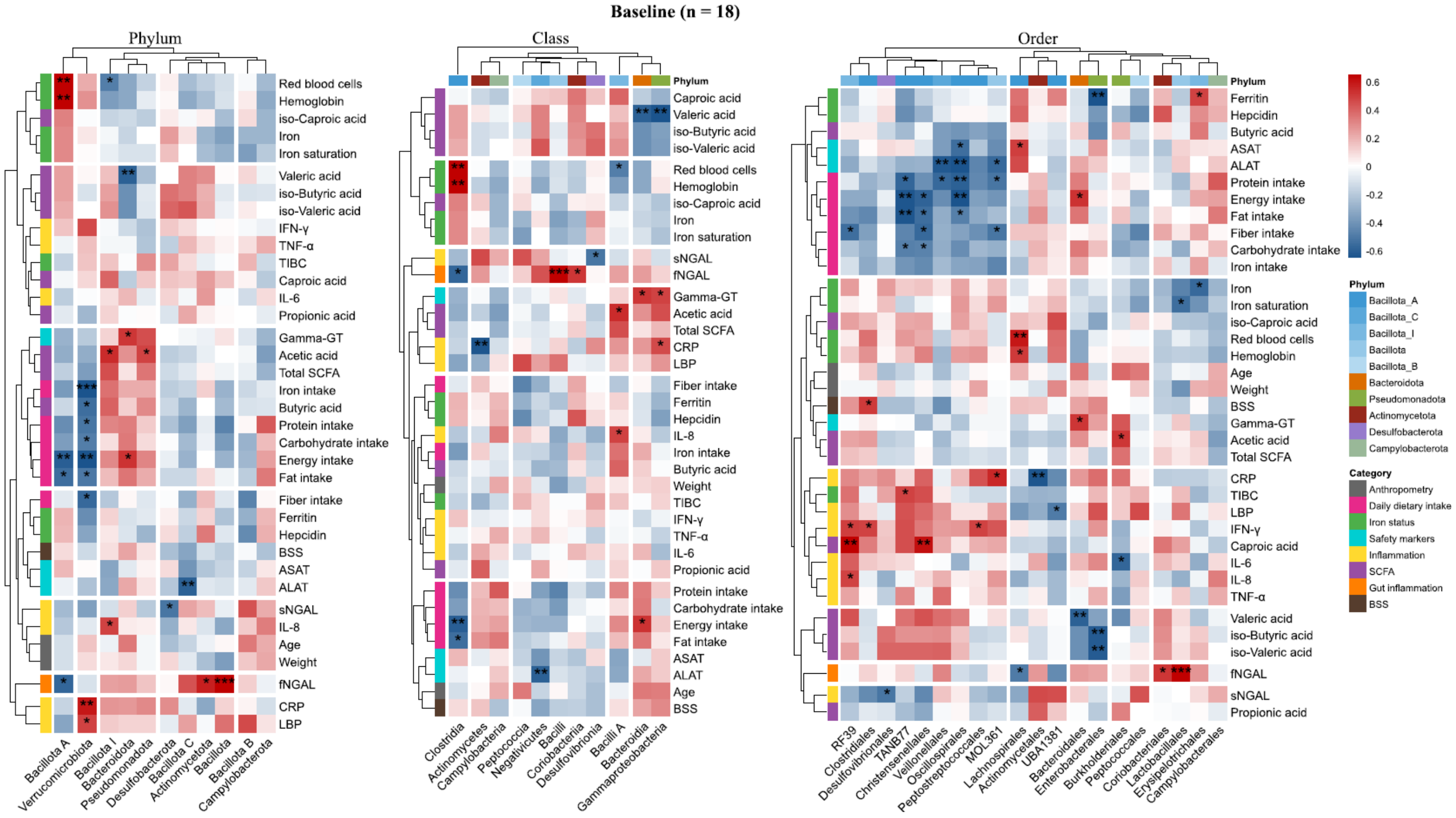

**Supplementary Figure S28**. Spearman's rank correlations at baseline (n = 18) between fecal relative abundances (phylum, class, order) and anthropometry, dietary intake, iron status, safety markers, inflammation, SCFA and BSS. The color scale represents correlation strength and direction (red, positive; blue, negative). Only taxa with a median relative abundance ≥ 0.01% in at least one study group were included. Significant correlations are indicated by asterisks (*$P < 0.05$, **$P < 0.01$, ***$P < 0.001$; uncorrected for multiple testing). ALAT, alanine aminotransferase; ASAT, aspartate aminotransferase; BSS, Bristol stool scale; CRP, C-reactive protein; Gamma-GT, gamma-glutamyl transferase; IFN-γ, interferon-gamma; IL, interleukin; LBP, lipopolysaccharide-binding protein; NGAL, neutrophil gelatinase-associated lipocalin; SCFA, short-chain fatty acids; TNF-α, tumor necrosis factor-alpha.

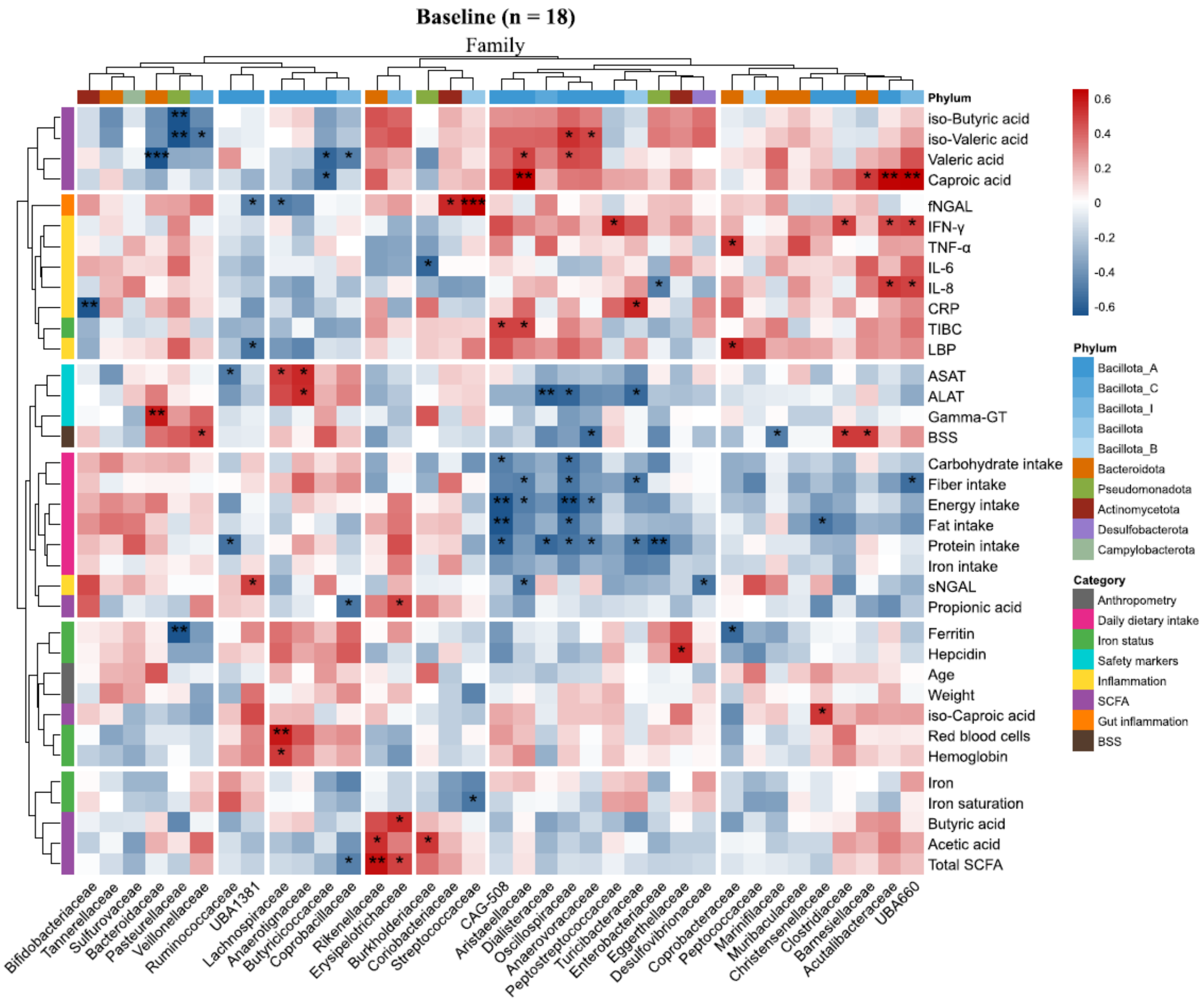

**Supplementary Figure S29**. Spearman's rank correlations at baseline (n = 18) between fecal bacterial families (relative abundance, %) and anthropometry, dietary intake, iron status, safety markers, inflammation, SCFA and BSS. The color scale represents correlation strength and direction (red, positive; blue, negative). Only taxa with a median relative abundance ≥ 0.01% in at least one study group were included. Significant correlations are indicated by asterisks (**P* < 0.05, ***P* < 0.01, ****P* < 0.001; uncorrected for multiple testing). ALAT, alanine aminotransferase; ASAT, aspartate aminotransferase; BSS, Bristol stool scale; CRP, C-reactive protein; Gamma-GT, gamma-glutamyl transferase; IFN-γ, interferon-gamma; IL, interleukin; LBP, lipopolysaccharide-binding protein; NGAL, neutrophil gelatinase-associated lipocalin; SCFA, short-chain fatty acids; TNF-α, tumor necrosis factor-alpha.

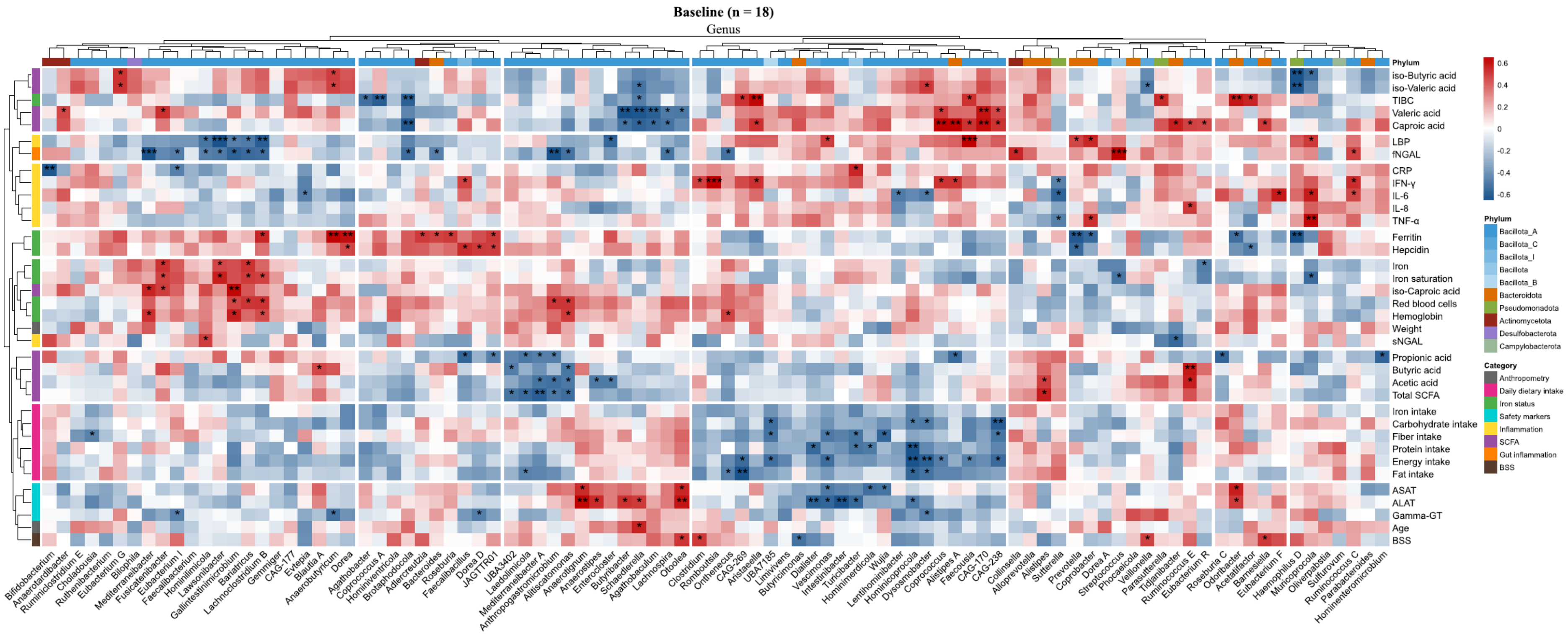

**Supplementary Figure S30**. Spearman's rank correlations at baseline (n = 18) between fecal bacterial genera (relative abundance, %) and anthropometry, dietary intake, iron status, safety markers, inflammation, SCFA and BSS. The color scale represents correlation strength and direction (red, positive; blue, negative). Only taxa with a median relative abundance ≥ 0.01% in at least one study group were included. Significant correlations are indicated by asterisks (*$P$ < 0.05, **$P$ < 0.01, ***$P$ < 0.001; uncorrected for multiple testing). ALAT, alanine aminotransferase; ASAT, aspartate aminotransferase; BSS, Bristol stool scale; CRP, C-reactive protein; Gamma-GT, gamma-glutamyl transferase; IFN-γ, interferon-gamma; IL, interleukin; LBP, lipopolysaccharide-binding protein; NGAL, neutrophil gelatinase-associated lipocalin; SCFA, short-chain fatty acids; TNF-α, tumor necrosis factor-alpha.

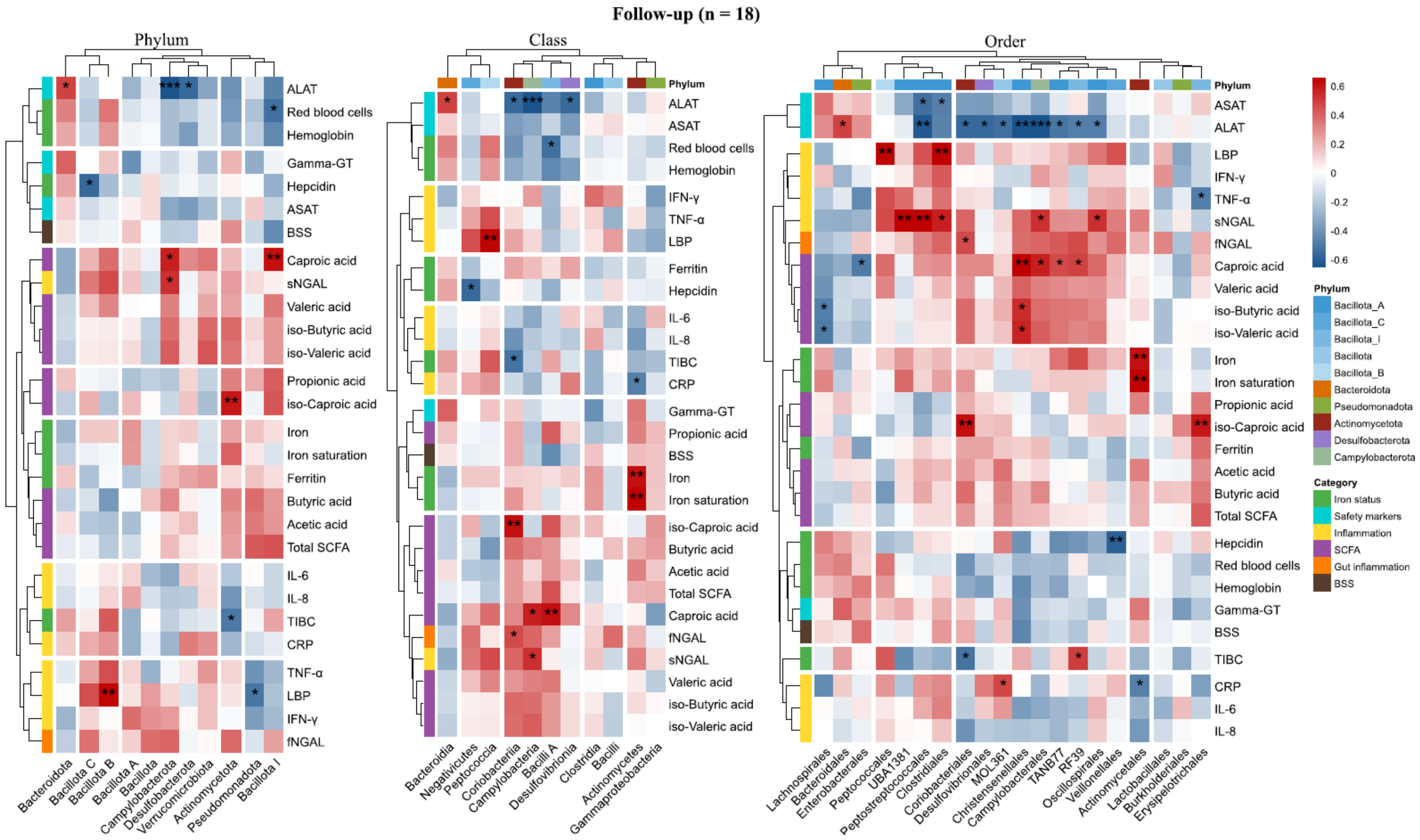

**Supplementary Figure S31**. Spearman's rank correlations at 3-month follow-up (n = 18) between fecal relative abundances (phylum, class, order) and iron status, safety markers, inflammation, SCFA and BSS. The color scale represents correlation strength and direction (red, positive; blue, negative). Only taxa with a median relative abundance ≥ 0.01% in at least one study group were included. Significant correlations are indicated by asterisks (**P* < 0.05, ***P* < 0.01, ****P* < 0.001; uncorrected for multiple testing). ALAT, alanine aminotransferase; ASAT, aspartate aminotransferase; BSS, Bristol stool scale; CRP, C-reactive protein; Gamma-GT, gamma-glutamyl transferase; IFN-γ, interferon-gamma; IL, interleukin; LBP, lipopolysaccharide-binding protein; NGAL, neutrophil gelatinase-associated lipocalin; SCFA, short-chain fatty acids; TNF-α, tumor necrosis factor-alpha.

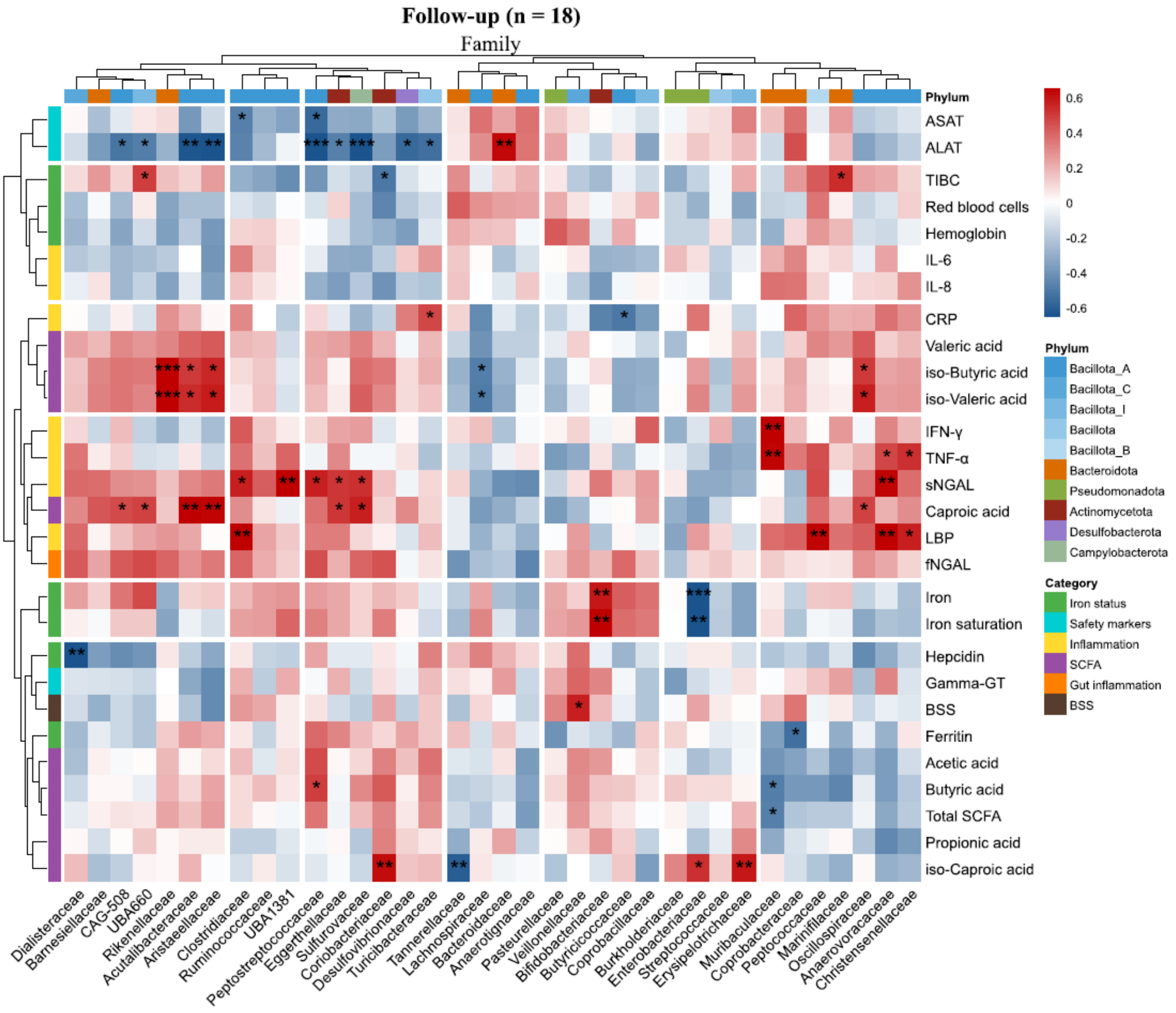

**Supplementary Figure S32**. Spearman's rank correlations at 3-month follow-up (n = 18) between fecal bacterial families (relative abundance, %) and iron status, safety markers, inflammation, SCFA and BSS. The color scale represents correlation strength and direction (red, positive; blue, negative). Only taxa with a median relative abundance ≥ 0.01% in at least one study group were included. Significant correlations are indicated by asterisks (*$P < 0.05$, **$P < 0.01$, ***$P < 0.001$; uncorrected for multiple testing). ALAT, alanine aminotransferase; ASAT, aspartate aminotransferase; BSS, Bristol stool scale; CRP, C-reactive protein; Gamma-GT, gamma-glutamyl transferase; IFN-γ, interferon-gamma; IL, interleukin; LBP, lipopolysaccharide-binding protein; NGAL, neutrophil gelatinase-associated lipocalin; SCFA, short-chain fatty acids; TNF-α, tumor necrosis factor-alpha.

Effects of low-dose iron supplementation on iron status, safety outcomes and gut microbiota in female soccer players: a randomized controlled study, Strømland et al.

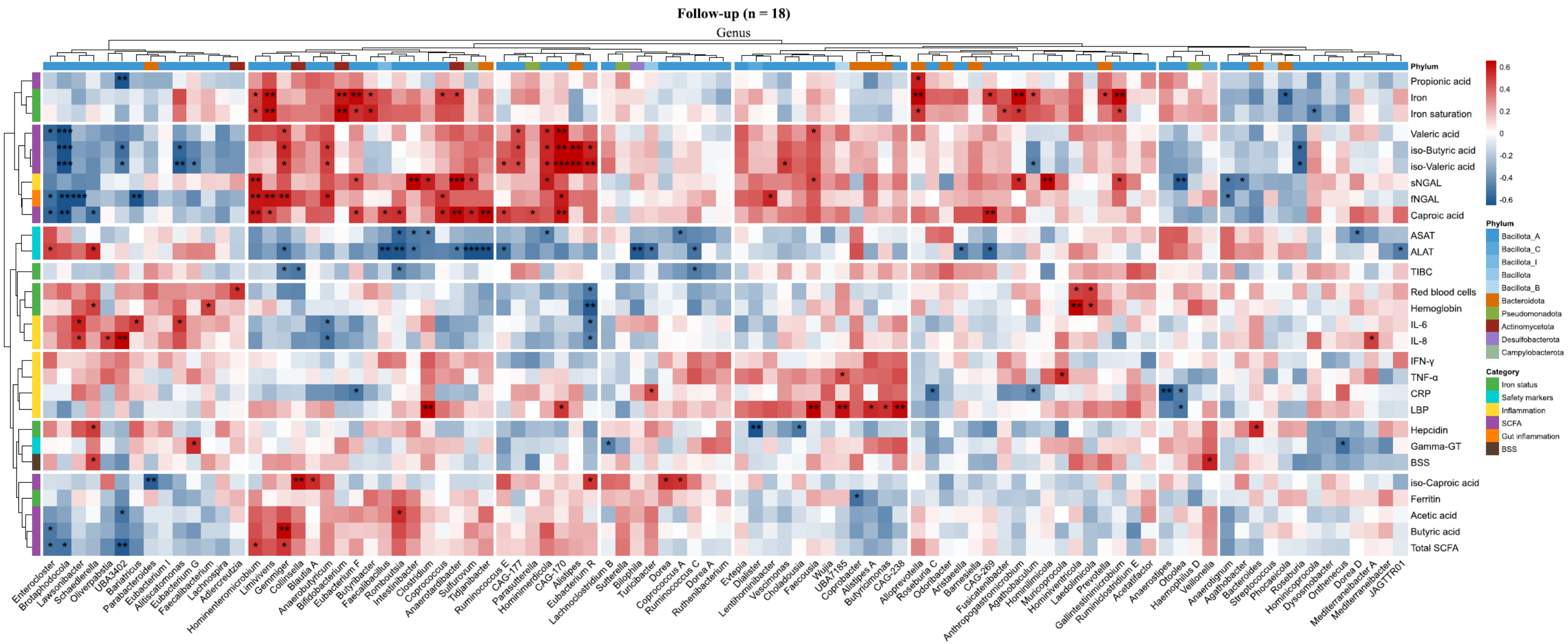

**Supplementary Figure S33**. Spearman's rank correlations at 3-month follow-up (n = 18) between fecal bacterial genera (relative abundance, %) and iron status, safety markers, inflammation, SCFA and BSS. The color scale represents correlation strength and direction (red, positive; blue, negative). Only taxa with a median relative abundance ≥ 0.01% in at least one study group were included. Significant correlations are indicated by asterisks (**P* < 0.05, ***P* < 0.01, ****P* < 0.001; uncorrected for multiple testing). ALAT, alanine aminotransferase; ASAT, aspartate aminotransferase; BSS, Bristol stool scale; CRP, C-reactive protein; Gamma-GT, gamma-glutamyl transferase; IFN-γ, interferon-gamma; IL, interleukin; LBP, lipopolysaccharide-binding protein; NGAL, neutrophil gelatinase-associated lipocalin; SCFA, short-chain fatty acids; TNF-α, tumor necrosis factor-alpha.

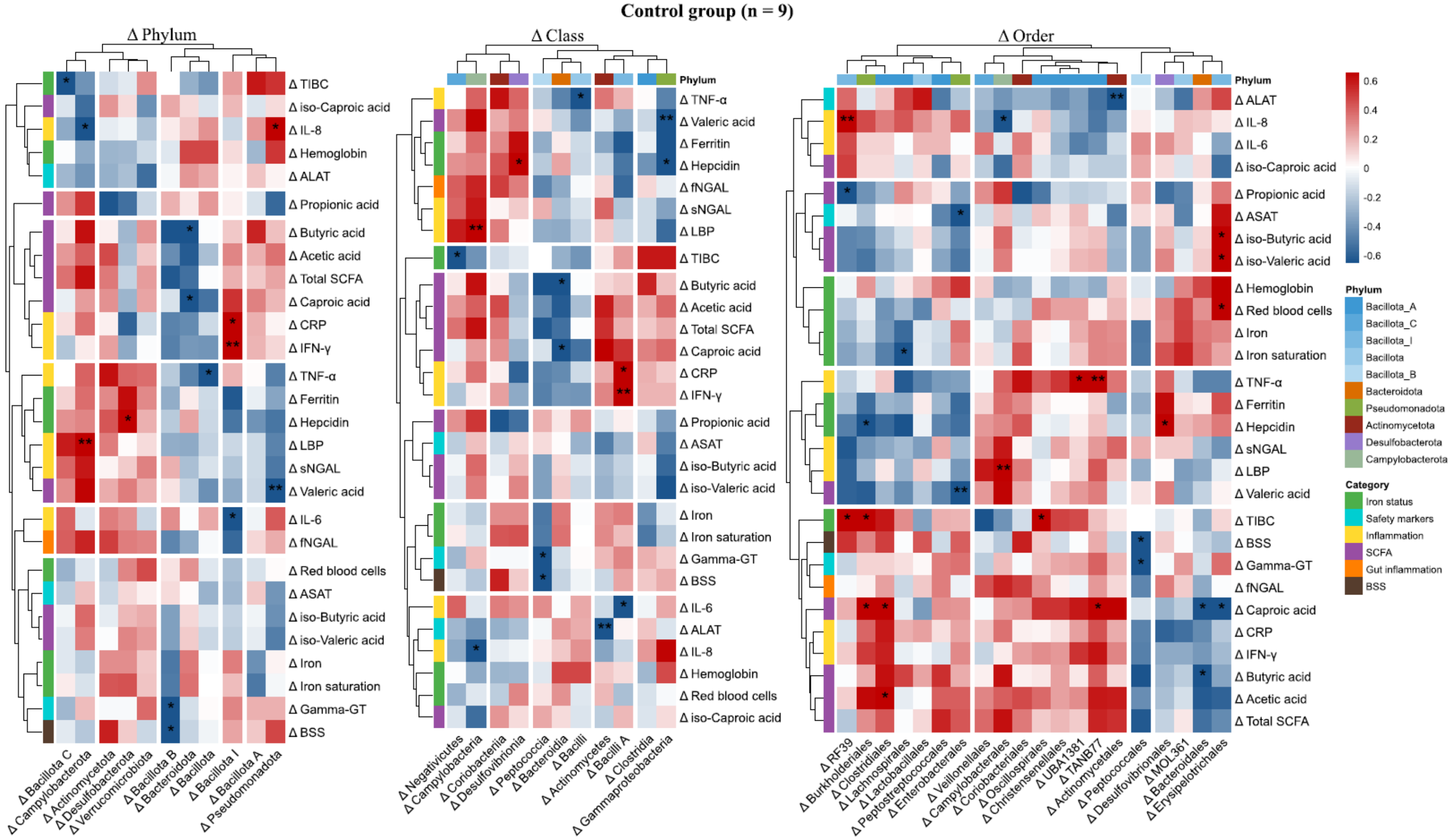

**Supplementary Figure S34**. Spearman's rank correlations in the control group (n = 9) between changes (Δ) from baseline to 3-month follow-up in fecal relative abundances (phylum, class, order) and iron status, safety markers, inflammation, SCFA and BSS. The color scale represents correlation strength and direction (red, positive; blue, negative). Only taxa with a median relative abundance ≥ 0.01% in at least one study group were included. Significant correlations are indicated by asterisks (**P* < 0.05, ***P* < 0.01, ****P* < 0.001; uncorrected for multiple testing). ALAT, alanine aminotransferase; ASAT, aspartate aminotransferase; BSS, Bristol stool scale; CRP, C-reactive protein; Gamma-GT, gamma-glutamyl transferase; IFN-γ, interferon-gamma; IL, interleukin; LBP, lipopolysaccharide-binding protein; NGAL, neutrophil gelatinase-associated lipocalin; SCFA, short-chain fatty acids; TNF-α, tumor necrosis factor-alpha.

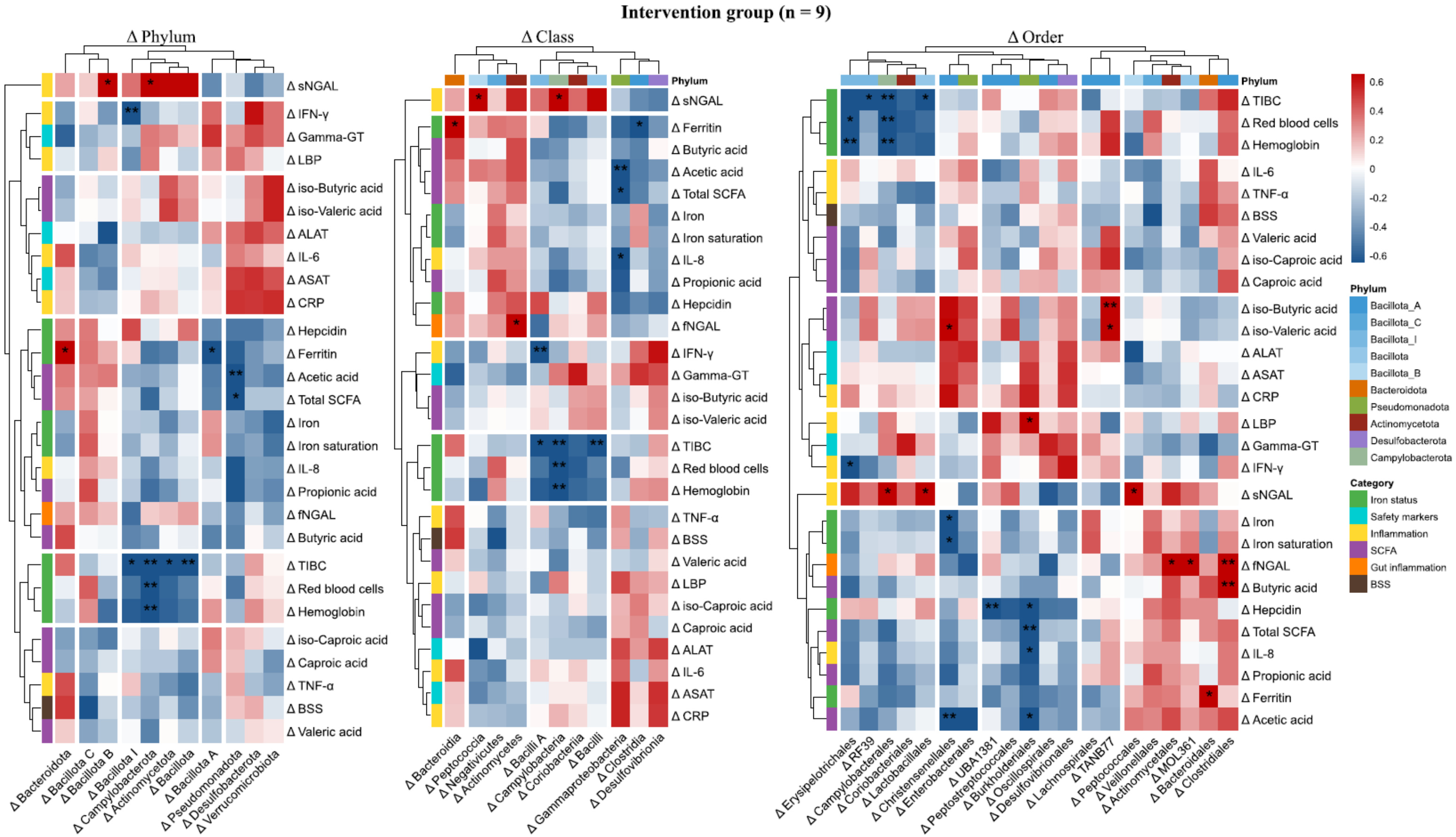

**Supplementary Figure S35**. Spearman's rank correlations in the intervention group (n = 9) between changes (Δ) from baseline to 3-month follow-up in fecal relative abundances (phylum, class, order) and iron status, safety markers, inflammation, SCFA and BSS. The color scale represents correlation strength and direction (red, positive; blue, negative). Only taxa with a median relative abundance ≥ 0.01% in at least one study group were included. Significant correlations are indicated by asterisks (**P* < 0.05, ***P* < 0.01, ****P* < 0.001; uncorrected for multiple testing). ALAT, alanine aminotransferase; ASAT, aspartate aminotransferase; BSS, Bristol stool scale; CRP, C-reactive protein; Gamma-GT, gamma-glutamyl transferase; IFN-γ, interferon-gamma; IL, interleukin; LBP, lipopolysaccharide-binding protein; NGAL, neutrophil gelatinase-associated lipocalin; SCFA, short-chain fatty acids; TNF-α, tumor necrosis factor-alpha.

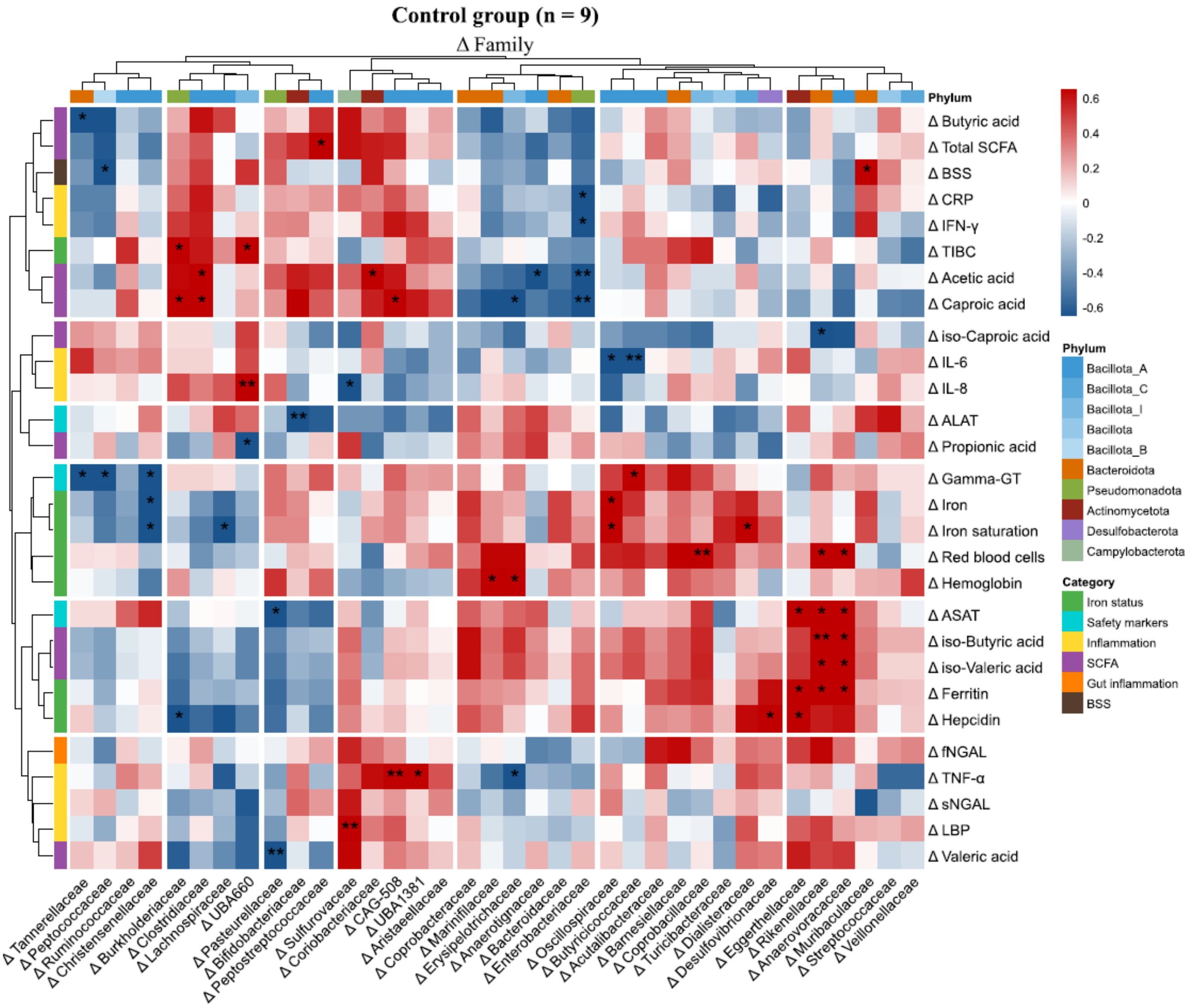

**Supplementary Figure S36**. Spearman's rank correlations in the control group (n = 9) between changes (Δ) from baseline to 3-month follow-up in fecal bacterial families (relative abundance, %) and iron status, safety markers, inflammation, SCFA and BSS. The color scale represents correlation strength and direction (red, positive; blue, negative). Only taxa with a median relative abundance ≥ 0.01% in at least one study group were included. Significant correlations are indicated by asterisks (**P* < 0.05, ***P* < 0.01, ****P* < 0.001; uncorrected for multiple testing). ALAT, alanine aminotransferase; ASAT, aspartate aminotransferase; BSS, Bristol stool scale; CRP, C-reactive protein; Gamma-GT, gamma-glutamyl transferase; IFN-γ, interferon-gamma; IL, interleukin; LBP, lipopolysaccharide-binding protein; NGAL, neutrophil gelatinase-associated lipocalin; SCFA, short-chain fatty acids; TNF-α, tumor necrosis factor-alpha.

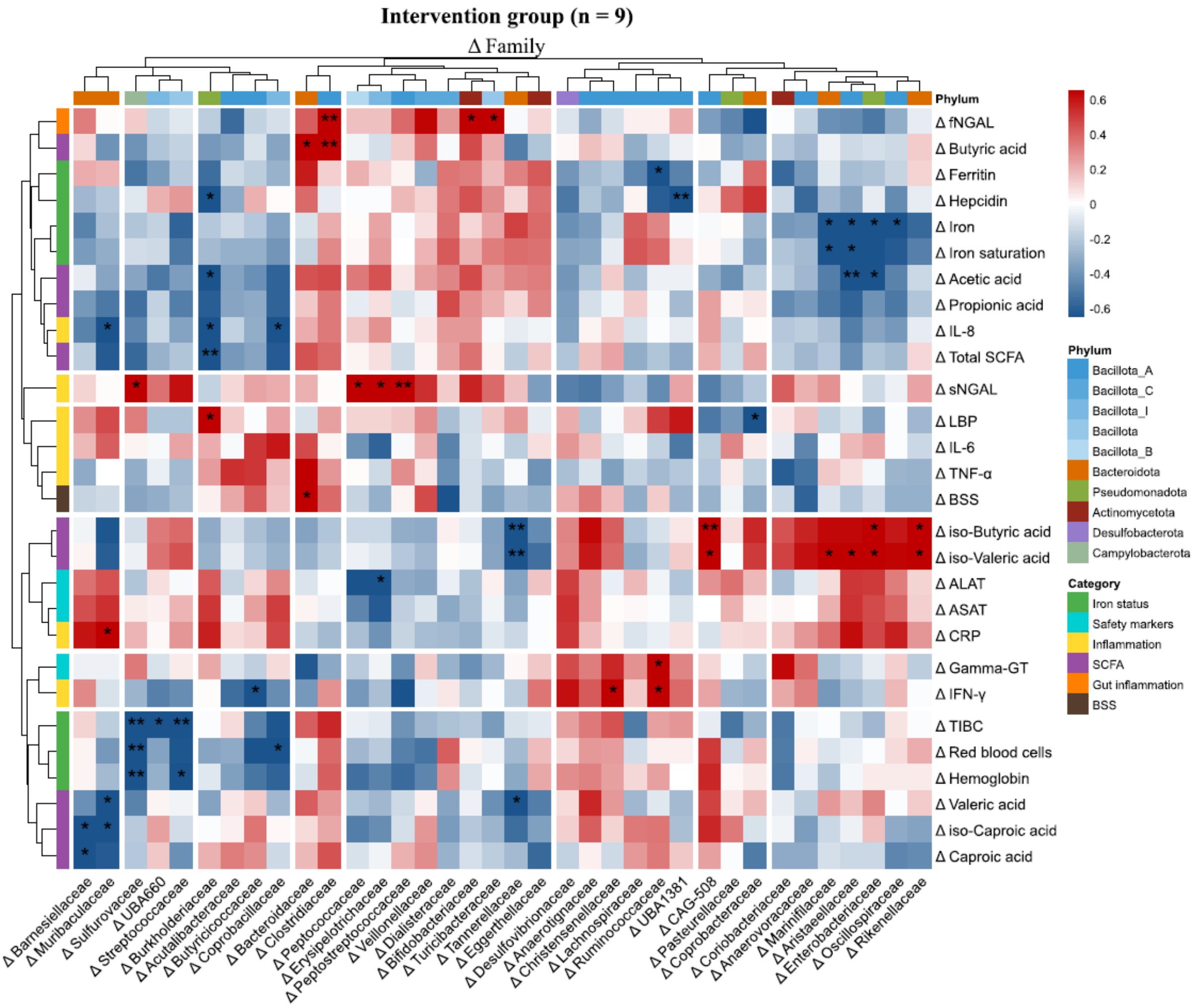

**Supplementary Figure S37**. Spearman's rank correlations in the intervention group (n = 9) between changes (Δ) from baseline to 3-month follow-up in fecal bacterial families (relative abundance, %) and iron status, safety markers, inflammation, SCFA and BSS. The color scale represents correlation strength and direction (red, positive; blue, negative). Only taxa with a median relative abundance ≥ 0.01% in at least one study group were included. Significant correlations are indicated by asterisks (**P* < 0.05, ***P* < 0.01, ****P* < 0.001; uncorrected for multiple testing). ALAT, alanine aminotransferase; ASAT, aspartate aminotransferase; BSS, Bristol stool scale; CRP, C-reactive protein; Gamma-GT, gamma-glutamyl transferase; IFN-γ, interferon-gamma; IL, interleukin; LBP, lipopolysaccharide-binding protein; NGAL, neutrophil gelatinase-associated lipocalin; SCFA, short-chain fatty acids; TNF-α, tumor necrosis factor-alpha.

Effects of low-dose iron supplementation on iron status, safety outcomes and gut microbiota in female soccer players: a randomized controlled study, Strømland et al.

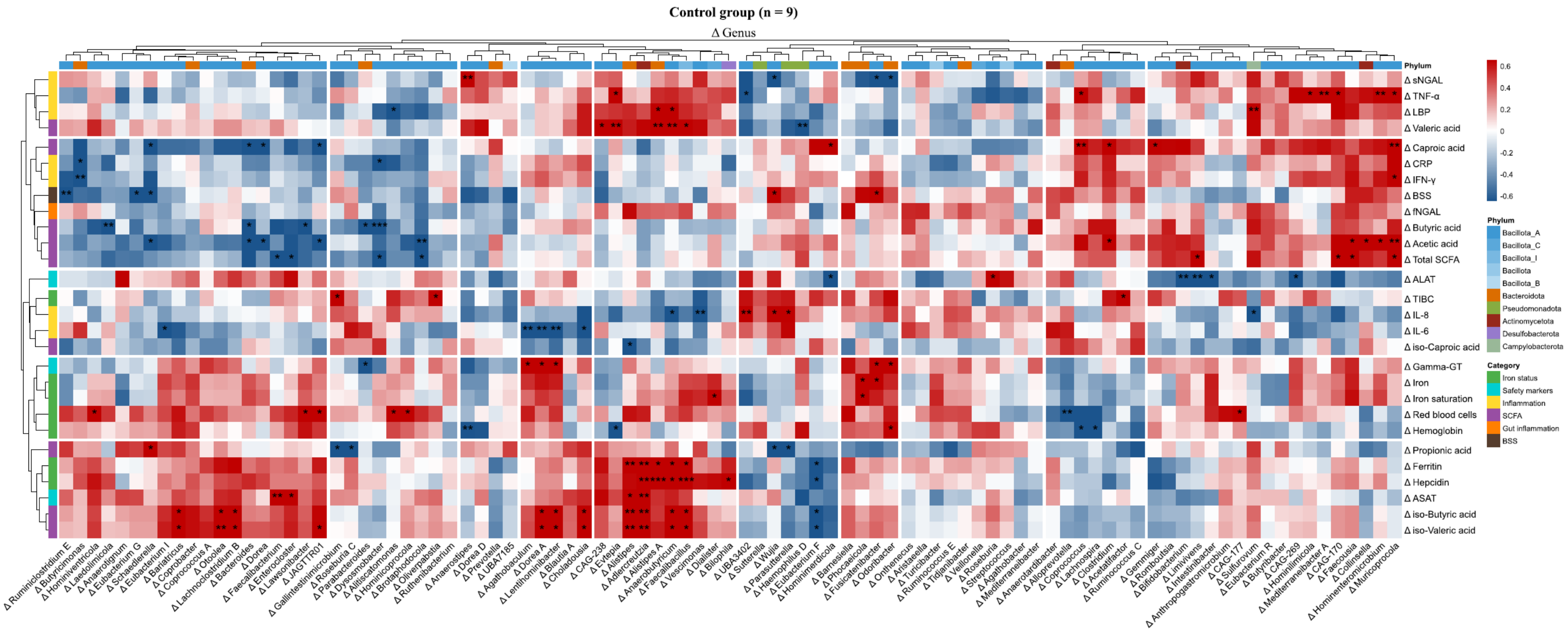

**Supplementary Figure S38**. Spearman's rank correlations in the control group (n = 9) between changes (Δ) from baseline to 3-month follow-up in fecal bacterial genera (relative abundance, %) and iron status, safety markers, inflammation, SCFA and BSS. The color scale represents correlation strength and direction (red, positive; blue, negative). Only taxa with a median relative abundance ≥ 0.01% in at least one study group were included. Significant correlations are indicated by asterisks (**P* < 0.05, ***P* < 0.01, ****P* < 0.001; uncorrected for multiple testing). ALAT, alanine aminotransferase; ASAT, aspartate aminotransferase; BSS, Bristol stool scale; CRP, C-reactive protein; Gamma-GT, gamma-glutamyl transferase; IFN-γ, interferon-gamma; IL, interleukin; LBP, lipopolysaccharide-binding protein; NGAL, neutrophil gelatinase-associated lipocalin; SCFA, short-chain fatty acids; TNF-α, tumor necrosis factor-alpha.

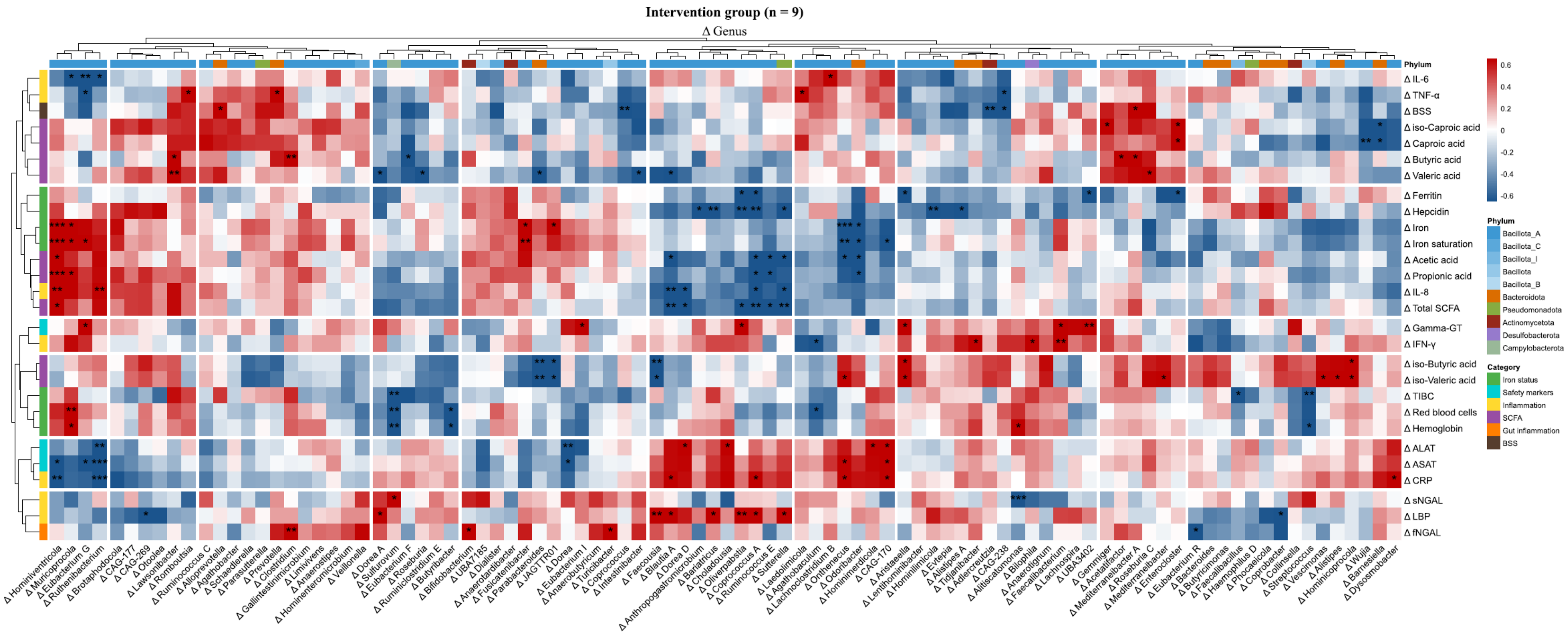

**Supplementary Figure S39**. Spearman's rank correlations in the intervention group (n = 9) between changes (Δ) from baseline to 3-month follow-up in fecal bacterial genera (relative abundance, %) and iron status, safety markers, inflammation, SCFA and BSS. The color scale represents correlation strength and direction (red, positive; blue, negative). Only taxa with a median relative abundance ≥ 0.01% in at least one study group were included. Significant correlations are indicated by asterisks (**P* < 0.05, ***P* < 0.01, ****P* < 0.001; uncorrected for multiple testing). ALAT, alanine aminotransferase; ASAT, aspartate aminotransferase; BSS, Bristol stool scale; CRP, C-reactive protein; Gamma-GT, gamma-glutamyl transferase; IFN-γ, interferon-gamma; IL, interleukin; LBP, lipopolysaccharide-binding protein; NGAL, neutrophil gelatinase-associated lipocalin; SCFA, short-chain fatty acids; TNF-α, tumor necrosis factor-alpha.

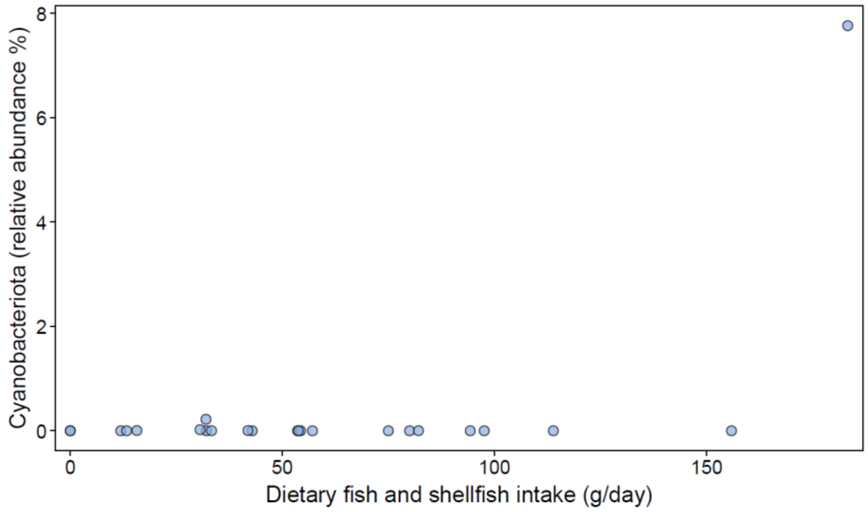

**Supplementary Figure S40**. Scatterplot of daily fish and shellfish intake (g/day) against the relative abundance (%) of the phylum *Cyanobacteriota* at baseline (n = 23). Each dot represents one participant.

### 9. CORRELATION BETWEEN CHANGES IN SERUM AND FECAL BIOMARKERS

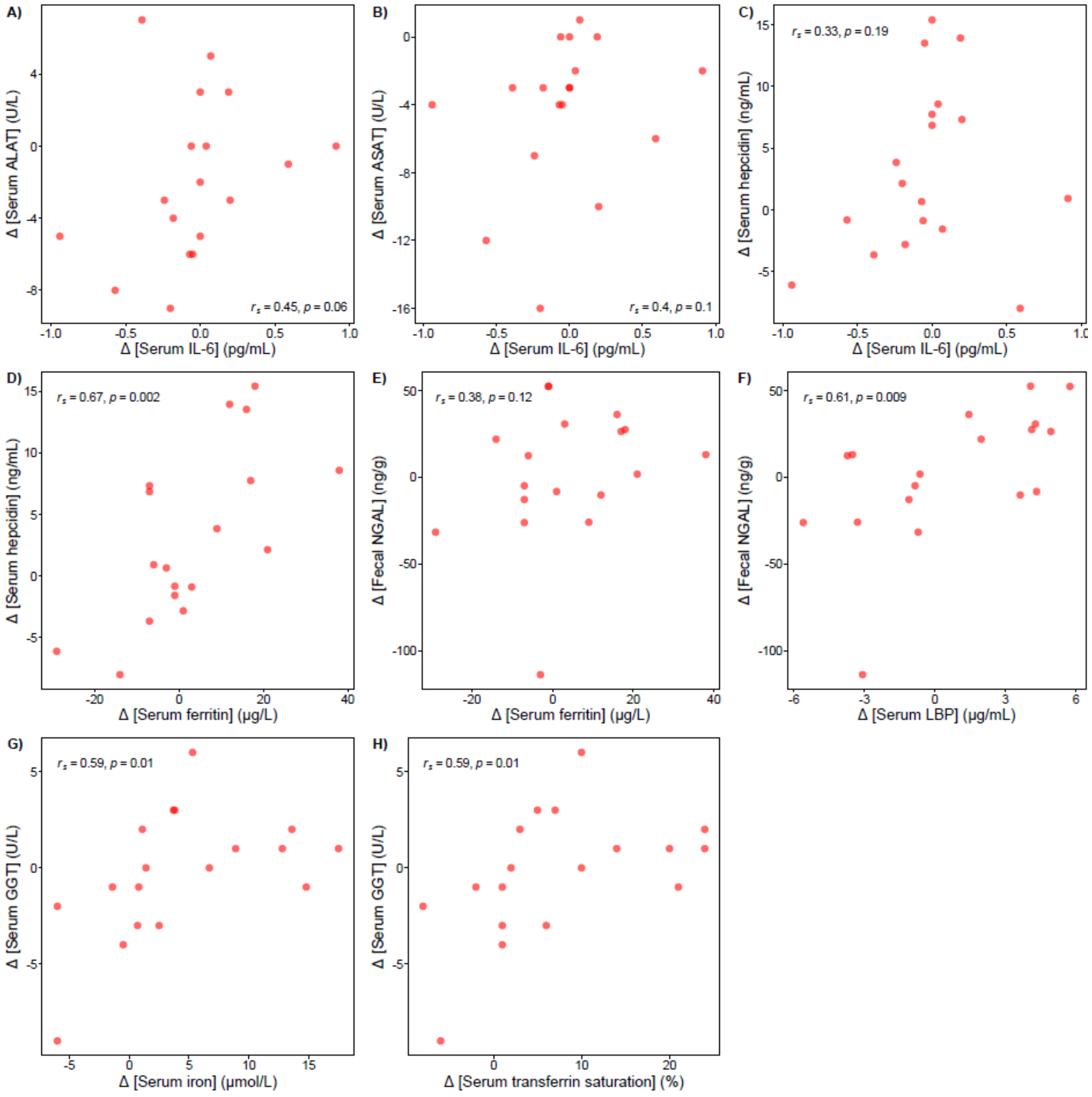

**Supplementary Figure S41.** Scatterplots showing Spearman's rank correlations between changes (Δ) in selected biomarkers from baseline to 3-month follow-up (n = 18). Changes were calculated by subtracting the baseline value from the follow-up value, with positive values indicating an increase over time. Spearman's correlations coefficients ($r_s$) with corresponding *P*-values are shown in each plot. ALAT, alanine aminotransferase; ASAT, aspartate aminotransferase; GGT, gamma-glutamyl transferase; IL, interleukin; LBP, lipopolysaccharide-binding protein; NGAL, neutrophil gelatinase-associated lipocalin.
